# Protective association of *HLA-DRB1*\*04 subtypes in neurodegenerative diseases implicates acetylated Tau PHF6 sequences

**DOI:** 10.1101/2021.12.26.21268354

**Authors:** Yann Le Guen, Guo Luo, Aditya Ambati, Vincent Damotte, Iris Jansen, Eric Yu, Aude Nicolas, Itziar de Rojas, Thiago Peixoto Leal, Akinori Miyashita, Céline Bellenguez, Michelle Mulan Lian, Kayenat Parveen, Takashi Morizono, Hyeonseul Park, Benjamin Grenier-Boley, Tatsuhiko Naito, Fahri Küçükali, Seth D. Talyansky, Selina Maria Yogeshwar, Vicente Sempere, Wataru Satake, Victoria Alvarez, Beatrice Arosio, Michael E. Belloy, Luisa Benussi, Anne Boland, Barbara Borroni, María J. Bullido, Paolo Caffarra, Jordi Clarimon, Antonio Daniele, Daniel Darling, Stéphanie Debette, Jean-François Deleuze, Martin Dichgans, Carole Dufouil, Emmanuel During, Emrah Düzel, Daniela Galimberti, Guillermo Garcia-Ribas, José María García-Alberca, Pablo García-González, Vilmantas Giedraitis, Oliver Goldhardt, Caroline Graff, Edna Grünblatt, Olivier Hanon, Lucrezia Hausner, Stefanie Heilmann-Heimbach, Henne Holstege, Jakub Hort, Yoo Jin Jung, Deckert Jürgen, Silke Kern, Teemu Kuulasmaa, Ling Ling, Carlo Masullo, Patrizia Mecocci, Shima Mehrabian, Alexandre de Mendonça, Mercè Boada, Pablo Mir, Susanne Moebus, Fermin Moreno, Benedetta Nacmias, Gael Nicolas, Børge G. Nordestgaard, Goran Papenberg, Janne Papma, Lucilla Parnetti, Florence Pasquier, Pau Pastor, Oliver Peters, Yolande A.L. Pijnenburg, Gerard Piñol-Ripoll, Julius Popp, Laura Molina Porcel, Raquel Puerta, Jordi Pérez-Tur, Innocenzo Rainero, Inez Ramakers, Luis M Real, Steffi Riedel-Heller, Eloy Rodriguez-Rodriguez, Jose Luís Royo, Dan Rujescu, Nikolaos Scarmeas, Philip Scheltens, Norbert Scherbaum, Anja Schneider, Davide Seripa, Ingmar Skoog, Vincenzo Solfrizzi, Gianfranco Spalletta, Alessio Squassina, John van Swieten, Raquel Sánchez-Valle, Eng-King Tan, Thomas Tegos, Charlotte Teunissen, Jesper Qvist Thomassen, Lucio Tremolizzo, Martin Vyhnalek, Frans Verhey, Margda Waern, Jens Wiltfang, Jing Zhang, Henrik Zetterberg, Kaj Blennow, Julie Williams, Philippe Amouyel, Frank Jessen, Patrick G. Kehoe, Ole Andreassen, Cornelia Van Duin, Magda Tsolaki, Pascual Sánchez-Juan, Ruth Frikke-Schmidt, Kristel Sleegers, Tatsushi Toda, Anna Zettergren, Martin Ingelsson, EABD contributors, GR@ACE study group, DEGESCO consortium, DemGene, EADI, GERAD, Asian Parkinson’s Disease Genetics consortium, Yukinori Okada, Giacomina Rossi, Mikko Hiltunen, Jungsoo Gim, Kouichi Ozaki, Rebecca Sims, Jia Nee Foo, Wiesje van der Flier, Takeshi Ikeuchi, Alfredo Ramirez, Ignacio Mata, Agustín Ruiz, Ziv Gan-Or, Jean-Charles Lambert, Michael D. Greicius, Emmanuel Mignot

## Abstract

Using genome-wide association data, we analyzed Human Leukocyte Antigen (HLA) associations in over 176,000 individuals with Parkinson’s (PD) or Alzheimer’s (AD) disease versus controls across ancestry groups. A shared genetic association was observed across diseases at rs601945 (PD: odds ratio (OR)=0.84; 95% confidence interval, [0.80; 0.88]; p=2.2×10^−13^; AD: OR=0.91[0.89; 0.93]; p=1.8×10^−22^), and with a protective HLA association recently reported in amyotrophic lateral sclerosis (ALS). Hierarchical protective effects of *HLA-DRB1*\*04 subtypes best accounted for the association, strongest with *HLA-DRB1*\*04:04 and *HLA-DRB1*\*04:07, intermediary with *HLA-DRB1*\*04:01 and *HLA-DRB1*\*04:03, and absent for *HLA-DRB1*\*04:05. The same signal was associated with decreased neurofibrillary tangles (but not neuritic plaque density) in postmortem brains and was more strongly associated with Tau levels than Aβ42 levels in the cerebrospinal fluid. Finally, protective *HLA-DRB1*\*04 subtypes strongly bound the aggregation-prone Tau PHF6 sequence, but only when acetylated at K311, a modification central to aggregation. An *HLA-DRB1*\*04-mediated adaptive immune response, potentially against Tau, decreases PD, AD and ALS risk, offering the possibility of new therapeutic avenues.

---

Alzheimer’s disease (AD), Parkinson’s disease (PD), Amyotrophic Lateral Sclerosis (ALS) and other neurodegenerative diseases are responsible for considerable morbidity and mortality. With incidence rising with aging, these also represent a growing societal challenge. Pathophysiology involves accumulation of Tau (neurofibrillary tangles) and Amyloid-β-rich (amyloid plaques) aggregates in AD, α-synuclein-rich aggregates (Lewy bodies) in PD and TDP-43 aggregates in ALS, although co-presence of these aggregates may occur. Other diseases such as some Frontotemporal Dementias (FTD) and Progressive Supranuclear Palsy (PSP) also involve Tau aggregates. Consensus is also growing that Tau may also play a key role in PD^1,2^ and ALS^3,4^. Here we show that Human Leukocyte Antigen (HLA) DRB1*04 protects against AD, PD and ALS, three prototypical neurodegenerative diseases, and that *HLA-DRB1*04* selectively binds the K311-acetylated epitope of the PHF6 sequence of the microtubule-associated protein Tau. Presence of DRB1*04 was also associated with lower CSF Tau and less neurofibrillary tangles in AD subjects. As K311-acetylated PHF6 is known to seed Tau aggregation, we propose that improved immune clearance of Tau occurs in DRB1*04 subjects, slowing or reducing neurodegeneration. Targeting this epitope through vaccination or immunotherapy may have preventive or therapeutic potential in multiple neurodegenerative diseases.

Innate immune responses and microglial involvement have long been implicated in neurodegenerative diseases^5–7^. More recently, a role for adaptive immunity in PD, AD and other neurodegenerative diseases has also been outlined through genetic^8–11^ and immunological studies^12–16^. A cornerstone of the adaptive immune response is the highly polymorphic HLA system (also called the Major Histocompatibility Complex, MHC) located on human chromosome 6. *HLA* genes encode a set of proteins that bind peptides derived from foreign or self-antigens, allowing recognition by T cells and subsequent coordination of immune responses. In this context, PD, AD and ALS have genome-wide association study (GWAS) associations in the HLA class II region, within a region containing HLA DR and DQ, two tightly linked set of genes. These genes form α*/*β heterodimers composed of DRA with either *HLA-DRB1, HLA-DRB3, HLA-DRB4* or *HLA-DRB5* gene products for DR molecules, and of *HLA-DQA1* and *HLA-DQB1* gene products for DQ molecules. *HLA-DRA* is almost invariant, while all other genes are highly polymorphic, notably in exon 2, a region that projects residues within a so-called HLA binding groove that functionally binds a 9-residue sequence using molecular pockets that host the P1, P4, P6 and P9 residues (HLA binding register) of the peptide. For the remaining P2, P3, P5, P7 and P8 residues, the amino acid side chain is presented to CD4^+^ T cells, which in turn orchestrate an immune response through T cell receptor (TCR) activation. Additional amino acids flanking the 9-mer core can also modulate HLA binding^17^, although weakly. Depending on one’s specific HLA, individuals present distinct repertoires of bound peptides to CD4^+^ T cells, so that the HLA system is how the immune system sees and reacts to foreign and self-antigens in different individuals.

Although initially attributed to *HLA-DRB1*\*15:01, recent GWAS studies have shown that the HLA signal in PD marks *HLA-DRB1*\*04^18–20^, while in AD and ALS, the signal remains uncharacterized^8,9,11,21^ or also assigned to *HLA-DRB1*\*15:01^9,22^. Additionally, a comparison of healthy controls and individuals with neurodegenerative disorders showed that a complex polyclonal T cell response develops against α-synuclein, Tau, and TDP-43^12–14,23^, proteins involved in pathological aggregates. Whether or not these responses are epiphenomenal, contribute to, or protect against neurodegeneration is unknown, although T cell responses to α-synuclein may occur early in PD^13,14^. Similarly, polyclonal autoantibodies targeting Tau or α-synuclein are detectable in both AD/PD patients and controls, without clear evidence of pathological involvement^15,16^.

To better understand the involvement of HLA in neurodegenerative diseases, we gathered GWAS data available in PD and AD, refining the signal to the HLA subtype level through HIBAG imputation^24^ and fine mapping studies. Because HLA signals are best dissected across ancestry groups, as first shown in narcolepsy with HLA-DQ^25^, we included Europeans, Asians^26–28^, Latinos^29^ and African Americans^30,31^ (**Supplementary Table 1** for samples involved). Here we show that subtypes of *HLA-DRB1*\*04 protect against both PD and AD potentially by presenting acetylated epitopes of PHF6 sequences of the microtubule-associated protein Tau to CD4^+^ T cells. As these epitopes are known to seed Tau aggregation, we propose that improved immune clearance of Tau ensues, reducing pathology. Targeting this epitope through vaccination or immunotherapy may have preventive or therapeutic potential.

## Colocalization of HLA SNP signals in AD and PD across ethnic groups

Two recent large studies in PD^18,19^ have shown that the HLA signal in PD is best summarized by a primary protective effect of *HLA-DRB1* amino acids 13H and 33H shared by all *HLA-DRB1*\*04 subtypes. *HLA-DRB1* amino acids 11V also showed a strong association; this amino acid is shared by *HLA-DRB1*\*04 and *HLA-DRB1*\*10:01, a subtype not associated with the disease. To extend on this, we meta-analyzed samples from North and South America, Europe and East Asia, thereby expanding the sample size to a total of 55,554 PD and PD-proxy cases and 1,454,443 controls (**Supplementary Table 1**). Asian samples were added because of the differential *HLA-DRB1*\*04 subtype distribution in Asians versus Europeans (key haplotypes distribution by ancestry, **Extended Data Figure 1**). The regional HLA association plot confirmed a primary effect driven by rs504594 (odds ratio (OR) = 0.84; 95% confidence interval [CI] [0.80; 0.88]; p=1.83×10^−13^) in PD (locuszoom, **Extended Data Figure 2**), with high association with *HLA-DRB1*\*04-specific amino acid 13H/33H across all ancestry groups (lead variant linkage disequilibrium with various *HLA-DRB1*\*04 subtypes and key amino acids across ancestry groups, **Extended Data Figure 3**) and disappearance of the signal after rs504594 conditioning^19^. This confirmed our initial findings in a larger and more diverse multi-ancestry sample. In contrast to PD, the HLA signal present in AD has not been finely characterized^8,9,21^. To further examine this, we analyzed the region across diverse ancestry groups in 121,371 AD and proxy-AD cases and 410,989 controls from North America, Europe, Africa, and East Asia (**Supplementary Table 1** for description of subsamples). As shown in **Extended Data Figure 2**, the signal peaked at rs35472547 (OR = 0.91; 95% CI [0.89; 0.93]; p= 9.7×10^−23^). The AD and PD signals colocalize in Europeans and across ancestry groups, with rs601945 as the best candidate representative of both signals (**Figure 1**, posterior probability of colocalization, PP4 = 99.5%).

**Figure 1.**
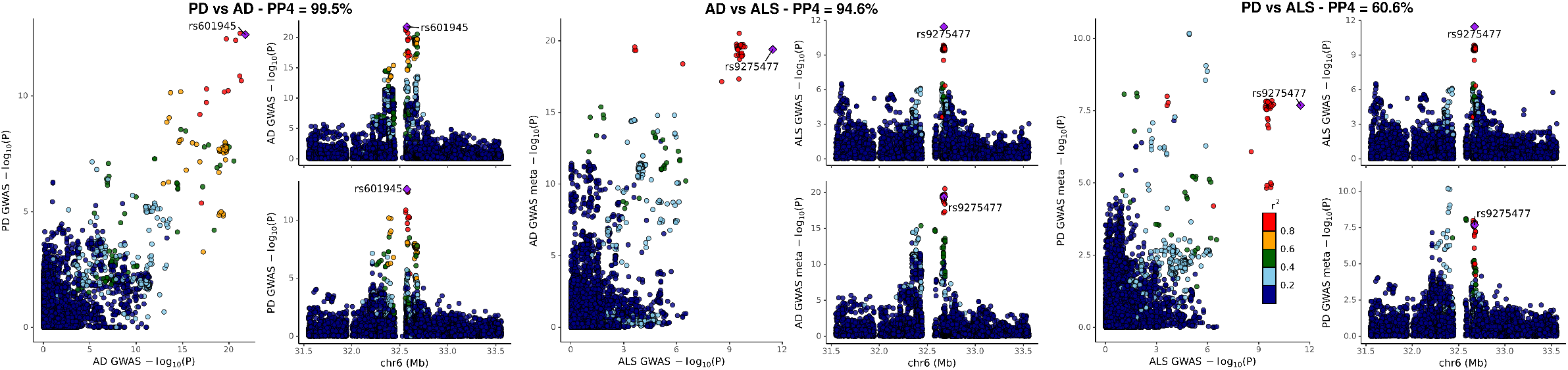
Colocalization of the HLA locus signal in Alzheimer’s disease, Parkinson’s disease and Amyotrophic Lateral Sclerosis.

## Fine mapping studies implicate protective effect of specific *HLA-DRB1*\*04 subtypes in AD and PD

We next conducted HLA imputation and HLA-DR∼DQ analysis in both diseases separately and jointly, as no heterogeneity was found (**Table 1**). As reported in PD^18,19^, *HLA-DRB1* 13H or 33H (two *HLA-DRB1*\*04 amino acids in complete linkage disequilibrium, **Extended Data Figure 3**) gave the highest association signal in both diseases. *HLA-DQB1*\*03:02, an allele highly linked with multiple *HLA-DRB1*\*04 subtypes (**Extended Data Figure 1**), was also associated, but less strongly (**Table 1**), suggesting a primary effect of *HLA-DRB1*. Further, various *HLA-DRB1*\*04 subtypes had hierarchical effects on susceptibility. Specifically, *HLA-DRB1*\*04:04 and *HLA-DRB1*\*04:07 conferred the strongest protection, followed by weaker effects of *HLA-DRB1*\*04:01 and *HLA-DRB1*\*04:03, and no effect for *HLA-DRB1*\*04:05 and *HLA-DRB1*\*04:06 (**Table 1**). The absence of association was notable for *HLA-DRB1*\*04:05, which is a frequent *HLA-DRB1*\*04 subtype in Asians. Although *HLA-DRB1*\*04:05 shares amino acids 13H or 33H, it is distinct from common European *HLA-DRB1*\*04:01, *HLA-DRB1*\*04:02, *HLA-DRB1*\*04:04 and Latino *HLA-DRB1*\*04:07 subtypes because of a D to S substitution at position 57. An additional weaker predisposing effect of the HLA haplotype *DRB1*\*01:01∼*DQA1*\*01:01∼*DQB1*\*05:01 was also observed in AD and was nominally significant in PD with the same direction of effect (**Table 1**). No other genome wide significant effect was observed (**Supplementary Tables 2 and 3**), notably with *HLA-DRB1*\*07:01 or *HLA-DRB1*\*09:01, subtypes which are linked with accessory allele *HLA-DRB4*\*01, also present on *HLA-DRB1*\*04 haplotypes. Of note, *HLA-DRB1* is more expressed than other DRBs, DQ and DP, and is generally the immunodominant response of CD4 cells^32,33^. Further, accessory *HLA-DRB*3/4/5 genes also commonly share a binding repertoire with primary *HLA-DRB1* genes and are typically minor components of HLA Class II immune responses^32,33^.

**Table 1.** *HLA-DRB1* alleles *HLA-DRB1*\*04:04 and *HLA-DRB1*\*04:01 are associated with a decreased risk of Parkinson’s and Alzheimer’s diseases. Effect sizes are reported as odds ratio (OR), with 95% confidence interval [CI], and significance (p-value). FreqC: frequency of carriers, N: number of individuals analyzed for a given allele/haplotype/amino acid, p-het: heterogeneity p-value.

|  | HLA | Parkinson's Disease (PD) |  |  |  | Alzheimer's Disease (AD) |  |  |  | PD + AD |  |  |
| --- | --- | --- | --- | --- | --- | --- | --- | --- | --- | --- | --- | --- |
|  |  | FreqC | N | OR | pval | FreqC | N | OR | pval | OR | pval | p-het |
| HLA Alleles | *HLA-DRB1* *04:01 | 0.196 | 1484656 | 0.92[0.89; 0.95] | 2.4E-08 | 0.191 | 487237 | 0.93[0.9; 0.96] | 6.5E-07 | 0.92[0.91; 0.94] | 9.1E-14 | 0.56 |
|  | *HLA-DRB1* *04:02 | 0.019 | 1474730 | 0.92[0.85; 0.99] | 0.02 | 0.022 | 155846 | 1.00[0.91; 1.10] | 0.99 | 0.95[0.89; 1.01] | 0.07 | 0.17 |
|  | *HLA-DRB1* *04:03 | 0.012 | 980868 | 0.89[0.81; 0.97] | 0.01 | 0.071 | 8346 | 1.11[0.93; 1.31] | 0.25 | 0.93[0.86; 1.01] | 0.1 | 0.03 |
|  | *HLA-DRB1* *04:04 | 0.074 | 1475574 | 0.84[0.80; 0.88] | 1.5E-11 | 0.073 | 476236 | 0.86[0.82; 0.90] | 8.9E-12 | 0.85[0.82; 0.88] | 9.3E-22 | 0.60 |
|  | *HLA-DRB1* *04:05 | 0.013 | 1507057 | 1.00[0.95; 1.05] | 0.86 | 0.027 | 169839 | 0.99[0.92; 1.07] | 0.77 | 0.99[0.95; 1.04] | 0.76 | 0.88 |
|  | *HLA-DRB1* *04:06 | 0.046 | 32327 | 0.95[0.82; 1.09] | 0.46 | 0.059 | 8346 | 1.00[0.83; 1.20] | 0.99 | 0.97[0.86; 1.08] | 0.55 | 0.66 |
|  | *HLA-DRB1* *04:07 | 0.021 | 526189 | 0.79[0.69; 0.91] | 7.3E-04 | 0.019 | 474840 | 0.88[0.81; 0.96] | 4.5E-03 | 0.86[0.79; 0.92] | 2.7E-05 | 0.18 |
|  | *HLA-DRB1* *04:10 | 0.041 | 4853 | 0.91[0.67; 1.25] | 0.57 | 0.032 | 8744 | 1.23[0.96; 1.58] | 0.10 | 1.09[0.90; 1.33] | 0.36 | 0.14 |
|  | *HLA-DRB1* *01:01 | 0.181 | 1485033 | 1.05[1.01; 1.09] | 7.0E-03 | 0.186 | 487879 | 1.07[1.04; 1.10] | 4.6E-07 | 1.06[1.04; 1.09] | 1.4E-08 | 0.45 |
|  | *HLA-DQB1* *03:02 | 0.191 | 1501065 | 0.91[0.88; 0.93] | 2.6E-14 | 0.190 | 510906 | 0.89[0.87; 0.91] | 4.3E-19 | 0.9[0.88; 0.91] | 1.5E-31 | 0.27 |
| HLA Haplotypes | DRB1*04:01~DQA1*03:01~DQB1*03:02 | 0.088 | 1473386 | 0.95[0.92; 0.98] | 3.6E-03 | 0.097 | 521560 | 0.91[0.87; 0.95] | 6.5E-06 | 0.93[0.91; 0.96] | 2.1E-07 | 0.11 |
|  | DRB1*04:01~DQA1*03:03~DQB1*03:01 | 0.12 | 1481518 | 0.98[0.97; 0.99] | 5.2E-04 | 0.146 | 532448 | 0.98[0.94; 1.02] | 0.24 | 0.98[0.97; 0.99] | 2.4E-04 | 0.66 |
|  | DRB1*04:04~DQA1*03:01~DQB1*03:02 | 0.077 | 1475734 | 0.84[0.80; 0.89] | 1.7E-10 | 0.092 | 524205 | 0.85[0.81; 0.89] | 5.4E-11 | 0.85[0.82; 0.88] | 3.8E-20 | 0.80 |
|  | DRB1*04:07~DQA1*03:01~DQB1*03:02 | 0.198 | 1498 | 0.58[0.44; 0.76] | 6.1E-05 | 0.063 | 1865 | 0.75[0.47; 1.21] | 0.23 | 0.67[0.53; 0.83] | 3.7E-04 | 0.36 |
|  | DRB1*04:07~DQA1*03:03~DQB1*03:01 | 0.021 | 524845 | 0.87[0.73; 1.04] | 0.12 | 0.019 | 519510 | 0.87[0.79; 0.97] | 8.0E-03 | 0.87[0.8; 0.95] | 2.1E-03 | 0.93 |
|  | DRB1*04:05~DQA1*03:03~DQB1*04:01 | 0.11 | 35369 | 0.99[0.92; 1.06] | 0.76 | 0.227 | 10995 | 1.08[0.98; 1.18] | 0.11 | 1.02[0.97; 1.08] | 0.46 | 0.15 |
|  | DRB1*08:02~DQA1*03:01~DQB1*03:02 | 0.026 | 5602 | 1.10[0.76; 1.60] | 0.61 | 0.026 | 10995 | 0.94[0.74; 1.19] | 0.59 | 0.98[0.8; 1.19] | 0.84 | 0.47 |
|  | DRB1*01:01~DQA1*01:01~DQB1*05:01 | 0.150 | 1485406 | 1.05[1.01; 1.09] | 0.02 | 0.233 | 537567 | 1.08[1.05; 1.11] | 3.4E-07 | 1.07[1.04; 1.09] | 3.6E-08 | 0.31 |
| Amino-acid: *HLA-DRB1* 13H/ 33H |  | 0.300 | 1507057 | 0.89[0.87; 0.91] | 9.3E-22 | 0.315 | 527932 | 0.91[0.89; 0.93] | 5.9E-18 | 0.90[0.89; 0.92] | 7.6E-38 | 0.31 |

## Tau-pathology is reduced in AD individuals with *HLA-DRB1*\*04 subtypes

How could an *HLA-DRB1*\*04 subtype-specific association be involved in AD? To investigate this question, we first used neuropathological information of 7,259 postmortem samples of European ancestry available through the Religious Orders Study and Memory and Aging Project^34^ and the National Institute on Aging - Alzheimer’s Disease Center cohorts 1 to 7^35^, looking at the effect of rs601945 on Tau Braak staging and neuritic plaques density in US-based pathological samples. As shown in **Table 2**, a strong association of rs601945, and by extension, *HLA-DRB1*\*04 13H and *HLA-DRB1*\*04 alleles (**Supplementary Table 4**) with decreased neurofibrillary tangles, but not amyloid plaques, was observed, suggesting the involvement of Tau. The analysis of cerebrospinal fluid (CSF) Aβ42 and Tau levels in 8,074 subjects of European ancestry independently confirmed this observation. In CSF, rs601945 and *HLA-DRB1*\*04:04 were significantly associated with significant lower levels of phosphorylated- and total-Tau (**Table 2**), but only weakly associated with increased Aβ42 levels. Interestingly, *HLA-DRB1*\*04 13H was also associated with an older age of onset in AD (**Supplementary Table 4**).

**Table 2.**
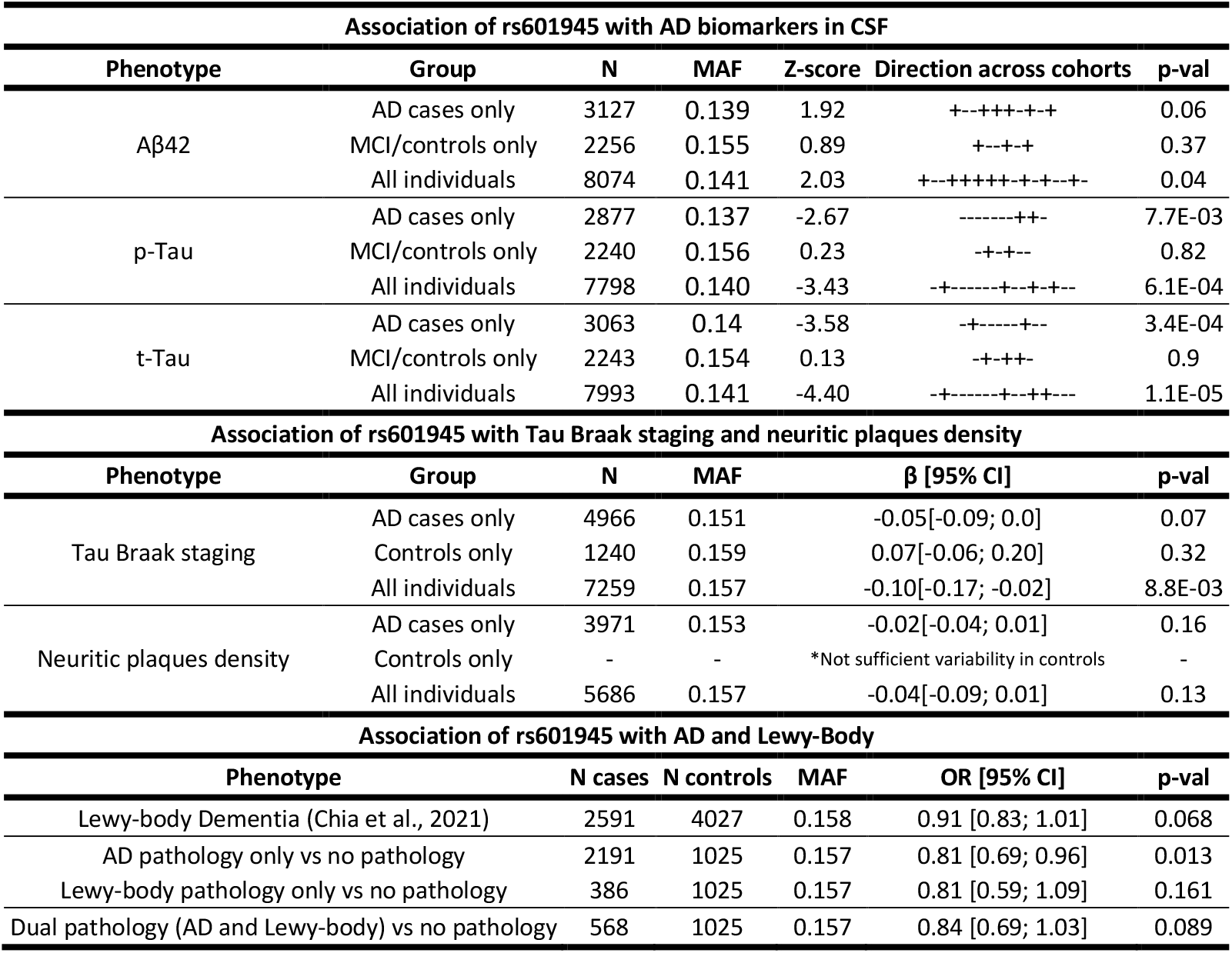
rs601945 is associated with reduced Tau in CSF and postmortem brain tissues, with protection against Lewy-body pathology, but association with increased Aβ42 and decreased neuritic plaques density is weak. p-Tau: phosphorylated Tau, t-Tau: total Tau, N: number of individuals, MAF: minor allele frequency, OR: odds ratio, β: parameter estimate, CI: confidence interval. Braak: Tau Braak staging, Neur: Neuritic plaques density.

## The same HLA effect is associated with decreased risk in other neurodegenerative diseases

The HLA locus association with AD and PD was found to colocalize with the protective HLA signal in ALS^11^, with PP4 = 94.6%, and PP4 = 60.6%, respectively (**Figure 1**). PP4 between PD and ALS is below the classic 80% level to confirm colocalization, which is likely due to the absence of lead variants from our PD GWAS in the ALS GWAS summary statistics^11^ (**Extended Data Figure 2**), We hypothesize that improved quality control and imputation on the TOPMed reference panel would strengthen colocalization. It’s worth noting that using summary statistics from a smaller PD GWAS in Europeans, van Rheenen et al.^11^ reported PP4 = 80.4% for PD vs. ALS.

In addition, studying a subset of autopsied individuals with dementia and with either Lewy Body, AD pathology or both pathologies, rs601945 shows a nominally significant protective association in the AD only pathology group and concordant protective effects in the Lewy Body only pathology group and in the dual pathology group (though smaller sample sizes here resulted in non-significant results, **Table 2**). This protective association was also observed in the largest GWAS to date of Lewy body dementia that included a mix of clinically diagnosed and pathologically confirmed cases (OR=0.91 [0.83; 1.01], p = 0.07). Taken together, these results indicate that the HLA locus may be involved in a common immune response across these neurodegenerative diseases.

## Immune response against post-translationally modified proteins

Based on these results, we hypothesized that a *HLA-DRB1*\*04-restricted adaptive immune response directed against Tau may be protective in AD and other neurodegenerative diseases. However, how could *only HLA-DRB1*\*04-restricted responses show a protective effect? Tau, like other proteins involved in neurodegeneration, is highly post-translationally-modified (PTM) through phosphorylation and acetylation, phenomena that likely predispose Tau to aggregation. In autoimmune diseases, PTMs (such as citrullination of fibrinogen for rheumatoid arthritis) are frequently found in culprit autoantigens. Further, PTMs contribute to reduced self-tolerance as these are not presented in the thymus for negative selection^36^. This likely explains the strong polyclonal T cell response observed in controls and AD/PD patients against Tau, β-amyloid and α-synuclein^12,37,38^. A similarly broad B cell response against Tau and α-synuclein is also reported in controls and patients^15,16^. As mentioned above, prior work has outlined strong polyclonal CD4^+^ T cell responses against α-synuclein, β-amyloid and Tau peptides when presented by various HLA subtypes in both cases and controls, thus it is unclear if these responses are effective in limiting disease or are a simple bystander effect^12^. One recent publication also showed the presence of CD4^+^ and CD8^+^ T cells in the CSF of PD and AD patients^39^, suggesting CD8^+^ T cell-mediated clearing of amyloid plaques.

Considering that CD4^+^ T cell and B cell responses against Tau are common and robust, we hypothesized that a specific *HLA-DRB1*\*04 restricted response, unlike other responses, may target a particular Tau epitope potentially important for aggregation. To test this hypothesis, we screened the most frequent PTM and non-PTM modified peptides for *HLA-DRB1*\*04 subtype-specific binding, using the highly protective *HLA-DRB1*\*04:04 subtype, the moderately protective *HLA-DRB1*\*04:01 subtype and the neutral *HLA-DRB1*\*04:05 subtype (**Figure 2, Supplementary Table 5**). Because PTMs are more likely to escape central tolerance^36^ and thus would be best candidate, tau PTMs that are common and/or have been involved in the pathophysiology of AD^40^ were included. As a control, alpha-synuclein, another commonly PTM-modified protein involved in PD was also tested **(Extended Data Figure 4, Supplementary Table 6)**.

**Figure 2.**
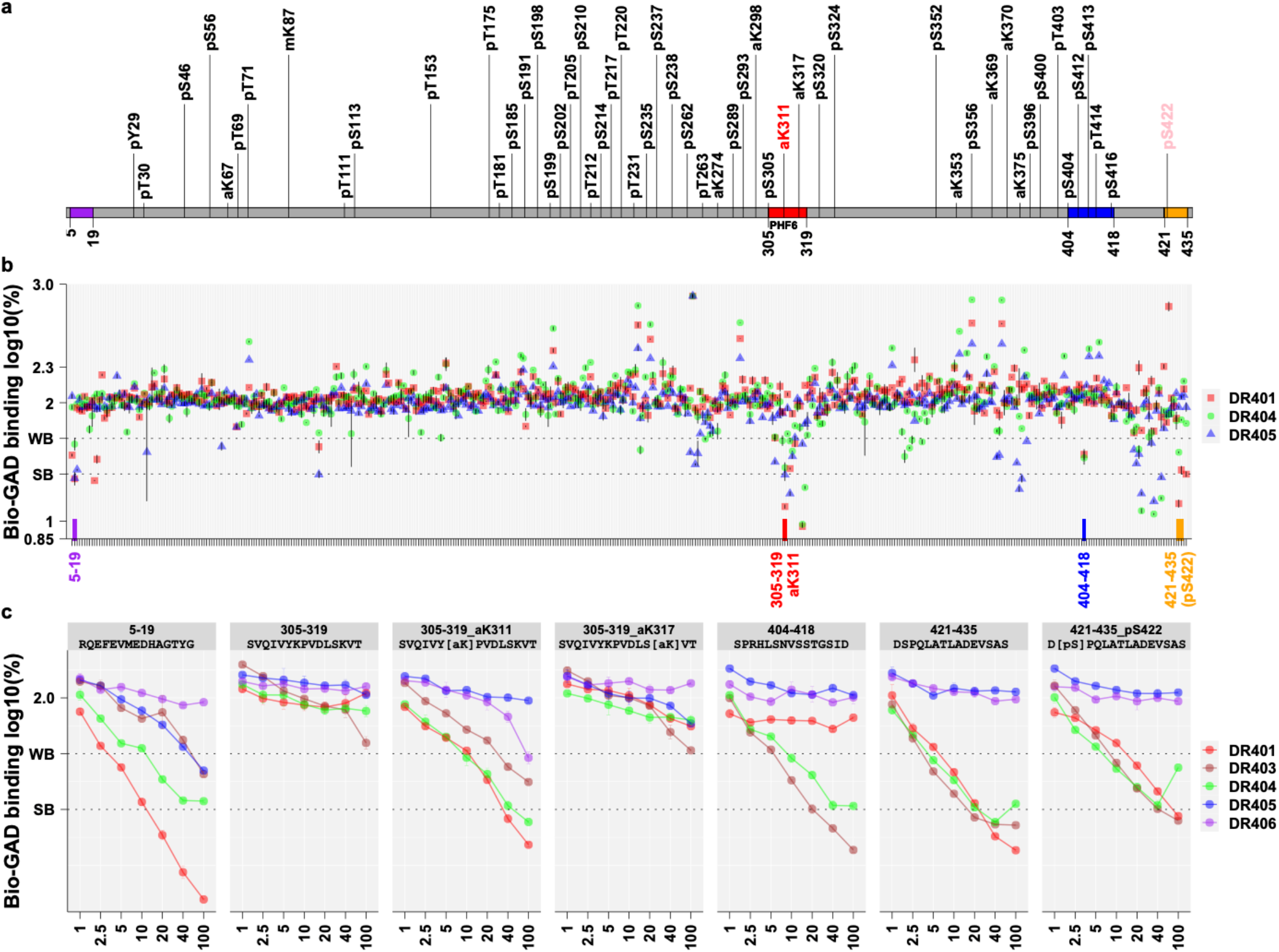
The pro-aggregation PHF6 region of Tau binds *HLA-DRB1*\*04 subtypes only when acetylated at K311. Fifteen-mer peptides (800 µM) encompassing the entirety of all Tau isoforms (schematized in the top panel), overlapping across 11 residues, were screened for HLA-DRB1*04:01, HLA-DRB1*04:04 and HLA-DRB1*04:05 binding (see methods), with and without common PTMs as reported by Wesseling et al^40^. Four regions (labelled in purple, red, blue, orange) containing strong HLA-DRB1*04 binders (log displacement <1-1.4, below 25% of baseline control) were further tested at various concentrations (bottom panel), showing three promising regions where binding was stronger with HLA-DRB1*04:04/ HLA-DRB1*04:01, intermediary with HLA-DRB1*04:03 and absent or weak with HLA-DRB1*04:05 and HLA-DRB1*04:06, a pattern similar to GWAS risk (Table 1). Among these regions, PHF6 ^306^VQ<u>IVY(acetylK)PVDLS</u>K^317^ that strongly binds HLA-DRB1*04:01, HLA-DRB1*04:03 and HLA-DRB1*04:04 only when post-translationally modified at K311. This segment is well known to seed Tau aggregation, and this process is increased in the presence of acetylK311.

## DRB1*04 preferentially binds acetylated PHF6, a sequence known to be important in aggregation seeding

Among over a thousand peptides tested, only a few peptides strongly bound *HLA-DRB1*\*04, with PHF6 in Tau (R3/wt; ^306^VQ<u>IVY(acetylK)PVDLS</u>KV^318^) standing as the ideal candidate (**Figure 2**). The PHF6 sequence is located within the Microtubule Binding Area of Tau, known to be important for aggregation^41^. PHF6 creates core nucleation sites that can, by themselves, induce formation of fibrils displaying the paired helical filament (PHF) morphology characteristic of neurofibrillary tangles^41,42^. Most interestingly, PHF6 sequences only bind *HLA-DRB1*\*04 when K311 is acetylated. Further, acetylated sequences have significantly less affinity for *HLA-DRB1*\*04:05 versus other subtypes (*HLA-DRB1*\*04:04= *HLA-DRB1*\*04:01> *HLA-DRB1*\*04:03> *HLA-DRB1*\*04:06> *HLA-DRB1*\*04:05), the same hierarchy observed in the case/control association results, with cores predicted to strongly bind *HLA-DRB1*\*04:04 (**Figure 2, Extended Data Figures 5 and 6**)^43,44^. This is most likely because serine in molecular pocket P9 is known to reduce binding to *HLA-DRB1*\*04:05 in comparison to other *HLA-DRB1*04* subtypes^43,44^ (**Extended Data Figure 5**). Additional PTMs in the area, such as acetylated K317, do not alter binding, although the presence of phosphoserine at S305, another frequent PTM, slightly reduces PHF6 binding (**Figure 2**). Other peptides were found to bind *HLA-DRB1*04* in both Tau (**Figure 2**) and α-synuclein (**Extended Data Figure 4**), but in no other case a PTM was necessary for epitope binding. A strong predicted binding for both acetylated and unacetylated PHF6 motifs was also observed with accessory gene *HLA-DRB4*\*01 (**Extended Data Figure 6**), but the absence of association with *HLA-DRB1*\*07:01 and *HLA-DRB1*\*09:01, whose haplotypes are in linkage with *HLA-DRB4*\*01, exclude its involvement (**Supplementary Table 7**).

## Discussion

The fact that *HLA-DRB1*\*04 only binds *acetylated* forms of PHF6 strongly favors involvement of this epitope in the protective effect of *HLA-DRB1*\*04 in AD. With K317 located nearby, the K311 PTM of PHF6 is the most differentiating Tau PTM found in AD versus control brains^44^. Further, K311 acetylation has been shown by multiple investigators to promote aggregation of PHF6 *in vivo*^45^, *in vitro*^46^ and *in silico*^47^, while K311 carbamylation is inhibitory^48^. Crystalography studies have also shown that acetylated PHF6 dominates in the formation of long fibrils as in neurofibrillary tangles of AD^49,50^. Finally, *HLA-DRB1*\*04-associated subtypes are the *only* frequent HLA-DR and DQ subtypes with predicted high affinity for this epitope **(Extended Data Figure 6)**, likely explaining why *HLA-DRB1*\*04, and no other subtypes, mediate this effect. Acetylation at the K311 Tau residue may be mediated by SIRT1 and/or HDAC6, current therapeutic targets in AD^45,51^. Similarly, recent evidence suggests that reducing acetylated Tau is neuroprotective in brain injury^52^.

K311 is not only acetylated, but also ubiquitinated^53,54^, or succinylated^55^, and the epitope trafficked to the NLRP3 inflammasome of microglial cells^56^,where HLA class II presentation of Tau fragments by *HLA-DRB1*\*04 to T cells is also likely to occur^57^. Involvement of microglial cells is also suggested by various GWAS association signals observed in AD^5^ and PD^6^. Although the ubiquitinated K311 epitope is unlikely to bind *HLA-DRB1*\*04 due to steric hinderance at P4, ubiquitination at K317 at P10 could further modulate *HLA-DRB1*\*04 binding and the effect of K311 succinylation on DRB1*04 binding is unknown. Additional experiments exploring *HLA-DRB1*\*04 subtype binding of PTM segments of PHF6 in various combinations will be needed to further this line of investigation.

Interestingly, besides PHF6, a homologous segment, PHF6* (R2/wt; ^273^GKVQIINKKLDL^284^) has similar pro-aggregation properties in vitro^41,42,46^, with acetylation of K280 in PHF6* also promoting aggregation *in vitro*^50,58^. Involvement of PHF6*, an earlier candidate for aggregation seeding, is however less clear *in vivo*, with acetylation of K280 being a rare PTM^54^. Further, although predicted to also bind *HLA-DRB1*04* with a similar repertoire (with acetylated K280 in position 4) (**Supplementary Figure 1**), acetylated PHF6* sequences did not bind *in vitro* (**Supplementary Figure 2**), illustrating the limitation of *in silico* predictions.

Another less probable candidate mediating the protective effect of *HLA-DRB1*04* is in the C-terminal end of Tau, the ^446^LATLADEVS^455^sequence. This sequence also binds *HLA-DRB1*\*04:04> *HLA-DRB1*\*04:01 > *HLA-DRB1*\*04:05 as expected from our association studies. This region may also be relevant to AD pathogenesis, as C-terminal cleavage of Tau is an early event leading to aggregation^54,59^. This C-terminal epitope is, however, also predicted to bind other frequent *HLA-DRB1* subtypes with similar affinity and core sequences (**Supplementary Figure 3**) and is thus a much less likely candidate.

## Conclusion

Overall, our results indicate that a *HLA-DRB1*\*04-subtype-specific adaptive immune response is protective against both AD and PD, with recent work suggesting the same is true for ALS. Although it is impossible to exclude involvement of other proteins in this effect, a CD4^+^ T cell reactivity toward PHF6 fragments containing the acetylated K311 epitope of Tau is a strong candidate for mediating this effect, based on current knowledge. We hypothesize that this T cell reactivity facilitates early clearing of toxic tau aggregate seeds, for example, by recruiting B cells and generating an antibody response targeting the same epitope, or simply by facilitating T cell clearing of early aggregates (**Figure 3**). Tau’s biology as a regulator of microtubule assembly in neurons is uniquely positioned to potentiate neurotoxicity across multiple diseases involving different aggregates, as reported here.

**Figure 3.**
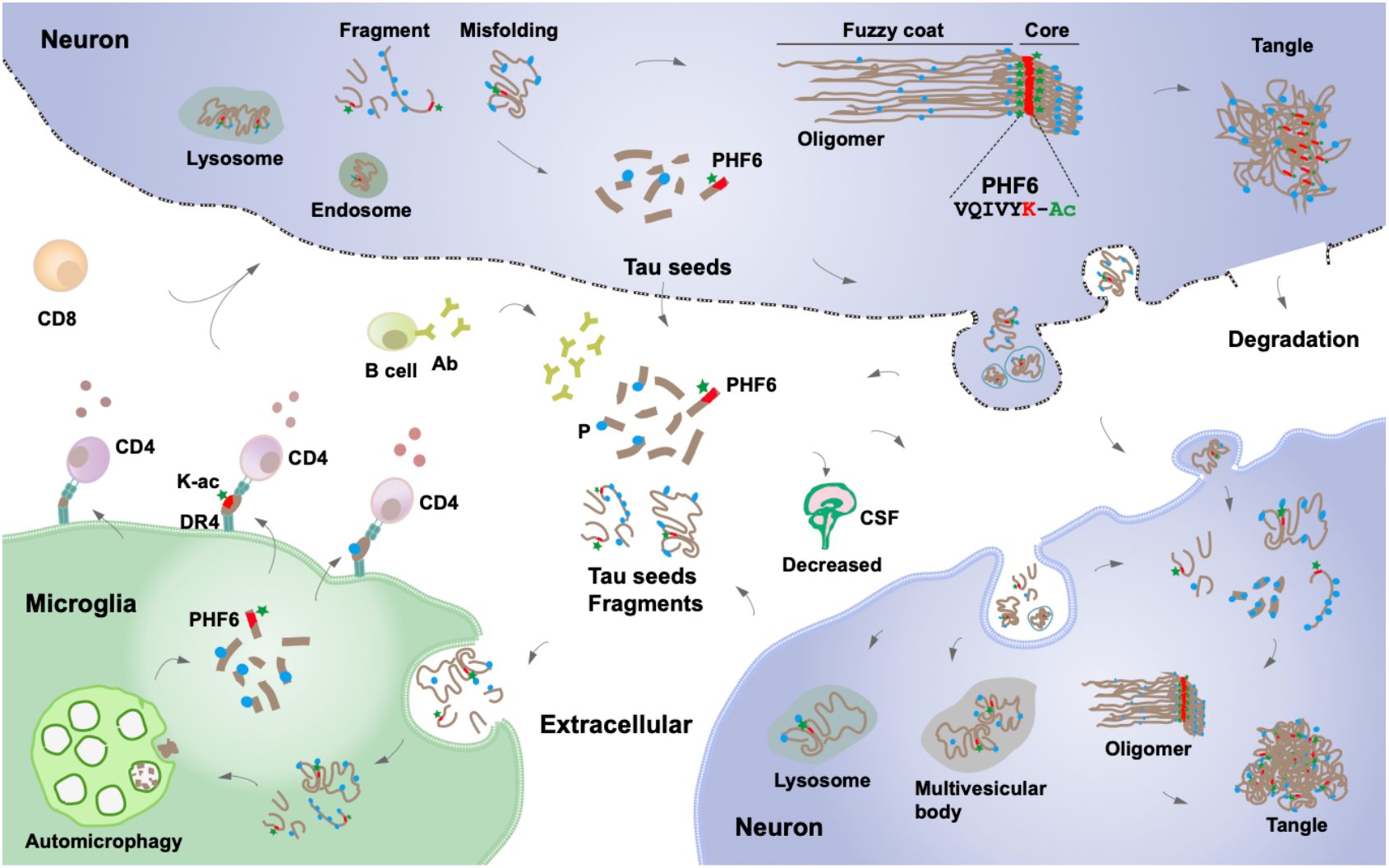
Immune clearance of Acetylated PHF6 Tau sequences reduces neurodegeneration in AD and PD. Pathological Tau seeds, soluble Tau fragments, or misfolded Tau present in the extracellular space are taken up and phagocytosed by activated microglia where it is processed. In addition to autophagy, resulting Tau peptide fragments, notably acetylated lysine (K-ac) 311 PHF6 are bound to HLA-DRB1*04:01 or HLA-DRB1*04:04 and the resulting HLA-peptide complexes presented by microglial cells (or other antigen presenting cells) to CD4^+^ T helper cells. CD4^+^ T cells trigger beneficial downstream immune responses perhaps involving CD8^+^ T and antibody producing B cells. These responses limit propagation of misfolded Tau and reduce neuropathology, also explaining reduced CSF Tau in *HLA-DRB1*\*04 individuals.

Our results also open the possibility that targeting Tau epitopes containing acetyl-K311 through chimeric antigen receptor T cells or antibodies could have therapeutic value. Further, vaccination with acetylated PHF6-like epitopes could reduce disease progression in *HLA-DRB1*\*04 individuals (∼30% of the European population). It is noteworthy that antibodies, although not targeting acetyl-K311 per se, but regions adjacent within PHF6, have been shown to reduce CSF Tau and Tau pathology in animal models^60^ and are being tested as a means of preventing autosomal dominant forms of AD^61^. Autoantibodies against Tau are also present in healthy and demented individuals^15,16^, but epitope specificities have yet to be determined. HLA studies of other tauopathies with different aggregates may also help clarifying how Tau modifications within PHF6 or other aggregation-prone domains modulate susceptibility to other diseases^41,42^. Tau may be an important modulator of neurodegeneration across multiple diseases.

## Data Availability

All data produced in the present work are contained in the manuscript Supplementary Tables.
Data used in preparation of this manuscript can be obtained upon application at:
-     dbGaP (https://www.ncbi.nlm.nih.gov/gap/advanced_search/)
-    NIAGADS and NIAGADS DSS (https://www.niagads.org/)
-    LONI (https://ida.loni.usc.edu/)
-   Synapse (https://adknowledgeportal.synapse.org/)
-    RADC Rush (https://www.radc.rush.edu/)
-    NACC (https://naccdata.org/)
-    UK Biobank (biobank.ndph.ox.ac.uk/showcase/)

https://www.ncbi.nlm.nih.gov/gap/advanced_search/

https://ida.loni.usc.edu/

https://adknowledgeportal.synapse.org/

https://www.radc.rush.edu/

https://naccdata.org/

## METHODS Online

### Samples

Participants or their caregivers provided written informed consent in the original studies. The current study protocol was granted an exemption by the Stanford University institutional review board because the analyses were carried out on deidentified, off-the-shelf data; therefore, additional informed consent was not required.

<u>The AD samples</u> included in the analysis are part of the following datasets with phenotype, genotype ascertainment, and quality control described elsewhere^9,28,30,8,60,61,62,63^: the European Alzheimer’s Disease BioBank (EADB), The Genome Research @ Fundació ACE project (GR@ACE), Genetic and Environmental Risk in AD/Defining Genetic, Polygenic and Environmental Risk for Alzheimer’s Disease Consortium (GERAD), the European Alzheimer’s Disease Initiative (EADI), the Norwegian DemGene (DemGene), the Bonn study (Bonn), the Copenhagen City Heart Study (CCHS), the Alzheimer’s Disease Genetics Consortium (ADGC), the Alzheimer Disease Sequencing Project (ADSP), the UK Biobank, the Gwangju Alzheimer’s and Related Dementias (GARD) study, the Japanese Genetic Study Consortium for Alzheimer’s Disease (JGSCAD) and National Center for Geriatrics and Gerontology (NCGG) both from Niigata University.

<u>The PD samples</u> included in the analysis are part of the following datasets for which the phenotyping, genotyping and quality control has been described elsewhere^10,18,19,26,29,66^: International Parkinson’s Disease Genomics Consortium (IPDGC) NeuroX dataset, McGill University (McGill), National Institute of Neurological Disorders and Stroke (NINDS) Genome-Wide genotyping in Parkinson’s Disease, NeuroGenetics Research Consortium (NGRC), Oslo Parkinson’s Disease Study (Oslo), Parkinson’s Progression Markers Initiative (PPMI), Autopsy-Confirmed Parkinson Disease GWAS Consortium (APDGC), the UK Biobank, East Asians samples from Japan, China, Singapore, Taiwan, and Hong-Kong (EastAsians-PD), 23andMe, and the Latin American Research Consortium on the Genetics of Parkinson’s Disease (LARGE-PD).

<u>The samples assessed for AD and Lewy-body neuropathology</u> included genetic data from the Rush Religious Orders Study and Memory and Aging Project (ROSMAP)^67^ and from the Alzheimer’s Disease Center (ADC) cohorts 1 to 7 parts of the ADGC^68^, and neuropathological assessment followed procedures described respectively in Naito et al.^34^ and in the National Alzheimer’s Coordinating Center (NACC) postmortem evaluation protocol^35^.

<u>The samples with CSF biomarkers</u> included in the analysis for which phenotype, genotype ascertainment, and quality control is described elsewhere^8,69^, are mostly part of the EADB. The remaining of this dataset includes samples originating from the Gothenburg H70 Birth Cohort studies and clinical AD samples from Sweden.

### Genome-wide association at the *HLA* locus and colocalization between AD and PD

Given the known signal at HLA in AD GWAS^8,68^ and PD GWAS^10^. We aimed at refining the signal at the HLA locus using a multi-ancestry meta-analysis design. We considered a region, #1MB around *HLA-DRB1*, on chromosome 6 from base pair positions (hg38) 31578952 to 33589718. For the PD local-GWAS at the HLA locus, we meta-analyzed the summary statistics from the largest available GWAS to date in European ancestry^10^ (distributed without 23andMe), with the Latino-Amerindian GWAS from Kunkle et al.^29^ and the Asian GWAS from Kang et al.^26^. For the AD local-GWAS at the HLA locus, we meta-analyzed the summary statistics the largest available GWAS to date in European ancestry^8^ (which did not include their Stage 2), with the Korean/Japanese GWAS from Satake et al.^65^, the Japanese GWAS from Loesch et al.^28^, and with in-house local-GWAS at the HLA locus on ADSP and ADGC data carried out by ancestry in European, Latino-Amerindian, African individuals, analyzed with a linear-mixed model as implemented in *GENESIS*^70^ (see Statistical analysis section) adjusted for 6 PCs and sex. All meta-analyses were implemented with a fixed-effect inverse variance weighted design implemented in *METAL*^71^. Colocalization between the AD and PD HLA signals in these multi-ancestry meta-analyses was assessed using the Bayesian model implemented in *coloc*^72^ using default priors. We report the posterior probability of colocalization (PP4) between AD and PD associations on **Figure 1**. Given the high linkage disequilibrium (r^2^ > 0.70) of the lead SNP at the HLA locus in the latest ALS GWAS^73^ with the lead SNPs in our local-GWAS at HLA in AD and PD, we also tested the colocalization of these two diseases with ALS using the same method as described above.

### Imputation and statistical analysis of *HLA* alleles, haplotypes, and amino acids

Two-field resolution alleles of *HLA-A, HLA-B, HLA-C* class I genes, and *HLA-DPB1, HLA-DQA1, HLA-DQB1*, and *HLA-DRB1* class II genes were imputed using R package HIBAG v1.22^24^ for the following dataset: EADB, GR@ACE, GERAD, EADI, DemGene, Bonn, CCHS, UK Biobank, IPDGC, NINDS, NGRC, McGill, Oslo, PPMI, APDGC, LARGE-PD, ADSP, ADGC, GARD, NCGG. When available, training sets specific to ancestry (European, East Asian, Latino, African) and genotyping array were used, either available through HIBAG^24^ or trained in-house as previously described^19^.

<u>In the allele-level analyses</u>, alleles with an imputation posterior probability lower than 0.5 were considered as undetermined as recommended by HIBAG developers. Each allele was considered as a variant and analyzed under a dominant model; supplementary methods provide details on the analysis per cohort.

<u>In the haplotype-level analyses</u>, only individuals with non-missing allele genotypes were included in the haplotype level analysis. Three-locus HLA class I or class II haplotypes were determined using the haplo.em function from the R haplo.stats package. Only haplotypes with posterior probability >0.5 and a carrier frequency of >1% were included in the analysis. Each haplotype was considered as a variant and analyzed under a dominant model; supplementary methods provide details on the analysis per cohort.

<u>In the amino-acid-level analyses</u>, HIBAG^24^ was used to convert P-coded alleles to amino acid sequences for exon 2, 3 of HLA class I genes, and exon 2 of class II genes. Each amino acid was considered as a variant and analyzed under a dominant model; supplementary methods provide details on the analysis per cohort.

For the EastAsians-PD and 23andMe cohorts, the HLA alleles, haplotypes, amino acids statistics were derived from GWAS summary statistics data using the DISH software^74^ as described in Naito et al.^18^.

The allele-, haplotype-, amino-acid-level analyses were respectively meta-analyzed separately between the two neurodegenerative diseases, and across diseases, using a fixed-effect inverse variance weighted design implemented in *METAL*^71^.

### Tau peptide binding

Competition binding studies were conducted as previously described^75^. In brief, Tau peptides at different concentrations were incubated with DRB1*04:01, DRB1*04:03, DRB1*04:04, DRB1*04:05, or DRB1*04:06 (from the Emory University NIH core tetramer facility http://tetramer.yerkes.emory.edu/support/faq) for 3 days at 37°C together with biotinylated GAD or EBV (Bio-GAD, EBV). The reaction was quenched by adding neutralization buffer and then transferred into anti-DR antibody precoated a 96-well plate. DELFIA^®^ time-resolved fluorescence (TRF) intensity was detected using a Tecan SPARK after adding Europium (Eu)-labelled streptavidin. Non-specific binding was removed through extensive wash. Each peptide was duplicated. Competitor Tau peptide with Eu TRF intensity that was lower than 25% and 25-50% of Bio-GAD or EBV epitope alone was considered strong binder and weak binder, respectively.

## Funding and acknowledgments

This work was supported by the Michael J. Fox Foundation grant MJFF-020161 (EM, ZGO), National Institute of Health and National Institute of Aging grants AG060747 (MDG), AG066206 (ZH), AG066515 (ZH, MDG), the European Union’s Horizon 2020 research and innovation program under the Marie Skłodowska-Curie (grant agreement No. 890650, YLG), the Alzheimer’s Association (AARF-20-683984, MEB), and the Iqbal Farrukh and Asad Jamal Fund, a grant from the EU Joint Programme – Neurodegenerative Disease Research (European Alzheimer DNA BioBank, EADB; JPND), the Japan Agency for Medical Research and Development (JP21dk0207045, TI), the Einstein Center for Neurosciences in Berlin (SMY). Inserm UMR1167 is also funded by the Inserm, Institut Pasteur de Lille, Lille Métropole Communauté Urbaine, and the French government’s LABEX DISTALZ program (development of innovative strategies for a transdisciplinary approach to Alzheimer’s disease). Additional funders of individual investigators and institutions who contributed to data collection and genotyping are provided in the **Supplementary Materials**.

---

**Extended Data Figure 1.**
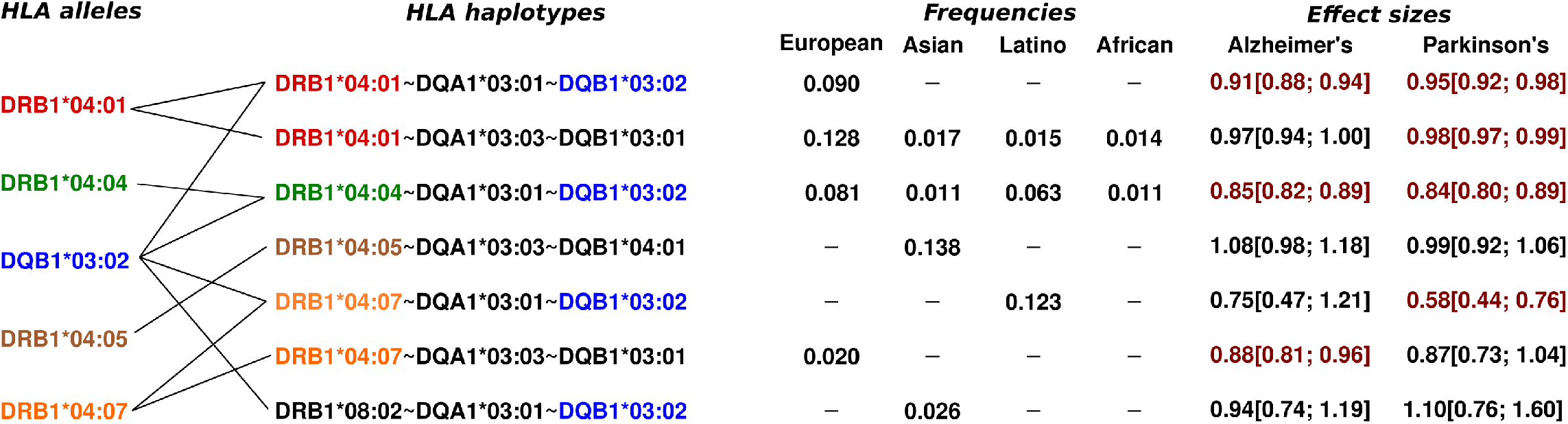
Haplotypes harboring key *HLA-DRB1*\*04 subtypes and/or *HLA-DQB1*\*03:02. Effect sizes highlighted in red were nominally significant (p < 0.05).

**Extended Data Figure 2.**
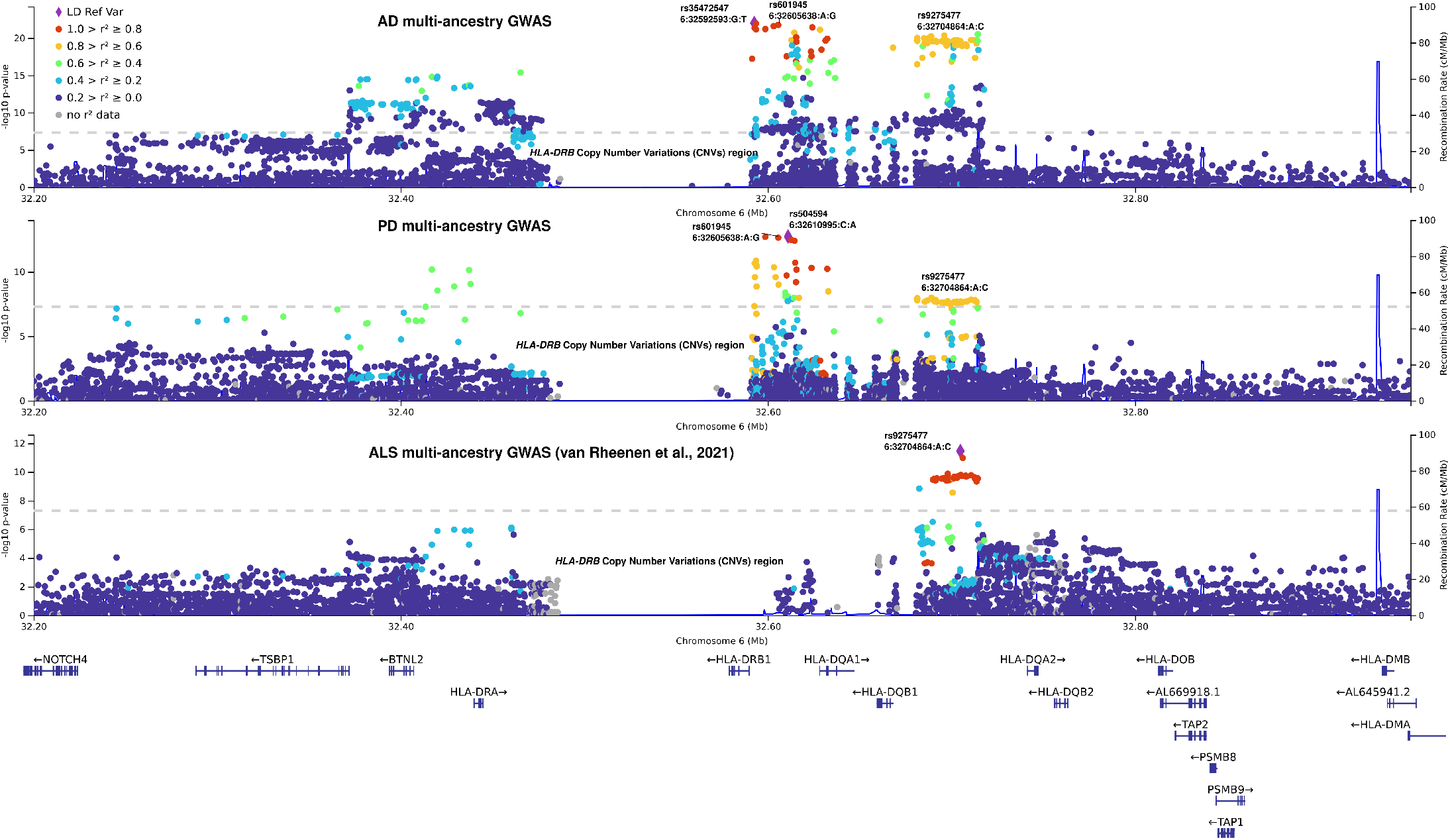
Locuszoom plots of the local-HLA GWAS in multi-ancestry AD, PD, and ALS analyses. The lead variants in the AD and PD GWAS are missing in the summary statistics of the latest ALS GWAS^11^, therefore the colocalization is imperfect.

**Extended Data Figure 3.**
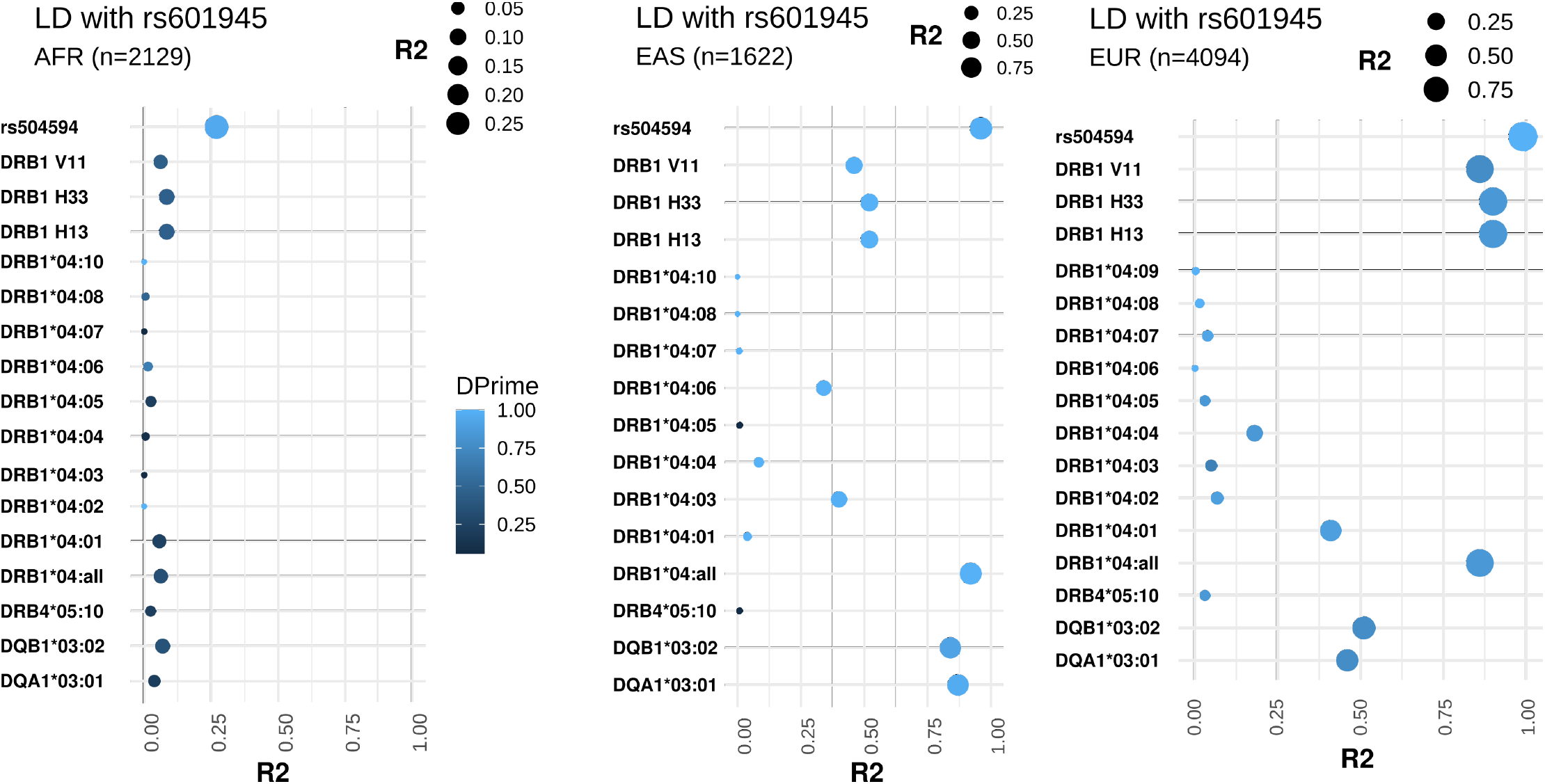
Linkage disequilibrium of the lead SNP with *HLA-DRB1* amino acids and *HLA-DRB1*\*04 allele subtypes.

**Extended Data Figure 4.**
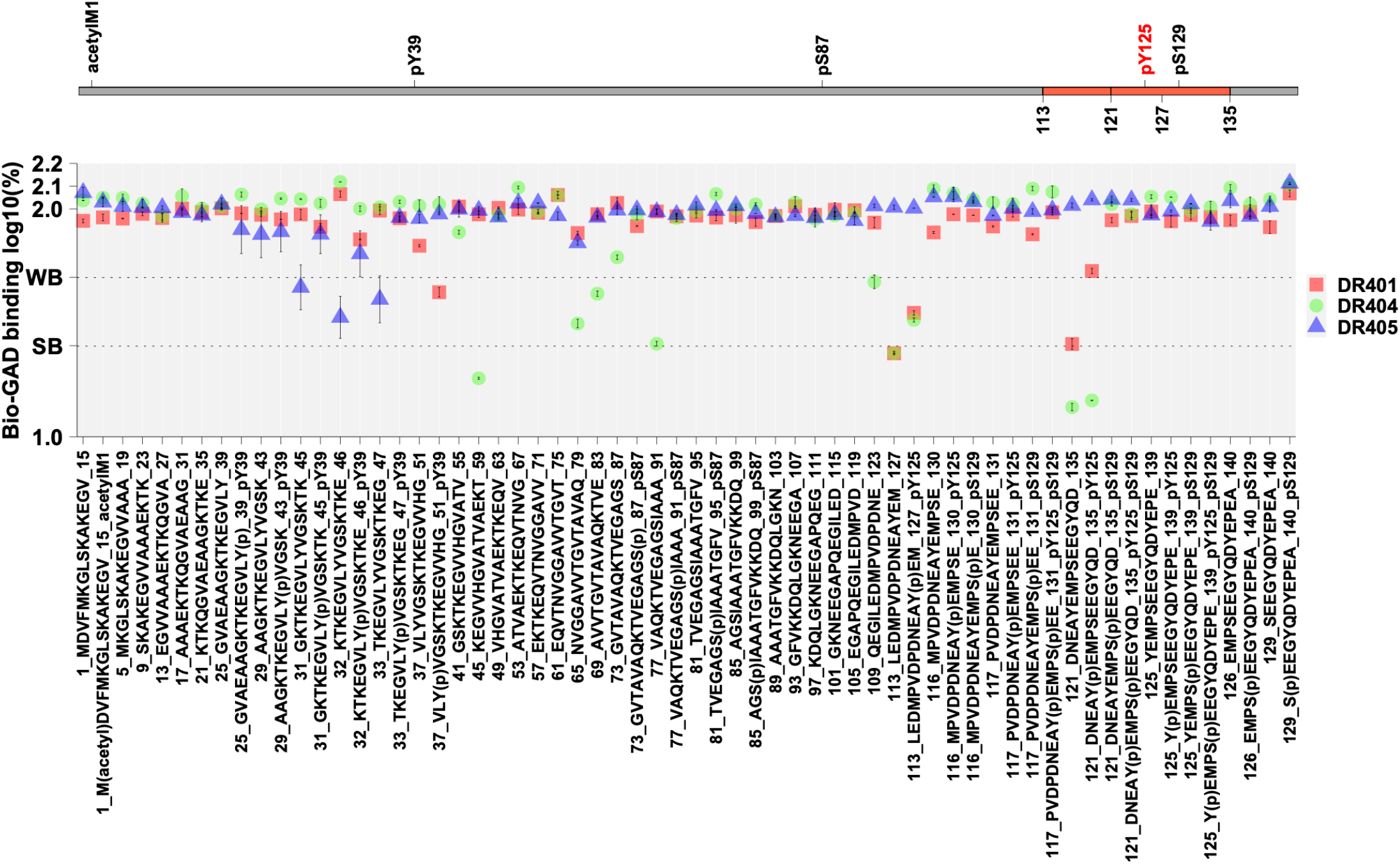
*HLA-DRB1*\*04 subtypes do not show increased to α-synuclein common PTMs. Note that HLA-DRB1*04:04 and HLA-DRB1*04:01 strongly bind to the reference in the region 121-135 but their binding efficiency decreases in the presence of PTMs pY125 and/or pS129. Because PTMs *reduce* central tolerance, this is unlikely to facilitate immune responses. See methods for details.

**Extended Data Figure 5.**
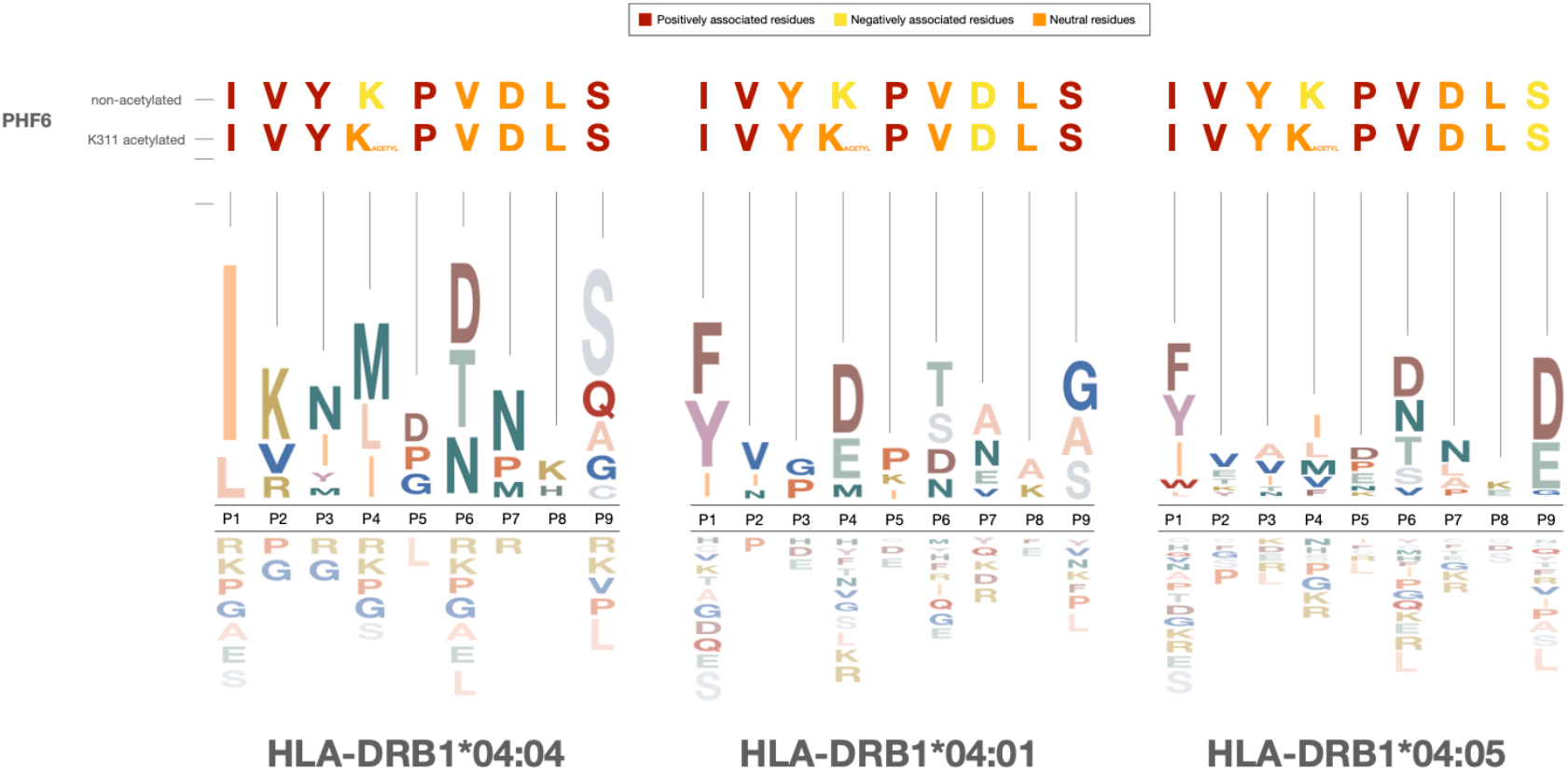
HLA-DRB1*04 binding motifs and PHF6 sensitivity. Residues of PHF6 in their unacetylated or PHF6-K311-acetylated forms are shown and their association with the binding motifs of **(a)** *HLA-DRB1*\*04:04, **(b)** *HLA-DRB1*\*04:01 and **(c)** *HLA-DRB1*\*04:05. Motifs are adapted from Scally et al., 2013^42^ (*HLA-DRB1*\*04:01 and *HLA-DRB1*\*04:04) and Ting et al., 2018^43^ (*HLA-DRB1*\*04:05)

**Extended Data Figure 6.**
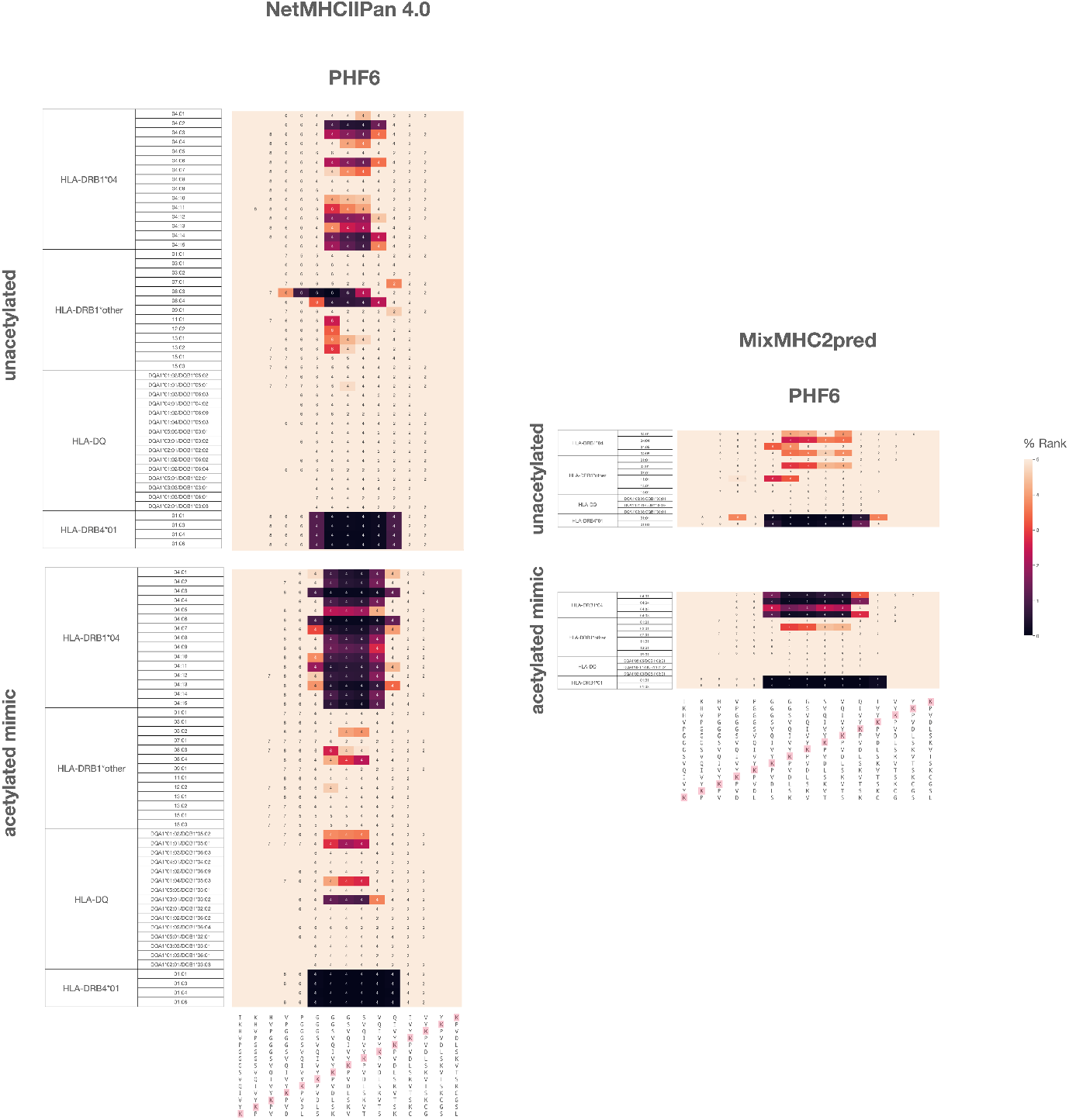
HLA binding predictions for PHF6. Predictions were made using NetMHCIIPan 4.0^74^ (left) using Mixed MHC pred 2 Server^75^ (right). Cmap indicates a percentile rank generated by comparing the peptide’s score against the scores of five million random 15 mers selected from SWISSPROT database (best score = 0%; worst score = 100%). 15mer sequences incorporating 15mer sequences incorporating PHF6 in its unacetylated (bottom) or K311 mimic acetylated (top) form. Annotations within heat map show the position of the lysines of interest (K311 for PHF6, see highlighted sequences) within the 9mer core that is predicted to bind to the HLA molecule. 9mer cores excluding the lysine of interest were not predicted for binding and left blank. Note prediction of PHF6 binding to *HLA-DRB1*\*04 subtypes associated with neurodegeneration, confirmed by our in vitro experiments and the absence of binding to predicted to other common HLA DR and DQ subtypes. One exception is prediction of binding of the PHF6 sequence to *HLA-DRB4*\*01 alleles for both acetylated and non-acetylated PHF6 sequences. Accessory *HLA-DRB4* genes are more weakly expressed than *HLA-DRB1*. These genes are present not only on *HLA-DRB1*\*04 haplotypes, but also on most *HLA-DRB1*\*07:01 and *HLA-DRB1*\*09:01 haplotypes. As *HLA-DRB1*\*07:01 and *HLA-DRB1*\*09:01 haplotypes are not associated with AD or PD (**Supplementary Table 7**), *HLA-DRB4*\*01 is unlikely to be involved in disease predisposition, maybe because of low expression and binding to both acetylated and non-acetylated sequences resulting in central tolerance. The significance of this prediction, not verified through in vitro analysis is unknown.

## References

1. Dourlen, P., Kilinc, D., Malmanche, N., Chapuis, J. & Lambert, J.-C. The new genetic landscape of Alzheimer’s disease: from amyloid cascade to genetically driven synaptic failure hypothesis? Acta Neuropathol 138, 221–236 (2019).

2. Lei, P. et al. Tau protein: relevance to Parkinson’s disease. Int J Biochem Cell Biol 42, 1775–1778 (2010).

3. Petrozziello, T. et al. Novel genetic variants in MAPT and alterations in tau phosphorylation in amyotrophic lateral sclerosis post-mortem motor cortex and cerebrospinal fluid. Brain Pathology n/a, e13035.

4. Strong, M. J., Donison, N. S. & Volkening, K. Alterations in Tau Metabolism in ALS and ALS-FTSD. Front Neurol 11, 598907 (2020).

5. Li, Y. et al. Genomics of Alzheimer’s disease implicates the innate and adaptive immune systems. Cell Mol Life Sci (2021) doi:10.1007/s00018-021-03986-5.

6. Andersen, M. S. et al. Heritability Enrichment Implicates Microglia in Parkinson’s Disease Pathogenesis. Ann Neurol 89, 942–951 (2021).

7. Keren-Shaul, H. et al. A Unique Microglia Type Associated with Restricting Development of Alzheimer’s Disease. Cell 169, 1276-1290.e17 (2017).

8. Bellenguez, C. et al. New insights on the genetic etiology of Alzheimer’s and related dementia. medRxiv 2020.10.01.20200659 (2020) doi:10.1101/2020.10.01.20200659.

9. Kunkle, B. W. et al. Genetic meta-analysis of diagnosed Alzheimer’s disease identifies new risk loci and implicates Aβ, tau, immunity and lipid processing. Nature Genetics 51, 414–430 (2019).

10. Nalls, M. A. et al. Identification of novel risk loci, causal insights, and heritable risk for Parkinson’s disease: a meta-analysis of genome-wide association studies. The Lancet Neurology 18, 1091–1102 (2019).

11. van Rheenen, W. et al. Common and rare variant association analyses in amyotrophic lateral sclerosis identify 15 risk loci with distinct genetic architectures and neuron-specific biology. Nat Genet 53, 1636–1648 (2021).

12. Dhanwani, R. et al. T Cell Responses to Neural Autoantigens Are Similar in Alzheimer’s Disease Patients and Age-Matched Healthy Controls. Front Neurosci 14, 874 (2020).

13. Sulzer, D. et al. T cells from patients with Parkinson’s disease recognize α-synuclein peptides. Nature 546, 656–661 (2017).

14. Lindestam Arlehamn, C. S. et al. α-Synuclein-specific T cell reactivity is associated with preclinical and early Parkinson’s disease. Nat Commun 11, 1875 (2020).

15. Kuhn, I. et al. Serum titers of autoantibodies against α-synuclein and tau in child-and adulthood. Journal of Neuroimmunology 315, 33–39 (2018).

16. Hromadkova, L. & Ovsepian, S. V. Tau-Reactive Endogenous Antibodies: Origin, Functionality, and Implications for the Pathophysiology of Alzheimer’s Disease. J Immunol Res 2019, 7406810 (2019).

17. Holland, C. J., Cole, D. K. & Godkin, A. Re-Directing CD4(+) T Cell Responses with the Flanking Residues of MHC Class II-Bound Peptides: The Core is Not Enough. Front Immunol 4, 172 (2013).

18. Naito, T. et al. Trans-Ethnic Fine-Mapping of the Major Histocompatibility Complex Region Linked to Parkinson’s Disease. Mov Disord 36, 1805–1814 (2021).

19. Yu, E. et al. Fine mapping of the HLA locus in Parkinson’s disease in Europeans. NPJ Parkinsons Dis 7, 84 (2021).

20. Hollenbach, J. A. et al. A specific amino acid motif of HLA-DRB1 mediates risk and interacts with smoking history in Parkinson’s disease. Proc Natl Acad Sci U S A 116, 7419–7424 (2019).

21. Wightman, D. P. et al. A genome-wide association study with 1,126,563 individuals identifies new risk loci for Alzheimer’s disease. Nat Genet 53, 1276–1282 (2021).

22. Steele, N. Z. R. et al. Fine-mapping of the human leukocyte antigen locus as a risk factor for Alzheimer disease: A case-control study. PLoS Med 14, e1002272 (2017).

23. Lindestam Arlehamn, C. S. et al. Widespread Tau-Specific CD4 T Cell Reactivity in the General Population. J Immunol 203, 84–92 (2019).

24. Zheng, X. et al. HIBAG—HLA genotype imputation with attribute bagging. Pharmacogenomics J 14, 192–200 (2014).

25. Matsuki, K. et al. DQ (rather than DR) gene marks susceptibility to narcolepsy. Lancet 339, 1052 (1992).

26. Foo, J. N. et al. Identification of Risk Loci for Parkinson Disease in Asians and Comparison of Risk Between Asians and Europeans: A Genome-Wide Association Study. JAMA Neurol 77, 746–754 (2020).

27. Kang, S. et al. Potential Novel Genes for Late-Onset Alzheimer’s Disease in East-Asian Descent Identified by APOE-Stratified Genome-Wide Association Study. J Alzheimers Dis 82, 1451–1460 (2021).

28. Shigemizu, D. et al. Ethnic and trans-ethnic genome-wide association studies identify new loci influencing Japanese Alzheimer’s disease risk. Transl Psychiatry 11, 151 (2021).

29. Loesch, D. P. et al. Characterizing the Genetic Architecture of Parkinson’s Disease in Latinos. Ann Neurol 90, 353–365 (2021).

30. Kunkle, B. W. et al. Novel Alzheimer Disease Risk Loci and Pathways in African American Individuals Using the African Genome Resources Panel: A Meta-analysis. JAMA Neurol 78, 102 (2021).

31. Jun, G. R. et al. Transethnic genome-wide scan identifies novel Alzheimer’s disease loci. Alzheimers Dement 13, 727–738 (2017).

32. Yamamoto, F. et al. Capturing Differential Allele-Level Expression and Genotypes of All Classical HLA Loci and Haplotypes by a New Capture RNA-Seq Method. Front Immunol 11, 941 (2020).

33. Grifoni, A. et al. Characterization of Magnitude and Antigen Specificity of HLA-DP, DQ, and DRB3/4/5 Restricted DENV-Specific CD4+ T Cell Responses. Front Immunol 10, 1568 (2019).

34. Schneider, J. A. et al. Cognitive impairment, decline and fluctuations in older community-dwelling subjects with Lewy bodies. Brain 135, 3005–3014 (2012).

35. Besser, L. M. et al. The revised national Alzheimer’s coordinating center’s neuropathology form-available data and new analyses. Journal of Neuropathology and Experimental Neurology 77, 717–726 (2018).

36. Raposo, B. et al. T cells specific for post-translational modifications escape intrathymic tolerance induction. Nat Commun 9, 353 (2018).

37. Singhania, A. et al. The TCR repertoire of α-synuclein-specific T cells in Parkinson’s disease is surprisingly diverse. Sci Rep 11, 302 (2021).

38. Lindestam Arlehamn, C. S. et al. α-Synuclein-specific T cell reactivity is associated with preclinical and early Parkinson’s disease. Nat Commun 11, 1875 (2020).

39. Gate, D. et al. Clonally expanded CD8 T cells patrol the cerebrospinal fluid in Alzheimer’s disease. Nature 577, (2020).

40. Wesseling, H. et al. Tau PTM Profiles Identify Patient Heterogeneity and Stages of Alzheimer’s Disease. Cell 183, 1699-1713.e13 (2020).

41. He, H., Liu, Y., Sun, Y. & Ding, F. Misfolding and Self-Assembly Dynamics of Microtubule-Binding Repeats of the Alzheimer-Related Protein Tau. J Chem Inf Model 61, 2916–2925 (2021).

42. Inouye, H., Sharma, D., Goux, W. J. & Kirschner, D. A. Structure of Core Domain of Fibril-Forming PHF/Tau Fragments. Biophysical Journal 90, 1774–1789 (2006).

43. Scally, S. W. et al. A molecular basis for the association of the HLA-DRB1 locus, citrullination, and rheumatoid arthritis. J Exp Med 210, 2569–2582 (2013).

44. Ting, Y. T. et al. The interplay between citrullination and HLA-DRB1 polymorphism in shaping peptide binding hierarchies in rheumatoid arthritis. J Biol Chem 293, 3236–3251 (2018).

45. Trzeciakiewicz, H. et al. An HDAC6-dependent surveillance mechanism suppresses tau-mediated neurodegeneration and cognitive decline. Nat Commun 11, 5522 (2020).

46. Ganguly, P. et al. Tau assembly: the dominant role of PHF6 (VQIVYK) in microtubule binding region repeat R3. J Phys Chem B 119, 4582–4593 (2015).

47. Kim, Y.-C. & Jeong, B.-H. In Silico Evaluation of Acetylation Mimics in the 27 Lysine Residues of Human Tau Protein. Curr Alzheimer Res 16, 379–387 (2019).

48. Guru KrishnaKumar, V., Baweja, L., Ralhan, K. & Gupta, S. Carbamylation promotes amyloidogenesis and induces structural changes in Tau-core hexapeptide fibrils. Biochim Biophys Acta Gen Subj 1862, 2590–2604 (2018).

49. Vaquer-Alicea, J., Diamond, M. I. & Joachimiak, L. A. Tau strains shape disease. Acta Neuropathol 142, 57–71 (2021).

50. Arakhamia, T. et al. Posttranslational Modifications Mediate the Structural Diversity of Tauopathy Strains. Cell 180, 633-644.e12 (2020).

51. Park, S., Lee, J. H., Jeon, J. H. & Lee, M. J. Degradation or aggregation: the ramifications of post-translational modifications on tau. BMB Rep 51, 265–273 (2018).

52. Shin, M.-K. et al. Reducing acetylated tau is neuroprotective in brain injury. Cell 184, 2715-2732.e23 (2021).

53. Kontaxi, C., Piccardo, P. & Gill, A. C. Lysine-Directed Post-translational Modifications of Tau Protein in Alzheimer’s Disease and Related Tauopathies. Front Mol Biosci 4, 56 (2017).

54. Wesseling, H. et al. Tau PTM Profiles Identify Patient Heterogeneity and Stages of Alzheimer’s Disease. Cell 183, 1699-1713.e13 (2020).

55. Acosta, D. M., Mancinelli, C., Bracken, C. & Eliezer, D. Post-translational modifications within tau paired helical filament nucleating motifs perturb microtubule interactions and oligomer formation. Journal of Biological Chemistry 0, (2021).

56. Panda, C. et al. Aggregated Tau-PHF6 (VQIVYK) Potentiates NLRP3 Inflammasome Expression and Autophagy in Human Microglial Cells. Cells 10, 1652 (2021).

57. Carmona-Abellan, M. et al. Microglia is associated with p-Tau aggregates in the olfactory bulb of patients with neurodegenerative diseases. Neurol Sci 42, 1473–1482 (2021).

58. Irwin, D. J. et al. Acetylated tau, a novel pathological signature in Alzheimer’s disease and other tauopathies. Brain 135, 807–818 (2012).

59. Flores-Rodríguez, P. et al. The relationship between truncation and phosphorylation at the C-terminus of tau protein in the paired helical filaments of Alzheimer’s disease. Frontiers in Neuroscience 9, 33 (2015).

60. Sankaranarayanan, S. et al. Passive immunization with phospho-tau antibodies reduces tau pathology and functional deficits in two distinct mouse tauopathy models. PLoS One 10, e0125614 (2015).

61. Šimić, G. et al. Tau Protein Hyperphosphorylation and Aggregation in Alzheimer’s Disease and Other Tauopathies, and Possible Neuroprotective Strategies. Biomolecules 6, 6 (2016).

62. Beecham, G. W. et al. The Alzheimer’s Disease Sequencing Project: Study design and sample selection. Neurol Genet 3, e194 (2017).

63. Bennett, D. A. et al. Overview and findings from the rush Memory and Aging Project. Current Alzheimer research 9, 646–63 (2012).

64. Le Guen, Y. et al. APOE missense variant R145C is associated with increased Alzheimer’s disease risk in African ancestry individuals with the APOE ε3/ε4 genotype. 2021.10.20.21265141 https://www.medrxiv.org/content/10.1101/2021.10.20.21265141v1 (2021) xdoi:10.1101/2021.10.20.21265141.

65. Kang, S. et al. Potential Novel Genes for Late-Onset Alzheimer’s Disease in East-Asian Descent Identified by APOE-Stratified Genome-Wide Association Study. J Alzheimers Dis 82, 1451–1460 (2021).

66. Satake, W. et al. Genome-wide association study identifies common variants at four loci as genetic risk factors for Parkinson’s disease. Nat Genet 41, 1303–1307 (2009).

67. Bennett, D. A. et al. Overview and findings from the rush Memory and Aging Project. Current Alzheimer research 9, 646–63 (2012).

68. Kunkle, B. W. et al. Genetic meta-analysis of diagnosed Alzheimer’s disease identifies new risk loci and implicates Aβ, tau, immunity and lipid processing. Nature Genetics 51, 414–430 (2019).

69. Najar, J. et al. Polygenic risk scores for Alzheimer’s disease are related to dementia risk in APOE ?4 negatives. Alzheimer’s & Dementia: Diagnosis, Assessment & Disease Monitoring 13, e12142 (2021).

70. Gogarten, S. M. et al. Genetic association testing using the GENESIS R/Bioconductor package. Bioinformatics 35, 5346–5348 (2019).

71. Willer, C. J., Li, Y. & Abecasis, G. R. METAL: fast and efficient meta-analysis of genomewide association scans. Bioinformatics 26, 2190–2191 (2010).

72. Giambartolomei, C. et al. Bayesian Test for Colocalisation between Pairs of Genetic Association Studies Using Summary Statistics. PLOS Genetics 10, e1004383 (2014).

73. van Rheenen, W. et al. Common and rare variant association analyses in amyotrophic lateral sclerosis identify 15 risk loci with distinct genetic architectures and neuron-specific biology. Nat Genet 53, 1636–1648 (2021).

74. Lim, J., Bae, S.-C. & Kim, K. Understanding HLA associations from SNP summary association statistics. Sci Rep 9, 1337 (2019).

75. Luo, G. et al. Autoimmunity to hypocretin and molecular mimicry to flu in type 1 narcolepsy. PNAS 115, E12323–E12332 (2018).

