## Supplementary Materials for "Protective association of *HLA-DRB1*\*04 subtypes in neurodegenerative diseases implicates acetylated Tau PHF6 sequences"

#### Table of Contents

**Supplementary Acknowledgments…………………………2**

**Consortia author list……………………………..…………..19**

**Supplementary Methods……………………………….......34**

**Supplementary Tables…………………………………..……46**

**Supplementary Figures……………………………….…….60**

### Additional Acknowledgments

Data for this study were prepared, archived, and distributed by the National Institute on Aging Alzheimer's Disease Data Storage Site (NIAGADS) at the University of Pennsylvania (U24-AG041689), funded by the National Institute on Aging.

**Acknowledgments for the use of ADSP WGS data**

*The Alzheimer's Disease Sequencing Project (ADSP) is comprised of two Alzheimer's Disease (AD) genetics consortia and three National Human Genome Research Institute (NHGRI) funded Large Scale Sequencing and Analysis Centers (LSAC). The two AD genetics consortia are the Alzheimer's Disease Genetics Consortium (ADGC) funded by NIA (U01 AG032984), and the Cohorts for Heart and Aging Research in Genomic Epidemiology (CHARGE) funded by NIA (R01 AG033193), the National Heart, Lung, and Blood Institute (NHLBI), other National Institute of Health (NIH) institutes and other foreign governmental and non-governmental organizations. The Discovery Phase analysis of sequence data is supported through UF1AG047133 (to Drs. Schellenberg, Farrer, Pericak-Vance, Mayeux, and Haines); U01AG049505 to Dr. Seshadri; U01AG049506 to Dr. Boerwinkle; U01AG049507 to Dr. Wijsman; and U01AG049508 to Dr. Goate and the Discovery Extension Phase analysis is supported through U01AG052411 to Dr. Goate, U01AG052410 to Dr. Pericak-Vance and U01 AG052409 to Drs. Seshadri and Fornage.*

*Sequencing for the Follow Up Study (FUS) is supported through U01AG057659 (to Drs. PericakVance, Mayeux, and Vardarajan) and U01AG062943 (to Drs. Pericak-Vance and Mayeux). Data generation and harmonization in the Follow-up Phase is supported by U54AG052427 (to Drs. Schellenberg and Wang). The FUS Phase analysis of sequence data is supported through U01AG058589 (to Drs. Destefano, Boerwinkle, De Jager, Fornage, Seshadri, and Wijsman), U01AG058654 (to Drs. Haines, Bush, Farrer, Martin, and Pericak-Vance), U01AG058635 (to Dr. Goate), RF1AG058066 (to Drs. Haines, Pericak-Vance, and Scott), RF1AG057519 (to Drs. Farrer and Jun), R01AG048927 (to Dr. Farrer), and RF1AG054074 (to Drs. Pericak-Vance and Beecham).*

*The ADGC cohorts include: Adult Changes in Thought (ACT) (UO1 AG006781, UO1 HG004610, UO1 HG006375, U01 HG008657), the Alzheimer's Disease Centers (ADC) ( P30 AG019610, P30 AG013846, P50 AG008702, P50 AG025688, P50 AG047266, P30 AG010133, P50 AG005146, P50 AG005134, P50 AG016574, P50 AG005138, P30 AG008051, P30 AG013854, P30 AG008017, P30 AG010161, P50 AG047366, P30 AG010129, P50 AG016573, P50 AG016570, P50 AG005131, P50 AG023501, P30 AG035982, P30 AG028383, P30 AG010124, P50 AG005133, P50 AG005142, P30 AG012300, P50 AG005136, P50 AG033514, P50 AG005681, and P50 AG047270), the Chicago Health and Aging Project (CHAP) (R01 AG11101, RC4 AG039085, K23 AG030944), Indianapolis Ibadan (R01 AG009956, P30 AG010133), the Memory and Aging Project (MAP) ( R01 AG17917), Mayo Clinic (MAYO) (R01 AG032990, U01 AG046139, R01 NS080820, RF1 AG051504, P50 AG016574), Mayo Parkinson's Disease controls (NS039764, NS071674, 5RC2HG005605), University of Miami (R01 AG027944, R01 AG028786, R01 AG019085, IIRG09133827, A2011048), the Multi-Institutional Research in Alzheimer's Genetic Epidemiology Study (MIRAGE) (R01 AG09029, R01 AG025259), the National Cell Repository for Alzheimer's Disease (NCRAD) (U24 AG21886), the National Institute on Aging Late Onset Alzheimer's Disease Family Study (NIA- LOAD) (R01 AG041797), the Religious Orders Study (ROS) (P30 AG10161, R01 AG15819), the Texas Alzheimer's Research and Care Consortium (TARCC) (funded by the Darrell K Royal Texas Alzheimer's Initiative), Vanderbilt University/Case Western Reserve University (VAN/CWRU) (R01 AG019757, R01 AG021547, R01 AG027944, R01 AG028786, P01 NS026630, and Alzheimer's Association), the Washington Heights-Inwood Columbia Aging Project (WHICAP) (RF1 AG054023), the University of Washington Families (VA Research Merit Grant, NIA: P50AG005136, R01AG041797, NINDS: R01NS069719), the Columbia University HispanicEstudio Familiar de Influencia Genetica de Alzheimer (EFIGA) (RF1 AG015473), the University of Toronto (UT) (funded by Wellcome Trust, Medical Research Council, Canadian Institutes of Health Research), and Genetic Differences (GD) (R01 AG007584). The CHARGE cohorts are supported in part by National Heart, Lung, and Blood Institute (NHLBI) infrastructure grant HL105756 (Psaty), RC2HL102419 (Boerwinkle) and the neurology working group is supported by the National Institute on Aging (NIA) R01 grant AG033193.*

*The CHARGE cohorts participating in the ADSP include the following: Austrian Stroke Prevention Study (ASPS), ASPS-Family study, and the Prospective Dementia Registry-Austria (ASPS/PRODEM-Aus), the Atherosclerosis Risk in Communities (ARIC) Study, the Cardiovascular Health Study (CHS), the Erasmus Rucphen Family Study (ERF), the Framingham Heart Study (FHS), and the Rotterdam Study (RS). ASPS is funded by the Austrian Science Fond (FWF) grant number P20545-P05 and P13180 and the Medical University of Graz. The ASPS-Fam is funded by the Austrian Science Fund (FWF) project I904),the EU Joint Programme - Neurodegenerative Disease Research (JPND) in frame of the BRIDGET project (Austria, Ministry of Science) and the Medical University of Graz and the Steiermärkische Krankenanstalten Gesellschaft. PRODEM-Austria is supported by the Austrian Research Promotion agency (FFG) (Project No. 827462) and by the Austrian National Bank (Anniversary Fund, project 15435. ARIC research is carried out as a collaborative study supported by NHLBI contracts (HHSN268201100005C, HHSN268201100006C, HHSN268201100007C, HHSN268201100008C, HHSN268201100009C, HHSN268201100010C, HHSN268201100011C, and HHSN268201100012C). Neurocognitive data in ARIC is collected by U01 2U01HL096812, 2U01HL096814, 2U01HL096899, 2U01HL096902, 2U01HL096917 from the NIH (NHLBI, NINDS, NIA and NIDCD), and with previous brain MRI examinations funded by R01-HL70825 from the NHLBI. CHS research was supported by contracts HHSN268201200036C, HHSN268200800007C, N01HC55222, N01HC85079, N01HC85080, N01HC85081, N01HC85082, N01HC85083, N01HC85086, and grants U01HL080295 and U01HL130114 from the NHLBI with additional contribution from the National Institute of Neurological Disorders and Stroke (NINDS). Additional support was provided by R01AG023629, R01AG15928, and R01AG20098 from the NIA. FHS research is supported by*

*NHLBI contracts N01-HC-25195 and HHSN268201500001I. This study was also supported by additional grants from the NIA (R01s AG054076, AG049607 and AG033040 and NINDS (R01 NS017950). The ERF study as a part of EUROSPAN (European Special Populations Research Network) was supported by European Commission FP6 STRP grant number 018947 (LSHG-CT-2006-01947) and also received funding from the European Community's Seventh Framework Programme (FP7/2007-2013)/grant agreement HEALTH-F4- 2007-201413 by the European Commission under the programme "Quality of Life and Management of the Living Resources" of 5th Framework Programme (no. QLG2-CT-2002- 01254). High-throughput analysis of the ERF data was supported by a joint grant from the Netherlands Organization for Scientific Research and the Russian Foundation for Basic Research (NWO-RFBR 047.017.043). The Rotterdam Study is funded by Erasmus Medical Center and Erasmus University, Rotterdam, the Netherlands Organization for Health Research and Development (ZonMw), the Research Institute for Diseases in the Elderly (RIDE), the Ministry of Education, Culture and Science, the Ministry for Health, Welfare and Sports, the European Commission (DG XII), and the municipality of Rotterdam. Genetic data sets are also supported by the Netherlands Organization of Scientific Research NWO Investments (175.010.2005.011, 911-03-012), the Genetic Laboratory of the Department of Internal Medicine, Erasmus MC, the Research Institute for Diseases in the Elderly (014-93-015; RIDE2), and the Netherlands Genomics Initiative (NGI)/Netherlands Organization for Scientific Research (NWO) Netherlands Consortium for Healthy Aging (NCHA), project 050-060-810. All studies are grateful to their participants, faculty and staff. The content of these manuscripts is solely the responsibility of the authors and does not necessarily represent the official views of the National Institutes of Health or the U.S. Department of Health and Human Services.*

*The FUS cohorts include: the Alzheimer's Disease Centers (ADC) ( P30 AG019610, P30 AG013846, P50 AG008702, P50 AG025688, P50 AG047266, P30 AG010133, P50 AG005146, P50 AG005134, P50 AG016574, P50 AG005138, P30 AG008051, P30 AG013854, P30 AG008017, P30 AG010161, P50 AG047366, P30 AG010129, P50 AG016573, P50 AG016570, P50 AG005131, P50 AG023501, P30 AG035982, P30 AG028383, P30 AG010124, P50 AG005133, P50 AG005142, P30 AG012300, P50 AG005136, P50 AG033514, P50 AG005681, and P50 AG047270), Alzheimer's Disease Neuroimaging Initiative (ADNI) (U19AG024904), Amish Protective Variant Study (RF1AG058066), Cache County Study (R01AG11380, R01AG031272, R01AG21136, RF1AG054052), Case Western Reserve University Brain Bank (CWRUBB) (P50AG008012), Case Western Reserve University Rapid Decline (CWRURD) (RF1AG058267, NU38CK000480), CubanAmerican Alzheimer's Disease Initiative (CuAADI) (3U01AG052410), Estudio Familiar de Influencia Genetica en Alzheimer (EFIGA) (5R37AG015473, RF1AG015473, R56AG051876), Genetic and Environmental Risk Factors for Alzheimer Disease Among African Americans Study (GenerAAtions) (2R01AG09029, R01AG025259, 2R01AG048927), Gwangju Alzheimer and Related Dementias Study (GARD) (U01AG062602), Hussman Institute for Human Genomics Brain Bank (HIHGBB) (R01AG027944, Alzheimer's Association "Identification of Rare Variants in Alzheimer Disease"), Ibadan Study of Aging (IBADAN) (5R01AG009956), Mexican Health and Aging Study (MHAS) (R01AG018016),*

*Multi-Institutional Research in Alzheimer's Genetic Epidemiology (MIRAGE) (2R01AG09029, R01AG025259, 2R01AG048927), Northern Manhattan Study (NOMAS) (R01NS29993), Peru Alzheimer's Disease Initiative (PeADI) (RF1AG054074), Puerto Rican 1066 (PR1066) (Wellcome Trust (GR066133/GR080002), European Research Council (340755)), Puerto Rican Alzheimer Disease Initiative (PRADI) (RF1AG054074), Reasons for Geographic and Racial Differences in Stroke (REGARDS) (U01NS041588), Research in African American Alzheimer Disease Initiative (REAAADI) (U01AG052410), Rush Alzheimer's Disease Center (ROSMAP) (P30AG10161, R01AG15819, R01AG17919), University of Miami Brain Endowment Bank (MBB), and University of Miami/Case Western/North Carolina A&T African American (UM/CASE/NCAT) (U01AG052410, R01AG028786).*

*The four LSACs are: the Human Genome Sequencing Center at the Baylor College of Medicine (U54 HG003273), the Broad Institute Genome Center (U54HG003067), The American Genome Center at the Uniformed Services University of the Health Sciences (U01AG057659), and the Washington University Genome Institute (U54HG003079).*

*Biological samples and associated phenotypic data used in primary data analyses were stored at Study Investigators institutions, and at the National Cell Repository for Alzheimer's Disease (NCRAD, U24AG021886) at Indiana University funded by NIA. Associated Phenotypic Data used in primary and secondary data analyses were provided by Study Investigators, the NIA funded Alzheimer's Disease Centers (ADCs), and the National Alzheimer's Coordinating Center (NACC, U01AG016976) and the National Institute on Aging Genetics of Alzheimer's Disease Data Storage Site (NIAGADS, U24AG041689) at the University of Pennsylvania, funded by NIA This research was supported in part by the Intramural Research Program of the National Institutes of health, National Library of Medicine. Contributors to the Genetic Analysis Data included Study Investigators on projects that were individually funded by NIA, and other NIH institutes, and by private U.S. organizations, or foreign governmental or nongovernmental organizations.*

An up to date acknowledgment statement can be found on the ADSP site: https://www.niagads.org/adsp/content/acknowledgement-statement.

Data collection and sharing for this project was funded by the Alzheimer's Disease Neuroimaging Initiative (ADNI) (National Institutes of Health Grant U01 AG024904) and DOD ADNI (Department of Defense award number W81XWH-12-2-0012). ADNI is funded by the National Institute on Aging, the National Institute of Biomedical Imaging and Bioengineering, and through generous contributions from the following: AbbVie, Alzheimer's Association; Alzheimer's Drug Discovery Foundation; Araclon Biotech; BioClinica, Inc.; Biogen; Bristol-Myers Squibb Company; CereSpir, Inc.; Cogstate; Eisai Inc.; Elan Pharmaceuticals, Inc.; Eli Lilly and Company; EuroImmun; F. Hoffmann-La Roche Ltd and its affiliated company Genentech, Inc.; Fujirebio; GE Healthcare; IXICO Ltd.; Janssen Alzheimer Immunotherapy Research & Development, LLC.; Johnson & Johnson Pharmaceutical Research & Development

LLC.; Lumosity; Lundbeck; Merck & Co., Inc.; Meso Scale Diagnostics, LLC.; NeuroRx Research; Neurotrack Technologies; Novartis Pharmaceuticals Corporation; Pfizer Inc.; Piramal Imaging; Servier; Takeda Pharmaceutical Company; and Transition Therapeutics. The Canadian Institutes of Health Research is providing funds to support ADNI clinical sites in Canada. Private sector contributions are facilitated by the Foundation for the National Institutes of Health (www.fnih.org). The grantee organization is the Northern California Institute for Research and Education, and the study is coordinated by the Alzheimer's Therapeutic Research Institute at the University of Southern California. ADNI data are disseminated by the Laboratory for Neuro Imaging at the University of Southern California.

Additional information to include in an acknowledgment statement can be found on the LONI site: https://adni.loni.usc.edu/wp-content/uploads/how_to_apply/ADNI_Data_Use_Agreement.pdf.

The Alzheimer's Disease Genetics Consortium (ADGC) supported sample preparation, whole exome sequencing and data processing through NIA grant U01AG032984. Sequencing data generation and harmonization is supported by the Genome Center for Alzheimer's Disease, U54AG052427, and data sharing is supported by NIAGADS, U24AG041689. Samples from the National Centralized Repository for Alzheimer's Disease and Related Dementias (NCRAD), which receives government support under a cooperative agreement grant (U24 AG021886) awarded by the National Institute on Aging (NIA), were used in this study. We thank contributors who collected samples used in this study, as well as patients and their families, whose help and participation made this work possible. NIH grants supported enrollment and data collection for the individual studies including: GenerAAtions R01AG20688 (PI M. Daniele Fallin, PhD); Miami/Duke R01 AG027944, R01 AG028786 (PI Margaret A. Pericak-Vance, PhD); NC A&T P20 MD000546, R01 AG28786-01A1 (PI Goldie S. Byrd, PhD); Case Western (PI Jonathan L. Haines, PhD); MIRAGE R01 AG009029 (PI Lindsay A. Farrer, PhD); ROS P30AG10161, R01AG15819, R01AG30146, TGen (PI David A. Bennett, MD); MAP R01AG17917, R01AG15819, TGen (PI David A. Bennett, MD). The NACC database is funded by NIA/NIH Grant U01 AG016976. NACC data are contributed by the NIA-funded ADCs: P30 AG019610 (PI Eric Reiman, MD), P30 AG013846 (PI Neil Kowall, MD), P30 AG062428-01 (PI James Leverenz, MD) P50 AG008702 (PI Scott Small, MD), P50 AG025688 (PI Allan Levey, MD, PhD), P50 AG047266 (PI Todd Golde, MD, PhD), P30 AG010133 (PI Andrew Saykin, PsyD), P50 AG005146 (PI Marilyn Albert, PhD), P30 AG062421-01 (PI Bradley Hyman, MD, PhD), P30 AG062422-01 (PI Ronald Petersen, MD, PhD), P50 AG005138 (PI Mary Sano, PhD), P30 AG008051 (PI Thomas Wisniewski, MD), P30 AG013854 (PI Robert Vassar, PhD), P30 AG008017 (PI Jeffrey Kaye, MD), P30 AG010161 (PI David Bennett, MD), P50 AG047366 (PI Victor Henderson, MD, MS), P30 AG010129 (PI Charles DeCarli, MD), P50 AG016573 (PI Frank LaFerla, PhD), P30 AG062429-01(PI James Brewer, MD, PhD), P50 AG023501 (PI Bruce Miller, MD), P30 AG035982 (PI Russell Swerdlow, MD), P30 AG028383 (PI Linda Van Eldik, PhD), P30 AG053760 (PI Henry Paulson, MD, PhD), P30 AG010124 (PI John Trojanowski, MD, PhD), P50 AG005133 (PI Oscar Lopez, MD), P50 AG005142 (PI Helena Chui, MD), P30 AG012300 (PI

Roger Rosenberg, MD), P30 AG049638 (PI Suzanne Craft, PhD), P50 AG005136 (PI Thomas Grabowski, MD), P30 AG062715-01 (PI Sanjay Asthana, MD, FRCP), P50 AG005681 (PI John Morris, MD), P50 AG047270 (PI Stephen Strittmatter, MD, PhD).

This work was supported by grants from the National Institutes of Health (R01AG044546, P01AG003991, RF1AG053303, R01AG058501, U01AG058922, RF1AG058501 and R01AG057777). The recruitment and clinical characterization of research participants at Washington University were supported by NIH P50 AG05681, P01 AG03991, and P01 AG026276. This work was supported by access to equipment made possible by the Hope Center for Neurological Disorders, and the Departments of Neurology and Psychiatry at Washington University School of Medicine.

We thank the contributors who collected samples used in this study, as well as patients and their families, whose help and participation made this work possible. Members of the National Institute on Aging Late-Onset Alzheimer Disease/National Cell Repository for Alzheimer Disease (NIA-LOAD NCRAD) Family Study Group include the following: Richard Mayeux, MD, MSc; Martin Farlow, MD; Tatiana Foroud, PhD; Kelley Faber, MS; Bradley F. Boeve, MD; Neill R. Graff-Radford, MD; David A. Bennett, MD; Robert A. Sweet, MD; Roger Rosenberg, MD; Thomas D. Bird, MD; Carlos Cruchaga, PhD; and Jeremy M. Silverman, PhD.

This work was partially supported by grant funding from NIH R01 AG039700 and NIH P50 AG005136. Subjects and samples used here were originally collected with grant funding from NIH U24 AG026395, U24 AG021886, P50 AG008702, P01 AG007232, R37 AG015473, P30 AG028377, P50 AG05128, P50 AG16574, P30 AG010133, P50 AG005681, P01 AG003991, U01MH046281, U01 MH046290 and U01 MH046373. The funders had no role in study design, analysis or preparation of the manuscript. The authors declare no competing interests.

This work was supported by the National Institutes of Health (R01 AG027944, R01 AG028786 to MAPV, R01 AG019085 to JLH, P20 MD000546); a joint grant from the Alzheimer's Association (SG-14-312644) and the Fidelity Biosciences Research Initiative to MAPV; the BrightFocus Foundation (A2011048 to MAPV). NIA-LOAD Family-Based Study supported the collection of samples used in this study through NIH grants U24 AG026395 and R01 AG041797 and the MIRAGE cohort was supported through the NIH grants R01 AG025259 and R01 AG048927. We thank contributors, including the Alzheimer's disease Centers who collected samples used in this study, as well as patients and their families, whose help and participation made this work possible. Study design: HNC, BWK, JLH, MAPV; Sample collection: MLC, JMV, RMC, LAF, JLH, MAPV; Whole exome sequencing and Sanger sequencing: SR, PLW; Sequencing data analysis: HNC, BWK, KLHN, SR, MAK, JRG, ERM, GWB, MAPV; Statistical analysis: BWK, KLHN, JMJ, MAPV; Preparation of manuscript: HNC, BWK. The authors jointly discussed the experimental results throughout the duration of the study. All authors read and approved the final manuscript.

Data collection and sharing for this project was supported by the Washington Heights-Inwood Columbia Aging Project (WHICAP, PO1AG07232, R01AG037212, RF1AG054023) funded by the National Institute on Aging (NIA) and by the National Center for Advancing Translational Sciences, National Institutes of Health, through Grant Number UL1TR001873. This manuscript has been reviewed by WHICAP investigators for scientific content and consistency of data interpretation with previous WHICAP Study publications. We acknowledge the WHICAP study participants and the WHICAP research and support staff for their contributions to this study.

This work was supported by grants from the National Institutes of Health (R01AG044546, P01AG003991, RF1AG053303, R01AG058501, U01AG058922, RF1AG058501 and R01AG057777). The recruitment and clinical characterization of research participants at Washington University were supported by NIH P50 AG05681, P01 AG03991, and P01 AG026276. This work was supported by access to equipment made possible by the Hope Center for Neurological Disorders, and the Departments of Neurology and Psychiatry at Washington University School of Medicine.

We thank the contributors who collected samples used in this study, as well as patients and their families, whose help and participation made this work possible. Members of the National Institute on Aging Late-Onset Alzheimer Disease/National Cell Repository for Alzheimer Disease (NIA-LOAD NCRAD) Family Study Group include the following: Richard Mayeux, MD, MSc; Martin Farlow, MD; Tatiana Foroud, PhD; Kelley Faber, MS; Bradley F. Boeve, MD; Neill R. Graff-Radford, MD; David A. Bennett, MD; Robert A. Sweet, MD; Roger Rosenberg, MD; Thomas D. Bird, MD; Carlos Cruchaga, PhD; and Jeremy M. Silverman, PhD.

This work was supported by grants from the National Institutes of Health (R01AG044546, P01AG003991, RF1AG053303, R01AG058501, U01AG058922, RF1AG058501 and R01AG057777). The recruitment and clinical characterization of research participants at Washington University were supported by NIH P50 AG05681, P01 AG03991, and P01 AG026276. This work was supported by access to equipment made possible by the Hope Center for Neurological Disorders, and the Departments of Neurology and Psychiatry at Washington University School of Medicine.

We thank the contributors who collected samples used in this study, as well as patients and their families, whose help and participation made this work possible. Members of the National Institute on Aging Late-Onset Alzheimer Disease/National Cell Repository for Alzheimer Disease (NIA-LOAD NCRAD) Family Study Group include the following: Richard Mayeux, MD, MSc; Martin Farlow, MD; Tatiana Foroud, PhD; Kelley Faber, MS; Bradley F. Boeve, MD; Neill R. Graff-Radford, MD; David A. Bennett, MD; Robert A. Sweet, MD; Roger Rosenberg, MD; Thomas D. Bird, MD; Carlos Cruchaga, PhD; and Jeremy M. Silverman, PhD.

**Mayo RNAseq Study-** Study data were provided by the following sources: The Mayo Clinic Alzheimer's Disease Genetic Studies, led by Dr. Nilufer Ertekin-Taner and Dr. Steven G.

Younkin, Mayo Clinic, Jacksonville, FL using samples from the Mayo Clinic Study of Aging, the Mayo Clinic Alzheimer's Disease Research Center, and the Mayo Clinic Brain Bank. Data collection was supported through funding by NIA grants P50 AG016574, R01 AG032990, U01 AG046139, R01 AG018023, U01 AG006576, U01 AG006786, R01 AG025711, R01 AG017216, R01 AG003949, NINDS grant R01 NS080820, CurePSP Foundation, and support from Mayo Foundation. Study data includes samples collected through the Sun Health Research Institute Brain and Body Donation Program of Sun City, Arizona. The Brain and Body Donation Program is supported by the National Institute of Neurological Disorders and Stroke (U24 NS072026 National Brain and Tissue Resource for Parkinson's Disease and Related Disorders), the National Institute on Aging (P30 AG19610 Arizona Alzheimer's Disease Core Center), the Arizona Department of Health Services (contract 211002, Arizona Alzheimer's Research Center), the Arizona Biomedical Research Commission (contracts 4001, 0011, 05-901 and 1001 to the Arizona Parkinson's Disease Consortium) and the Michael J. Fox Foundation for Parkinson's Research

**ROSMAP**- We are grateful to the participants in the Religious Order Study, the Memory and Aging Project. This work is supported by the US National Institutes of Health [U01 AG046152, R01 AG043617, R01 AG042210, R01 AG036042, R01 AG036836, R01 AG032990, R01 AG18023, RC2 AG036547, P50 AG016574, U01 ES017155, KL2 RR024151, K25 AG041906-01, R01 AG30146, P30 AG10161, R01 AG17917, R01 AG15819, K08 AG034290, P30 AG10161 and R01 AG11101.

**Mount Sinai Brain Bank (MSBB)**- This work was supported by the grants R01AG046170, RF1AG054014, RF1AG057440 and R01AG057907 from the NIH/National Institute on Aging (NIA). R01AG046170 is a component of the AMP-AD Target Discovery and Preclinical Validation Project. Brain tissue collection and characterization was supported by NIH HHSN271201300031C.

This study was supported by the National Institute on Aging (NIA) grants AG030653, AG041718, AG064877 and P30-AG066468.

We would like to thank study participants, their families, and the sample collectors for their invaluable contributions. This research was supported in part by the National Institute on Aging grant U01AG049508 (PI Alison M. Goate). This research was supported in part by Genentech, Inc. (PI Alison M. Goate, Robert R. Graham).

The NACC database is funded by NIA/NIH Grant U01 AG016976. NACC data are contributed by these NIA-funded ADCs: P30 AG013846 (PI Neil Kowall, MD), P50 AG008702 (PI Scott Small, MD), P50 AG025688 (PI Allan Levey, MD, PhD), P30 AG010133 (PI Andrew Saykin, PsyD), P50 AG005146 (PI Marilyn Albert, PhD), P50 AG005134 (PI Bradley Hyman, MD, PhD), P50 AG016574 (PI Ronald Petersen, MD, PhD), P30 AG013854 (PI M. Marsel Mesulam, MD), P30 AG008017 (PI Jeffrey Kaye, MD), P30 AG010161 (PI David Bennett, MD), P30 AG010129

(PI Charles DeCarli, MD), P50 AG016573 (PI Frank LaFerla, PhD), P50 AG005131 (PI Douglas Galasko, MD), P30 AG028383 (PI Linda Van Eldik, PhD), P30 AG010124 (PI John Trojanowski, MD, PhD), P50 AG005142 (PI Helena Chui, MD), P30 AG012300 (PI Roger Rosenberg, MD), P50 AG005136 (PI Thomas Grabowski, MD), P50 AG005681 (PI John Morris, MD), P30 AG028377 (Kathleen Welsh-Bohmer, PhD), and P50 AG008671 (PI Henry Paulson, MD, PhD).

Samples from the National Cell Repository for Alzheimer's Disease (NCRAD), which receives government support under a cooperative agreement grant (U24 AG21886) awarded by the National Institute on Aging (NIA), were used in this study. We thank contributors who collected samples used in this study, as well as patients and their families, whose help and participation made this work possible.

The Alzheimer's Disease Genetics Consortium supported the collection of samples used in this study through National Institute on Aging (NIA) grants U01AG032984 and RC2AG036528.

We acknowledge the generous contributions of the Cache County Memory Study participants. Sequencing for this study was funded by RF1AG054052 (PI: John S.K. Kauwe)

**Acknowledgments for the use of GWAS data distributed by NIAGADS**

The NIA Genetics of Alzheimer's Disease Data Storage Site (NIAGADS) is supported by a collaborative agreement from the National Institute on Aging, U24AG041689.

NG00047: The NIA supported this work through grants U01-AG032984, RC2-AG036528, U01-AG016976 (Dr Kukull); U24 AG026395, U24 AG026390, R01AG037212, R37 AG015473 (Dr Mayeux); K23AG034550 (Dr Reitz); U24-AG021886 (Dr Foroud); R01AG009956, RC2 AG036650 (Dr Hall); UO1 AG06781, UO1 HG004610 (Dr Larson); R01 AG009029 (Dr Farrer); 5R01AG20688 (Dr Fallin); P50 AG005133, AG030653 (Dr Kamboh); R01 AG019085 (Dr Haines); R01 AG1101, R01 AG030146, RC2 AG036650 (Dr Evans); P30AG10161, R01AG15819, R01AG30146, R01AG17917, R01AG15819 (Dr Bennett); R01AG028786 (Dr Manly); R01AG22018, P30AG10161 (Dr Barnes); P50AG16574 (Dr Ertekin-Taner, Dr Graff-Radford), R01 AG032990 (Dr Ertekin-Taner), KL2 RR024151 (Dr Ertekin-Taner); R01 AG027944, R01 AG028786 (Dr Pericak-Vance); P20 MD000546, R01 AG28786-01A1 (Dr Byrd); AG005138 (Dr Buxbaum); P50 AG05681, P01 AG03991, P01 AG026276 (Dr Goate); and P30AG019610, P30AG13846, U01-AG10483, R01CA129769, R01MH080295, R01AG017173, R01AG025259, R01AG33193, P50AG008702, P30AG028377, AG05128, AG025688, P30AG10133, P50AG005146, P50AG005134, P01AG002219, P30AG08051, MO1RR00096, UL1RR029893, P30AG013854, P30AG008017, R01AG026916, R01AG019085, P50AG016582, UL1RR02777, R01AG031581, P30AG010129, P50AG016573, P50AG016575, P50AG016576, P50AG016577, P50AG016570, P50AG005131, P50AG023501, P50AG019724, P30AG028383, P50AG008671, P30AG010124, P50AG005142, P30AG012300, AG010491, AG027944, AG021547, AG019757, P50AG005136 (Alzheimer Disease GeneticsConsortium [ADGC]). We thank Creighton Phelps,

Stephen Synder, and Marilyn Miller from the NIA, who are ex-officio members of the ADGC. Support was also provided by the Alzheimer's Association (IIRG-08-89720 [Dr Farrer] and IIRG-05-14147 [Dr Pericak-Vance]), National Institute of Neurological Disorders and Stroke grant NS39764, National Institute of Mental Health grant MH60451, GlaxoSmithKline, and the Office of Research and Development, Biomedical Laboratory Research Program, US Department of Veterans Affairs Administration. For the ADGC, biological samples and associated phenotypic data used in primary data analyses were stored at principal investigators' institutions and at the National Cell Repository for Alzheimer's Disease (NCRAD) at Indiana University, funded by the NIA. Associated phenotypic data used in secondary data analyses were stored at the National Alzheimer's Coordinating Center and at the NIA Alzheimer's Disease Data Storage Site at the University of Pennsylvania, funded by the NIA. Contributors to the genetic analysis data included principal investigators on projects ndividually funded by the NIA, other NIH institutes, or private entities.

**Acknowledgments for the use of MARS and LATC**

We thank all Minority Aging Research Study and Latino Core participants and the Rush Alzheimer’s Disease Center staff. This database was funded by the NIH/NIA grants R01AG22018 (MARS) and P30AG 072975 (ADC).

**Acknowledgments for CCHS and CGPS**

We thank the staff and participants of the CCHS and CGPS for their important contributions. This work was supported by the Research Council at Rigshospitalet and the Lundbeck Foundation (grant #R278-2018-804).

**Acknowledgments for EADB**

The work for this manuscript was further supported by the CoSTREAM project (www.costream.eu) and funding from the European Union's Horizon 2020 research and innovation programme under grant agreement No 667375. This work is also funded by la fondation pour la recherché médicale (FRM) (EQU202003010147) Italian Ministry of Health (Ricerca Corrente); Ministero dell'Istruzione, del l'Università e della Ricerca–MIUR project “Dipartimenti di Eccellenza 2018–2022” to Department of Neuroscience “Rita Levi Montalcini”, University of Torino (IR), and AIRAlzh Onlus-ANCC-COOP (SB); Partly supported by “Ministero della Salute”, I.R.C.C.S. Research Program, Ricerca Corrente 2018-2020, Linea n. 2 “Meccanismi genetici, predizione e terapie innovative delle malattie complesse” and by the “5 x 1000” voluntary contribution to the Fondazione I.R.C.C.S. Ospedale “Casa Sollievo della Sofferenza”; and RF-2018-12366665, Fondi per la ricerca 2019 (Sandro Sorbi). Copenhagen General Population Study (CGPS): We thank staff and participants of the CGPS for their important contributions. Karolinska Institutet AD cohort: Dr. Graff and co-authors of the Karolinska Institutet AD cohort report grants from Swedish Research Council (VR) 2015-02926, 2018-02754, 2015-06799, Swedish Alzheimer Foundation, Stockholm County Council ALF and resarch school, Karolinska Institutet StratNeuro, Swedish Demensfonden, and Swedish brain foundation, during the conduct of the study. ADGEN: This work was supported by Academy of Finland (grant numbers 307866); Sigrid Jusélius Foundation; the Strategic Neuroscience Funding of the University of Eastern Finland; EADB project in the JPNDCO-FUND program (grant number 301220). CBAS: The research leading to these results has received funding from the EEA/ Norway Grants 2014-2021 and the Technology Agency of the Czech Republic - project number TO01000215 Supported by Ministry of Health of the Czech Republic, grant nr. 19-04-00560 the Ministry of Health, Czech Republic—conceptual development of research organization, University Hospital Motol, Prague, Czech Republic Grant No. 00064203; the Czech Ministry of Health Project AZV Grant No. 19-04-00560; and Institutional Support of Excellence 2. LF UK Grant No. 6990332. Jakub Hort was supported by the Ministry of Health of the Czech Republic, grant no. AZV-NV18-04-00455. LF UK Grant No. 699012. CNRMAJ-Rouen: This study received fundings from the Centre National de Référence Malades Alzheimer Jeunes (CNRMAJ). The Finnish Geriatric Intervention Study for the Prevention of Cognitive Impairment and Disability (FINGER) data collection was supported by grants from the Academy of Finland, La Carita Foundation, Juho Vainio Foundation, Novo Nordisk Foundation, Finnish Social Insurance Institution, Ministry of Education and Culture Research Grants, Yrjö Jahnsson Foundation, Finnish Cultural Foundation South Osthrobothnia Regional Fund, and EVO/State Research Funding grants of University Hospitals of Kuopio, Oulu and Turku, Seinäjoki Central Hospital and Oulu City Hospital, Alzheimer's Research & Prevention Foundation USA, AXA Research Fund, Knut and Alice Wallenberg Foundation Sweden, Center for Innovative Medicine (CIMED) at Karolinska Institutet Sweden, and Stiftelsen Stockholms sjukhem Sweden. FINGER cohort genotyping was funded by EADB project in the JPND CO-FUND (grant number 301220). Research at the Belgian EADB site is funded in part by the Alzheimer Research Foundation (SAO-FRA), The Research Foundation Flanders (FWO), and the University of Antwerp Research Fund. FK is supported by a BOF DOCPRO fellowship of the University of Antwerp Research Fund. SNAC-K is financially supported by the Swedish Ministry of Health and Social Affairs, the participating County Councils and Municipalities, and the Swedish Research Council. BDR Bristol: We would like to thank the South West Dementia Brain Bank (SWDBB) for providing brain tissue for this study. The SWDBB is part of the Brains for Dementia Research programme, jointly funded by Alzheimer's Research UK and Alzheimer's Society and is supported by BRACE (Bristol Research into Alzheimer's and Care of the Elderly) and the Medical Research Council. BDR Manchester: We would like to thank the Manchester Brain Bankfor providing brain tissue for this study. The Manchester Brain Bank is part of the Brains for Dementia Research programme, jointly funded by Alzheimer's Research UK and Alzheimer's Society. BDR KCL: Human post-mortem tissue was provided by the London Neurodegenerative Diseases Brain Bank which receives funding from the UK Medical Research Council and as part of the Brains for Dementia Research programme, jointly funded by Alzheimer's Research UK and the Alzheimer's Society. The CFAS Wales study was funded by the ESRC (RES-060-25-0060) and HEFCW as 'Maintaining function and well-being in later life: a longitudinal cohort study', (Principal Investigators: R.T Woods, L.Clare, G.Windle, V. Burholt, J. Philips, C. Brayne, C. McCracken, K. Bennett, F. Matthews). We are grateful to the NISCHR Clinical Research Centre for their assistance in tracing participants and in interviewing and in collecting blood samples, and to general practices in the study areas for their cooperation. MRC: We thank all individuals who participated in this study. Cardiff University was supported by the Alzheimer's Society (AS; grant RF014/164) and the Medical Research Council (MRC; grants G0801418/1, MR/K013041/1, MR/L023784/1) (R. Sims is an AS Research Fellow). Cardiff University was also supported by the European Joint Programme for Neurodegenerative Disease (JPND; grant MR/L501517/1), Alzheimer's Research UK (ARUK; grant ARUK-PG2014-1), the Welsh Assembly Government (grant SGR544:CADR), Brain's for dementia Research and a donation from the Moondance Charitable Foundation. Cardiff University acknowledges the support of the UK Dementia Research Institute, of which J. Williams is an associate director. Cambridge University acknowledges support from the MRC. Patient recruitment for the MRC Prion Unit/UCL Department of Neurodegenerative Disease collection was supported by the UCLH/UCL Biomedical Centre and NIHR Queen Square Dementia Biomedical Research Unit. The University of Southampton acknowledges support from the AS. King's College London was supported by the NIHR Biomedical Research Centre for Mental Health and the Biomedical Research Unit for Dementia at the South London and Maudsley NHS Foundation Trust and by King's College London and the MRC. ARUK and the Big Lottery Fund provided support to Nottingham University. Alfredo Ramirez: Part of the work was funded by the JPND EADB grant (German Federal Ministry of Education and Research (BMBF) grant: 01ED1619A). Alfredo Ramirez is also supported by the German Research Foundation (DFG) grants Nr: RA 1971/6-1, RA1971/7-1, and RA 1971/8-1. German Study on Ageing, Cognition and Dementia in Primary Care Patients (AgeCoDe): This study/publication is part of the German Research Network on Dementia (KND), the German Research Network on Degenerative Dementia (KNDD; German Study on Ageing, Cognition and Dementia in Primary Care Patients; AgeCoDe), and the Health Service Research Initiative (Study on Needs, health service use, costs and health-related quality of life in a large sample of oldestold primary care patients (85+; AgeQualiDe)) and was funded by the German Federal Ministry of Education and Research (grants KND: 01GI0102, 01GI0420, 01GI0422, 01GI0423, 01GI0429, 01GI0431, 01GI0433, 01GI0434; grants KNDD: 01GI0710, 01GI0711, 01GI0712, 01GI0713, 01GI0714, 01GI0715, 01GI0716; grants Health Service Research Initiative: 01GY1322A, 01GY1322B, 01GY1322C, 01GY1322D, 01GY1322E, 01GY1322F, 01GY1322G). VITA study: The VITA study was supported by the Ludwig Boltzmann Institute of Aging Research, Vienna, Austria. The former VITA study group should be acknowledged: W. Danielczyk, P. Fischer, G. Gatterer, K Jellinger, S Jugwirth, P. Riederer, KH Tragl, S Zehetmayer. Vogel Study: This work was financed by a research grant of the ''Vogelstiftung Dr. Eckernkamp''. HELIAD study: This study was supported by the grants: IIRG-09-133014 from the Alzheimer's Association, 189 10276/8/9/2011 from the ESPA-EU program Excellence Grant (ARISTEIA) and the ΔΥ2β/οικ.51657/14.4.2009 of the Ministry for Health and Social Solidarity (Greece). Biobank Department of Psychiatry, UMG: Biobank Department of Psychiatry, UMG: Prof. Jens Wiltfang is supported is supported by an Ilídio Pinho professorship, iBiMED (UIDB/04501/2020) at the University of Aveiro, Portugal. Lausanne study: This work was supported by grants from the Swiss National Research Foundation (SNF 320030_141179). PAGES study: Harald Hampel is an employee of Eisai Inc. During part of this work he was supported by the AXA Research Fund, the "Fondation partenariale Sorbonne Université" and the "Fondation pour la Recherche sur Alzheimer", Paris, France. Mannheim, Germany Biobank: Department of geriatric Psychiatry,

Central Institute for Mental Health, Mannheim, University of Heidelberg, Germany. Genotyping for the Swedish Twin Studies of Aging was supported by NIH/NIA grant R01 AG037985. Genotyping in TwinGene was supported by NIH/NIDDK U01 DK066134. WvdF is recipient of Joint Programming for Neurodegenerative Diseases (JPND) grants PERADES (ANR-13-JPRF-0001) and EADB (733051061). Gothenburg Birth Cohort (GBC) Studies: We would like to thank UCL Genomics for performing the genotyping analyses. The studies were supported by The Stena Foundation, The Swedish Research Council (2015-02830, 2013-8717), The Swedish Research Council for Health, Working Life and Wellfare (2013-1202, 2005-0762, 2008-1210, 2013-2300, 2013- 2496, 2013-0475), The Brain Foundation, Sahlgrenska University Hospital (ALF), The Alzheimer's Association (IIRG-03-6168), The Alzheimer's Association Zenith Award (ZEN-01-3151), Eivind och Elsa K:son Sylvans Stiftelse, The Swedish Alzheimer Foundation. Clinical AD, Sweden: We would like to thank UCL Genomics for performing the genotyping analyses. Barcelona Brain Biobank: Brain Donors of the Neurological Tissue Bank of the Biobanc-Hospital Clinic-IDIBAPS and their families for their generosity. We are indebted to the Biobanc-Hospital Clinic-IDIBAPS for samples and data procurement. Hospital Clínic de Barcelona Spanish Ministry of Economy and Competitiveness-Instituto de Salud Carlos III and Fondo Europeo de Desarrollo Regional (FEDER), Unión Europea, "Una manera de hacer Europa" grants (PI16/0235 to Dr. R. Sánchez-Valle and PI17/00670 to Dr. A.Antonelli). AA is funded by Departament de Salut de la Generalitat de Catalunya, PERIS 2016-2020 (SLT002/16/00329). Work at JP-T laboratory was possible thanks to funding from Ciberned and generous gifts from Consuelo Cervera Yuste and Juan Manuel Moreno Cervera. Sydney Memory and Ageing Study (Sydney MAS): We gratefully acknowledge and thank the following for their contributions to Sydney MAS: participants, their supporters and the Sydney MAS Research Team (current and former staff and students). Funding was awarded from the Australian National Health and Medical Research Council (NHMRC) Program Grants (350833, 568969, 109308). AddNeuroMed consortium was led by Simon Lovestone, Bruno Vellas, Patrizia Mecocci, Magda Tsolaki, Iwona Kłoszewska, Hilkka Soininen. This work was supported by InnoMed (Innovative Medicines in Europe), an integrated project funded by the European Union of the Sixth Framework program priority (FP6-2004- LIFESCIHEALTH-5). Oviedo: This work was partly supported by Grant from Fondo de Investigaciones Sanitarias-Fondos FEDER EuropeanUnion to Victoria Alvarez PI15/00878. Project MinE: The ProjectMinE study was supported by the ALS Foundation Netherlands and the MND association (UK) (Project MinE, www.projectmine.com). The SPIN cohort: We are indebted to patients and their families for their participation in the "Sant Pau Initiative on Neurodegeneration cohort", at the Sant Pau Hospital (Barcelona). This is a multimodal research cohort for biomarker discovery and validation that is partially funded by Generalitat de Catalunya (2017 SGR 547 to JC), as well as from the Institute of Health Carlos III-Subdirección General de Evaluación and the Fondo Europeo de Desarrollo Regional (FEDER-"Una manera de Hacer Europa") (grants PI11/02526, PI14/01126, and PI17/01019 to JF; PI17/01895 to AL), and the Centro de Investigación Biomédica en Red Enfermedades Neurodegenerativas programme (Program 1, Alzheimer Disease to AL). We would also like to thank the Fundació Bancària Obra Social La Caixa (DABNI project) to JF and AL; and Fundación BBVA (to AL), for their support in funding this follow-up study. Adolfo López de Munain is supported by Fundación Salud 2000 (PI2013156), CIBERNED and Diputación Foral de Gipuzkoa

(Exp.114/17). Pascual Sánchez-Juan is supported by CIBERNED and Carlos III Institute of Health, Spain (PI08/0139, PI12/02288, and PI16/01652, PI20/01011), jointly funded by Fondo Europeo de Desarrollo Regional (FEDER), Unión Europea, “Una manera de hacer Europa”. We thank Biobanco Valdecilla for their support. Amsterdam dementia Cohort (ADC): Research of the Alzheimer center Amsterdam is part of the neurodegeneration research program of Amsterdam Neuroscience. The AlzheimerCenter Amsterdam is supported by Stichting Alzheimer Nederland and Stichting VUmc fonds. The clinical database structure was developed with funding from Stichting Dioraphte. Genotyping of the Dutch case-control samples was performed in the context of EADB (European Alzheimer&Dementia biobank) funded by the JPco-fuND FP-829-029 (ZonMW project number #733051061). This research is performed by using data from the Parelsnoer Institute an initiative of the Dutch Federation of University Medical Centres (www.parelsnoer.org). 100-Plus study: We are grateful for the collaborative efforts of all participating centenarians and their family members and/or relations. We thank the Netherlands Brain Bank for supplying DNA for genotyping. This work was supported by Stichting AlzheimerNederland (WE09.2014-03), Stichting Diorapthe, Horstingstuit foundation, Memorabel (ZonMW project number #733050814, #733050512) and Stichting VUmcFonds. Additional support for EADB cohorts: WF, SL, HH are recipients of ABOARD, a public-private partnership receiving funding from ZonMW (#73305095007) and Health~Holland, Topsector Life Sciences & Health (PPP-allowance; #LSHM20106). The DELCODE study was funded by the German Center for Neurodegenerative Diseases (Deutsches Zentrum für Neurodegenerative Erkrankungen (DZNE)), reference number BN012. The Hellenic Longitudinal Investigation of Aging and Diet study was supported by Grant IIRG-09-133,014 from the Alzheimer’s Association; Grant 18910276/8/9/2011 from the European Social Fund; and Grant ΔΥ2β/οικ0.51657/14.4.2009 from the Ministry of Health and Social Solidarity (Greece).

**Acknowledgments for GR@ACE/DEGESCO**

We would like to thank patients, controls and researchers who participated in GR@ACE/DEGESCO project. I. de Rojas is supported by national grant from the Instituto de Salud Carlos III FI20/00215. The Genome Research @ Fundació ACE project (GR@ACE) is supported by Grifols SA, Fundación bancaria “La Caixa”, Fundació ACE, and CIBERNED. A.R. and M.B. receive support from the European Union/EFPIA Innovative Medicines Initiative Joint undertaking ADAPTED and MOPEAD projects (grant numbers 115975 and 115985, respectively). M.B. and A.R. are also supported by national grants PI13/02434, PI16/01861, PI17/01474, PI19/01240 and PI19/01301. Acción Estratégica en Salud is integrated into the Spanish National R+D+I Plan and funded by ISCIII (Instituto de Salud Carlos III)—Subdirección General de Evaluación and the Fondo Europeo de Desarrollo Regional (FEDER—“Una manera de hacer Europa”). Some control samples and data from patients included in this study were provided in part by the National DNA Bank Carlos III (www.bancoadn.org, University of Salamanca, Spain) and Hospital Universitario Virgen de Valme (Sevilla, Spain); they were processed following standard operating procedures with the appropriate approval of the Ethical and Scientific Committee.

**Acknowledgments for EADI**

This work has been developed and supported by the LABEX (laboratory of excellence program investment for the future) DISTALZ grant (Development of Innovative Strategies for a Transdisciplinary approach to ALZheimer's disease) including funding from MEL (Metropole européenne de Lille), ERDF (European Regional Development Fund) and Conseil Régional Nord Pas de Calais. This work was supported by INSERM, the National Foundation for Alzheimer's disease and related disorders, the Institut Pasteur de Lille and the Centre National de Recherche en Génomique Humaine, CEA, the JPND PERADES, the Laboratory of Excellence GENMED (Medical Genomics) grant no. ANR-10-LABX-0013 managed by the National Research Agency (ANR) part of the Investment for the Future program, and the FP7 AgedBrainSysBio. The Three-City Study was performed as part of collaboration between the Institut National de la Santé et de la Recherche Médicale (Inserm), the Victor Segalen Bordeaux II University and Sanofi-Synthélabo. The Fondation pour la Recherche Médicale funded the preparation and initiation of the study. The 3C Study was also funded by the Caisse Nationale Maladie des Travailleurs Salariés, Direction Générale de la Santé, MGEN, Institut de la Longévité, Agence Française de Sécurité Sanitaire des Produits de Santé, the Aquitaine and Bourgogne Regional Councils, Agence Nationale de la Recherche, ANR supported the COGINUT and COVADIS projects. Fondation de France and the joint French Ministry of Research/INSERM "Cohortes et collections de données biologiques" programme. Lille Génopôle received an unconditional grant from Eisai. The Three-city biological bank was developed and maintained by the laboratory for genomic analysis LAG-BRC - Institut Pasteur de Lille.

**Acknowledgments for GERAD**

We thank all individuals who participated in this study. Cardiff University was supported by the Wellcome Trust, Alzheimer's Society (AS; grant RF014/164), the Medical Research Council (MRC; grants G0801418/1, MR/K013041/1, MR/L023784/1), the European Joint Programme for Neurodegenerative Disease (JPND, grant MR/L501517/1), Alzheimer's Research UK (ARUK, grant ARUK-PG2014-1), Welsh Assembly Government (grant SGR544:CADR), a donation from the Moondance Charitable Foundation, UK Dementia's Platform (DPUK, reference MR/L023784/1), and the UK Dementia Research Institute at Cardiff. Cambridge University acknowledges support from the MRC. ARUK supported sample collections at the Kings College London, the South West Dementia Bank, Universities of Cambridge, Nottingham, Manchester and Belfast. King's College London was supported by the NIHR Biomedical Research Centre for Mental Health and Biomedical Research Unit for Dementia at the South London and Maudsley NHS Foundation Trust and Kings College London and the MRC. Alzheimer's Research UK (ARUK) and the Big Lottery Fund provided support to Nottingham University. Ulster Garden Villages, AS, ARUK, American Federation for Aging Research, NI R&D Office and the Royal College of Physicians/Dunhill Medical Trust provided support for Queen's University, Belfast. The University of Southampton acknowledges support from the AS. The MRC and Mercer's Institute for Research on Ageing supported the Trinity College group. DCR is a Wellcome Trust Principal Research fellow. The South

West Dementia Brain Bank acknowledges support from Bristol Research into Alzheimer's and Care of the Elderly. The Charles Wolfson Charitable Trust supported the OPTIMA group. Washington University was funded by NIH grants, Barnes Jewish Foundation and the Charles and Joanne Knight Alzheimer's Research Initiative. Patient recruitment for the MRC Prion Unit/UCL Department of Neurodegenerative Disease collection was supported by the UCLH/UCL Biomedical Research Centre and their work was supported by the NIHR Queen Square Dementia BRU, the Alzheimer's Research UK and the Alzheimer's Society. LASER-AD was funded by Lundbeck SA. The AgeCoDe study group was supported by the German Federal Ministry for Education and Research grants 01 GI 0710, 01 GI 0712, 01 GI 0713, 01 GI 0714, 01 GI 0715, 01 GI 0716, 01 GI 0717. Genotyping of the Bonn case-control sample was funded by the German centre for Neurodegenerative Diseases (DZNE), Germany. The GERAD Consortium also used samples ascertained by the NIMH AD Genetics Initiative. HH was supported by a grant of the Katharina-Hardt-Foundation, Bad Homburg vor der Höhe, Germany. The KORA F4 studies were financed by Helmholtz Zentrum München; German Research Center for Environmental Health; BMBF; German National Genome Research Network and the Munich Center of Health Sciences. The Heinz Nixdorf Recall cohort was funded by the Heinz Nixdorf Foundation (Dr. Jur. G.Schmidt, Chairman) and BMBF. We acknowledge use of genotype data from the 1958 Birth Cohort collection and National Blood Service, funded by the MRC and the Wellcome Trust which was genotyped by the Wellcome Trust Case Control Consortium and the Type-1 Diabetes Genetics Consortium, sponsored by the National Institute of Diabetes and Digestive and Kidney Diseases, National Institute of Allergy and Infectious Diseases, National Human Genome Research Institute, National Institute of Child Health and Human Development and Juvenile Diabetes Research Foundation International. The project is also supported through the following funding organisations under the aegis of JPND - www.jpnd.eu (United Kingdom, Medical Research Council (MR/L501529/1; MR/R024804/1) and Economic and Social Research Council (ES/L008238/1)) and through the Motor Neurone Disease Association. This study represents independent research part funded by the National Institute for Health Research (NIHR) Biomedical Research Centre at South London and Maudsley NHS Foundation Trust and King's College London. Prof. Jens Wiltfang is supported is supported by an Ilídio Pinho professorship, iBiMED (UIDB/04501/2020) at the University of Aveiro, Portugal.

**Acknowledgments for DemGene**

The project has received funding from The Research Council of Norway (RCN) Grant Nos. 213837, 223273, 225989, 248778, and 251134 and EU JPND Program RCN Grant Nos. 237250, 311993, the South-East Norway Health Authority Grant No. 2013-123, the Norwegian Health Association, and KG Jebsen Foundation. The RCN FRIPRO Mobility grant scheme (FRICON) is co-funded by the European Union's Seventh Framework Programme for research, technological development and demonstration under Marie Curie grant agreement No 608695. European Community's grant PIAPP-GA-2011-286213 PsychDPC.

**Acknowledgments for other GWAS and phenotype data**

The NACC database is funded by NIA/NIH Grant U01 AG016976. NACC data are contributed by the NIA-funded ADCs: P30 AG019610 (PI Eric Reiman, MD), P30 AG013846 (PI Neil Kowall, MD), P30 AG062428-01 (PI James Leverenz, MD) P50 AG008702 (PI Scott Small, MD), P50 AG025688 (PI Allan Levey, MD, PhD), P50 AG047266 (PI Todd Golde, MD, PhD), P30 AG010133 (PI Andrew Saykin, PsyD), P50 AG005146 (PI Marilyn Albert, PhD), P30 AG062421-01 (PI Bradley Hyman, MD, PhD), P30 AG062422-01 (PI Ronald Petersen, MD, PhD), P50 AG005138 (PI Mary Sano, PhD), P30 AG008051 (PI Thomas Wisniewski, MD), P30 AG013854 (PI Robert Vassar, PhD), P30 AG008017 (PI Jeffrey Kaye, MD), P30 AG010161 (PI David Bennett, MD), P50 AG047366 (PI Victor Henderson, MD, MS), P30 AG010129 (PI Charles DeCarli, MD), P50 AG016573 (PI Frank LaFerla, PhD), P30 AG062429-01(PI James Brewer, MD, PhD), P50 AG023501 (PI Bruce Miller, MD), P30 AG035982 (PI Russell Swerdlow, MD), P30 AG028383 (PI Linda Van Eldik, PhD), P30 AG053760 (PI Henry Paulson, MD, PhD), P30 AG010124 (PI John Trojanowski, MD, PhD), P50 AG005133 (PI Oscar Lopez, MD), P50 AG005142 (PI Helena Chui, MD), P30 AG012300 (PI Roger Rosenberg, MD), P30 AG049638 (PI Suzanne Craft, PhD), P50 AG005136 (PI Thomas Grabowski, MD), P30 AG062715-01 (PI Sanjay Asthana, MD, FRCP), P50 AG005681 (PI John Morris, MD), P50 AG047270 (PI Stephen Strittmatter, MD, PhD).
The genotypic and associated phenotypic data used in the study “Multi-Site Collaborative Study for Genotype-Phenotype Associations in Alzheimer’s Disease (GenADA)” were provided by the GlaxoSmithKline, R&D Limited.
ROSMAP study data were provided by the Rush Alzheimer’s Disease Center, Rush University Medical Center, Chicago. Data collection was supported through funding by NIA grants P30AG10161, R01AG15819, R01AG17917, R01AG30146, R01AG36836, U01AG32984, U01AG46152, the Illinois Department of Public Health, and the Translational Genomics Research Institute.
The AddNeuroMed data are from a public-private partnership supported by EFPIA companies and SMEs as part of InnoMed (Innovative Medicines in Europe), an Integrated Project funded by the European Union of the Sixth Framework program priority FP6-2004-LIFESCIHEALTH-5. Clinical leads responsible for data collection are Iwona Kłoszewska (Lodz), Simon Lovestone (London), Patrizia Mecocci (Perugia), Hilkka Soininen (Kuopio), Magda Tsolaki (Thessaloniki), and Bruno Vellas (Toulouse), imaging leads are Andy Simmons (London), Lars-Olad Wahlund (Stockholm) and Christian Spenger (Zurich) and bioinformatics leads are Richard Dobson (London) and Stephen Newhouse (London).

**Acknowledgement for the McGill cohort**

We would like to thank the participants. The access to part of the participants for this research has been made possible thanks to the Quebec Parkinson’s Network (http://rpq-qpn.ca/en/). Dr. Gan-Or is supported by the Fonds de recherche du Québec - Santé (FRQS) Chercheurs-boursiers award in collaboration with Parkinson Quebec, by the Young Investigator Award by Parkinson Canada, and is a William Dawson Scholar. This work was financially supported by grants from the Michael J. Fox Foundation, the Canadian Consortium on Neurodegeneration in Aging (CCNA), the Canada First Research Excellence Fund (CFREF), awarded to McGill University for the Healthy Brains for Healthy Lives initiative (HBHL), and Parkinson Canada.

### Consortia author list

#### EADB

Céline Bellenguez[1], Fahri Küçükali[2,3,4], Iris Jansen[5,6], Victor Andrade[7,8], Sonia Moreno-Grau[9,10], Najaf Amin[11,12], Benjamin Grenier-Boley[1], Rafael Campos-Martin[7], Peter A. Holmans[13], Anne Boland[14], Luca Kleineidam[7,8,15], Vincent Damotte[1], Sven J. van der Lee[5,16], Teemu Kuulasmaa[17], Itziar de Rojas[9,10], Amber Yaqub[11], Ivana Prokic[11], Marcos R, Costa[1,18], Julien Chapuis[1], Shahzad Ahmad[11,19], Vilmantas Giedraitis[20], Dag Aarsland[21,22], Pablo Garcia-Gonzalez[9,10], Carla Abdelnour[9,10], Emilio Alarcón-Martín[9,23], Daniel Alcolea[10,24], Montserrat Alegret[9,10], Ignacio Alvarez[25,26], Victoria Álvarez[27,28], Nicola J. Armstrong[29], Tsolaki Anthoula[30,31], Ildebrando Appollonio[32,33], Marina Arcaro[34], Silvana Archetti[35], Alfonso Arias Pastor[36,37], Beatrice Arosio[38,39], Lavinia Athanasiu[40], Henri Bailly[41], Nerisa Banaj[42], Miquel Baquero[43], Ana Belén Pastor[44], Luisa Benussi[45], Claudine Berr[46], Céline Besse[14], Valentina Bessi[47,48], Giuliano Binetti[45,49], Alessandra Bizarro[50], Rafael Blesa[10,24], Mercè Boada[9,10], Barbara Borroni[51], Silvia Boschi[52], Paola Bossù[53], Geir Bråthen[54,55], Catherine Bresner[13], Henry Brodaty[29,56], Keeley J. Brookes[57], Luis Ignacio Brusco[58,59,60], Dolores Buiza-Rueda[10,61], Katharina Bûrger[62,63], Vanessa Burholt[64,65], Miguel Calero[10,44,66], Geneviève Chene[67,68], Ángel Carracedo[69,70], Roberta Cecchetti[71], Laura Cervera-Carles[10,24], Camille Charbonnier[72], Caterina Chillotti[73], Simona Ciccone[39], Jurgen A.H.R. Claassen[74], Jordi Clarimon[10,24], Christopher Clark[75], Elisa Conti[32], Anaïs Corma-Gómez[76], Emanuele Costantini[77], Carlo Custodero[78], Delphine Daian[14], Maria Carolina Dalmasso[7], Antonio Daniele[77], Efthimios Dardiotis[79], Jean-François Dartigues[80], Peter Paul de Deyn[81], Stéphanie Debette[80,82], Jürgen Deckert[83], Teodoro del Ser[44], Nicola Denning[84], Martin Dichgans[62,63,85], Janine Diehl-Schmid[86], Mónica Diez-Fairen[25,26], Paolo Dionigi Rossi[39], Srdjan Djurovic[40], Emmanuelle Duron[41], Emrah Düzel[87,88], Carole Dufouil[67,68], Valentina Escott-Price[13,84], Ana Espinosa[9,10], Michael Ewers[62,63], Marta Fernández-Fuertes[76], Catarina B Ferreira[89], Evelyn Ferri[39], Bertrand Fin[14], Peter Fischer[90], Tormod Fladby[91], Klaus Fließbach[8,15], Juan Fortea[10,24], Silvia Fostinelli[45], Nick C. Fox[92], Emlio Franco-Macías[93], María J. Bullido[10,94,95], Ana Frank-García[10,94,96], Lutz Froelich[97], Daniela Galimberti[34,88], Jose Maria García-Alberca[10,98], Pablo García-González[9], Sebastian Garcia-Madrona[99], Guillermo Garcia-Ribas[99], Roberta Ghidoni[45], Ina Giegling[100], Giaccone Giorgio[85], Oliver Goldhardt[86], Antonio González-Pérez[101], Caroline Graff[102,118], Giulia Grande[103], Emma Green[104], Timo Grimmer[86], Edna Grünblatt[105,106,107], Tamar Guetta-Baranes[108], Annakaisa Haapasalo[109], Georgios Hadjigeorgiou[110], Harald Hampel[111,112], Olivier Hanon[41], John Hardy[113], Annette M. Hartmann[100], Lucrezia Hausner[97], Janet Harwood[13], Stefanie Heilmann-Heimbach[114], Seppo Helisalmi[115,116], Michael T. Heneka[8,16], Isabel Hernández[9,10], Martin J. Herrmann[83], Per Hoffmann[114], Clive Holmes[117], Henne Holstege[5,16], Raquel Huerto Vilas[36,37], Marc Hulsman[5,16], Charlotte Johansson[102,118], Lena Kilander[20], Anne Kinhult Ståhlbom[102,118], Miia Kivipelto[119,120,121,122], Anne Koivisto[115], Johannes Kornhuber[123], Mary H. Kosmidis[124], Carmen Lage[10,125], Erika J Laukka[103,126], Alessandra Lauria[50], Jenni Lehtisalo[115,127], Ondrej Lerch[128,129], Alberto Lleó[10,24], Adolfo Lopez de Munain[10,130], Malin Löwemark[20], Lauren Luckcuck[13], Juan Macías[76], Catherine A. MacLeod[131], Wolfgang Maier[8,15], Francesca Mangialasche[119], Spallazzi Marco[132], Marta Marquié[9,10], Rachel Marshall[13], Angel Martín Montes[10,94,96], Carmen Martínez Rodríguez[28], Carlo Masullo[133], Simon Mead[134], Patrizia Mecocci[71], Miguel Medina[10,44], Alun Meggy[84], Shima Mehrabian[135], Silvia Mendoza[98], Manuel Menéndez-González[28], Pablo Mir[10,61], Susanne Moebus[136], Merel Mol[137], Laura Molina-Porcel[138,139], Laura Montrreal[9], Laura Morelli[140], Fermin Moreno[10,130], Kevin Morgan[141], Markus M Möthen[114], Carolina Muchnik[58], Benedetta Nacmias[47,142], Tiia Ngandu[127], Gael Nicolas[72], Børge G. Nordestgaard[143,144], Robert Olaso[14], Adelina Orellana[9,10], Michela Orsini[77], Gemma Ortega[9,10],

Alessandro Padovani[51], Caffarra Paolo[145], Goran Papenberg[103], Lucilla Parnetti[87], Pau Pastor[25,26], Alba Pérez-Cordón[9], Jordi Pérez-Tur[10,146,147], Pierre Pericard[148], Oliver Peters[149,150], Yolande A.L. Pijnenburg[5], Juan A Pineda[76], Gerard Piñol-Ripoll[36,37], Claudia Pisanu[151], Thomas Polak[83], Julius Popp[152,153,154], Danielle Posthuma[6], Josef Priller[150,155], Raquel Puerta[9], Olivier Quenez[72], Inés Quintela[69], Jesper Qvist Thomassen[156], Alberto Rábano[10,44], Innocenzo Rainero[52], Inez Ramakers[157], Luis M Real[76,158], Marcel J.T. Reinders[159], Steffi Riedel-Heller[160], Peter Riederer[161], Natalia Roberto[9], Eloy Rodriguez-Rodriguez[10,125], Arvid Rongve[162,163], Irene Rosas Allende[27,28], Maitée Rosende-Roca[9,10], Jose Luis Royo[164], Elisa Rubino[165], Dan Rujescu[100], María Eugenia Sáez[101], Paraskevi Sakka[166], Ingvild Saltvedt[55,167], Ángela Sanabria[9,10], María Bernal Sánchez-Arjona[93], Florentino Sanchez-Garcia[168], Pascual Sánchez Juan[10,125], Raquel Sánchez-Valle[169], Sigrid B Sando[54,55], Michela Scamosci[71], Nikolaos Scarmeas[170,171], Elio Scarpini[34,88], Philip Scheltens[5], Norbert Scherbaum[172], Martin Scherer[173], Matthias Schmid[16,174], Anja Schneider[8,16], Jonathan M. Schott[92], Geir Selbæk[91,175], Davide Seripa[176], Alexey A Shadrin[40], Olivia Skrobot[119], Hilkka Soininen[115], Vincenzo Solfrizzi[78], Alina Solomon[115], Sandro Sorbi[47,142], Oscar Sotolongo-Grau[9], Gianfranco Spalletta[42], Annika Spottke[16], Alessio Squassina[177], Eystein Stordal[178], Juan Pablo Tartan[9], Lluís Tárraga[9,10], Niccolo Tesí[5,16], Anbupalam Thalamuthu[29], Tegos Thomas[30,31], Latchezar Traykov[135], Lucio Tremolizzo[32,33], Anne Tybjærg-Hansen[144,156], Andre Uitterlinden[179], Abbe Ullgren[102], Ingun Ulstein[175], Sergi Valero[9,10], Aad van der Lugt[180], Jasper Van Dongen[2,3,4], Jeroen van Rooij[137], John van Swieten[137], Rik Vandenberghe[181,182], Frans Verhey[157], Jean-Sébastien Vidal[41], Jonathan Vogelgsang[183,184], Martin Vyhnalek[128,129], Michael Wagner[8,16], David Wallon[185], Leonie Weinhold[174], Jens Wiltfang[183,186,187], Gill Windle[131], Bob Woods[131], Mary Yannakoulia[188], Miren Zulaica[10,189], Jan Laczo[128,129], Vaclav Matoska[190], Maria Serpente[88], Francesca Assogna[42], Fabrizio Piras[42], Federica Piras[42], Valentina Ciullo[42], Jacob Shofany[42], Carlo Ferrarese[32,33], Simona Andreoni[32], Gessica Sala[32], Chiara Paola Zoia[32], Maria Del Zompo[177], Alberto Benussi[51], Patrizia Bastiani[191], Mari Takalo*[192], Teemu Natunen*[192], Tiina Laatikainen[120,127], Jaakko Tuomilehto[120,127], Riitta Antikainen[193,194], Timo Strandberg[193,195], Jaana Lindström[127], Markku Peltonen[127], Richard Abraham[196], Ammar Al-Chalabi[197], Nicholas J. Bass[198], Carol Brayne[199], Kristelle S. Brown[200], John Collinge[201], David Craig[202], Pangiotis Deloukas[203], Nick Fox[204], Amy Gerrish[204], Michael Gill[205], Rhian Gwilliam[203], John Hardy[206], Denise Harold[207], Paul Hollingworth[196], Jarret A, Johnston[208], Lesley Jones[196], Brian Lawlor[205], Gill Livingston[198], Simon Lovestone[209], Michelle Lupton[210,211], Aoibhinn Lynch[205], David Mann[212], Bernadette McGuinness[208], Andrew McQuillin[198], Michael C. O'Donovan[196], Michael J. Owen[196], Peter Passmore[208], John F, Powell[210,211], Petra Proitsi[210,211], Martin Rossor[204], Christopher E. Shaw[197], A. David Smith[213], Hugh Gurling[214], Stephen Todd[215], Catherine Mummery[216], Nathalie Ryan[216], Giordano Lacidogna[77], Ad Adarmes-Gómez[10,61], Ana Mauleón[9], Ana Pancho[9], Anna Gailhajenet[9], Asunción Lafuente[9], D Macias-García[10,61], Elvira Martín[9], Esther Pelejà[9], F Carrillo[10,61], Isabel Sastre Merlín[10,95], L Garrote-Espina[10,61], Liliana Vargas[9], M Carrion-Claro[10,61], M Marín[93], Ma Labrador[10,61], Mar Buendia[9], María Dolores Alonso[217], Marina Guitart[9], Mariona Moreno[9], Marta Ibarria[9], Mt Periñán[10,61], Nuria Aguilera[9], P Gómez-Garre[10,61], Pilar Cañabate[9], R Escuela[10,61], R Pineda-Sánchez[10,61], R Vigo-Ortega[10,61], S Jesús[10,61], Silvia Preckler[9], Silvia Rodrigo-Herrero[93], Susana Diego[9], Alessandro Vacca[52], Fausto Roveta[52], Nicola Salvadori[87], Elena Chipi[87], Henning Boecker[15,218], Christoph Laske[219,220], Robert Perneczky[65,221], Costas Anastasiou[188], Daniel Janowitz[62], Rainer Malik[62], Anna Anastasiou[30], Kayenat Parveen[7], Carmen Lage[222], Sara López-García[222], Anna Antonell[169], Kalina Yonkova Mihova[223], Diyana Belezhanska[135], Heike Weber[224], Silvia Kochen[225], Patricia Solis[225], Nancy Medel[225], Julieta Lisso[225], Zulma Sevillano[225], Daniel G Politis[225,226], Valeria Cores[225,226], Carolina Cuesta[225,226], Cecilia Ortiz[227], Juan Ignacio Bacha[227], Mario Rios[228], Aldo Saenz[228], Mariana Sanchez Abalos[229], Eduardo Kohler[230], Dana Lis Palacio[231], Ignacio Etchepareborda[231], Matias Kohler[231], Gisela Novack[232], Federico Ariel Prestia[232], Pablo Galeano[232], Eduardo M. Castaño[232], Sandra Germani[233], Carlos Reyes Toso[233], Matias Rojo[233],

Carlos Ingino[233], Carlos Mangone[233], Sebastiaan Engelborghs[234,235,236,237], Tagliavini Fabrizio[238], Sune Fallgaard Nielsen[239], Lucia Farotti[240], Chiara Fenoglio[241], Geert Jan Biessels[242], Seth Love[243], Patrick G. Kehoe[243], Florence Pasquier[244], Christine Van Broeckhoven[2,3,245], David C. Rubinsztein[246], Stefan Teipel[247], Nathalie Fievet[1], Vincent Deramecourt[244], Charlotte Forsell[102,118], Håkan Thonberg[102,118], Maria Bjerke[69], Ellen De Roeck[69], María Teresa Martínez-Larrad[2248], Natividad Olivar[233], Mohsen Ghanbari[11], Perminder Sachdev[29], Karen Mather[29], Frank Jessen[8,16], M. Arfan Ikram[11], Alexandre de Mendonça[89], Jakub Hort[128,129], Tsolaki Magda[30,31], Philippe Amouyel[1], Julie Williams[13], Ruth Frikke-Schmidt[144,156], Jordi Clarimon[10,24], Jean-François Deleuze[14], Giacomina Rossi[85], Ole A. Andreassen[40], Martin Ingelsson[20], Mikko Hiltunen[17], Kristel Sleegers[2,3,4], Cornelia M. van Duijn[11,12], Rebecca Sims[13], Wiesje M. van der Flier[5], Agustín Ruiz[9,10], Alfredo Ramirez[7,8,16,249], Jean-Charles Lambert[1]

1. Univ. Lille, Inserm, CHU Lille, Institut Pasteur de Lille, U1167-RID-AGE facteurs de risque et déterminants moléculaires des maladies liés au vieillissement, Lille, France
2. Complex Genetics of Alzheimer's Disease Group, VIB Center for Molecular Neurology, VIB, Antwerp, Belgium
3. Laboratory of Neurogenetics, Institute Born - Bunge, Antwerp, Belgium
4. Department of Biomedical Sciences, University of Antwerp, Neurodegenerative Brain Diseases Group,
5. Alzheimer Center Amsterdam, Department of Neurology, Amsterdam Neuroscience, Vrije Universiteit Amsterdam, Amsterdam UMC, Amsterdam, The Netherlands
6. Department of Complex Trait Genetics, Center for Neurogenomics and Cognitive Research, Amsterdam, The Netherlands
7. Division of Neurogenetics and Molecular Psychiatry, Department of Psychiatry and Psychotherapy, University of Cologne, Medical Faculty, Cologne, Germany.
8. Department of Neurodegenerative Diseases and Geriatric Psychiatry, University Hospital Bonn, Bonn, Germany
9. Research Center and Memory clinic Fundació ACE, Institut Català de Neurociències Aplicades, Universitat Internacional de Catalunya, Barcelona, Spain
10. CIBERNED, Network Center for Biomedical Research in Neurodegenerative Diseases, National Institute of Health Carlos III, Madrid, Spain
11. Department of Epidemiology, ErasmusMC, Totterdam, The Netherlands
12. Nuffield Department of Population Health Oxford University, Oxford, UK
13. MRC Centre for Neuropsychiatric Genetics and Genomics, , Division of Psychological Medicine and Clinical
14. Université Paris-Saclay, CEA, Centre National de Recherche en Génomique Humaine, 91057, Evry, France
15. German Center for Neurodegenerative Diseases (DZNE Bonn), Bonn, Germany
16. Section Genomics of Neurdegenerative Diseases and Aging, Department of Human Genetics Amsterdam, The Netherlands
17. Institute of Biomedicine, University of Eastern Finland, Kuopio, Finland
18. Brain Institute, Federal University of Rio Grande do Norte, Av. Nascimento de Castro 2155 Natal, Brazil
19. LACDR, Leiden, The Netherlands
20. Dept.of Public Health and Carins Sciences / Geriatrics, Uppsala University, Sweden
21. Centre of Age-Related Medicine, Stavanger University Hospital, Norway
22. Institute of Psychiatry, Psychology & Neuroscience, PO 70, 16 De Crespigny Park, London, UK
23. Department of Surgery, Biochemistry and Molecular Biology, School of Medicine, University of Málaga, Málaga, Spain.
24. Department of Neurology, II B Sant Pau, Hospital de la Santa Creu i Sant Pau, Universitat Autònoma de Barcelona, Barcelona, Spain.
25. Fundació Docència i Recerca MútuaTerrassa and Movement Disorders Unit, Department of

Neurology, University Hospital MútuaTerrassa, Terrassa 08221, Barcelona, Spain
26. Memory Disorders Unit, Department of Neurology, Hospital Universitari Mutua de Terrassa, Terrassa, Barcelona, Spain.
27. Laboratorio de Genética. Hospital Universitario Central de Asturias, Oviedo, Spain
28. Servicio de Neurología HOspital Universitario Central de Asturias- Oviedo and Instituto de Investigación Biosanitaria del Principado de Asturias, Oviedo, Spain
29. Centre for Healthy Brain Ageing, School of Psychiatry, Faculty of Medicine, University of New South Wales, Sydney, Australia
30. 1st Department of Neurology, Medical school, Aristotle University of Thessaloniki, Thessaloniki, Makedonia, Greece
31. Alzheimer Hellas, Thessaloniki, Makedonia, Greece
32. School of Medicine and Surgery, University of Milano-Bicocca, Italy
33. Neurology Unit, "San Gerardo" hospital, Monza, Italy
34. Fondazione IRCCS Ca' Granda, Ospedale Policlinico, Milan, Italy
35. Department of Laboratory Diagnostics, III Laboratory of Analysis, Brescia Hospital, Brescia, Italy
36. Unitat Trastorns Cognitius, Hospital Universitari Santa Maria de Lleida, Lleida, Spain
37. Institut de Recerca Biomedica de Lleida (IRBLLeida), Lleida, Spain
38. Department of Clinical Sciences and Community Health, University of Milan, Italy
39. Geriatic Unit, Fondazione Cà Granda, IRCCS Ospedale Maggiore Policlinico, Milan, Italy
40. NORMENT Centre, University of Oslo, Oslo, Norway
41. Université de Paris, EA 4468, APHP, Hôpital Broca, Paris, France
42. Laboratory of Neuropsychiatry, Department of Clinical and Behavioral Neurology, IRCCS Santa Lucia Foundation, Rome, Italy
43. Servei de Neurologia, Hospital Universitari i Politècnic La Fe, Valencia, Spain.
44. CIEN Foundation/Queen Sofia Foundation Alzheimer Center, Madrid, Spain
45. Molecular Markers Laboratory, IRCCS Istituto Centro San Giovanni di Dio Fatebenefratelli, Brescia, Italy
46. Univ. Montpellier, Inserm U1061, Neuropsychiatry: epidemiological and clinical research, PSNREC, Montpellier, France
47. Department of Neuroscience, Psychology, Drug Research and Child Health University of Florence, Florence Italy
48. Azienda Ospedaliero-Universitaria Careggi, Florence, Italy
49. MAC - Memory Clinic, IRCCS Istituto Centro San Giovanni di Dio Fatebenefratelli, Brescia
50. Geriatrics Unit Fondazione Policlinico A. Gemelli IRCCS, Rome , Italy
51. Centre for Neurodegenerative Disorders, Department of Clinical and Experimental Sciences, University of Brescia, Brescia, Italy
52. Department of Neuroscience “Rita Levi Montalcini”, University of Torino, Torino, Italy
53. Experimental Neuro-psychobiology Laboratory,Department of Clinical and Behavioral Neurology, IRCCS Santa Lucia Foundation, Rome, Italy
54. Department of Neurology and Clinical Neurophysiology, University Hospital of Trondheim, Trondheim, Norway
55. Department of Neuromedicine and Movement Science, Norwegian University of Science and Technology, Trondheim, Norway
56. Dementia Centre for Research Collaboration, School of Psychiatry, University of New South Wales, Sydney, Australia
57. Biosciences, School of Science and Technology, Nottingham Trent University, Nottingham UK
58. Centro de Neuropsiquiatría y Neurología de la Conducta (CENECON), Facultad de Medicina, Universidad de Buenos Aires (UBA), C.A.B.A, Buenos Aires, Argentina.
59. Departamento Ciencias Fisiológicas UAII, Facultad de Medicina, UBA, C.A.B.A, Buenos Aires, Argentina.
60. Hospital Interzonal General de Agudos Eva Perón, San Martín, Buenos Aires, Argentina.
61. Unidad de Trastornos del Movimiento, Servicio de Neurología y Neurofisiología. Instituto de Biomedicina de Sevilla (IBiS), Hospital Universitario Virgen del Rocío/CSIC/Universidad de

Sevilla, Seville, Spain
62. Institute for Stroke and Dementia Research, Klinikum der Universität München, Ludwig-Maximilians-Universität LMU, Munich, Germany.
63. German Center for Neurodegenerative Diseases (DZNE, Munich), Munich, Germany.
64. Faculty of Medical & Health Sciences, University of Auckland, New Zealand
65. Wales Centre for Ageing & Dementia Research, Swansea University, Wales, New Zealand
66. UFIEC, Instituto de Salud Carlos III, , Madrid, Spain
67. Inserm, Bordeaux Population Health Research Center, UMR 1219, Univ. Bordeaux, ISPED, CIC 1401-EC, Univ Bordeaux,Bordeaux, France
68. CHU de Bordeaux, Pole santé publique, Bordeaux, France
69. Grupo de Medicina Xenómica, Centro Nacional de Genotipado (CEGEN-PRB3-ISCIII). Universidade de Santiago de Compostela, Santiago de Compostela, Spain.
70. Fundación Pública Galega de Medicina Xenómica- CIBERER-IDIS, University of Santiago de Compostela, Santiago de Compostela, Spain.
71. Institute of Gerontology and Geriatrics, Department of Medicine and Surgery, University of Perugia Perugia, Italy
72. Normandie Univ, UNIROUEN, Inserm U1245 and CHU Rouen, Department of Genetics and CNR-MAJ, Rouen, France
73. Unit of Clinical Pharmacology, University Hospital of Cagliari, Cagliari, Italy
74. Radboudumc Alzheimer Center, Department of Geriatrics, Radboud University Medical Center, Nijmegen, the Netherlands
75. Institute for Regenerative Medicine, University of Zürich, Schlieren, Switzerland
76. Unidad Clínica de Enfermedades Infecciosas y Microbiología. Hospital Universitario de Valme, Sevilla, Spain
77. Department of Neuroscience, Catholic University of Sacred Heart, Fondazione Policlinico Universitario A. Gemelli IRCCS, Rome, Italy
78. University of Bari, “A. Moro”, Bari, Italy
79. School of Medicine, University of Thessaly, Larissa, Greece
80. University Bordeaux, Inserm, Bordeaux Population Health Research Center, France
81. Department of Neurology, University Medical Center Groningen, the Netherlands
82. Department of Neurology, Bordeaux University Hospital, Bordeaux, France
83. Department of Psychiatry, Psychosomatics and Psychotherapy, Center of Mental Health, University Hospital, Wuerzburg
84. UKDRI@ Cardiff, School of Medicine, Cardiff University, Cardiff, UK
85. Munich Cluster for Systems Neurology (SyNergy), Munich, Germany.
86. Technical University of Munich, School of Medicine, Klinikum rechts der Isar, Department of Psychiatry and Psychotherapy, Munich, Germany
87. Institute of Cognitive Neurology and Dementia Research (IKND), Otto-Von-Guericke University, Magdeburg, Germany.
88. German Center for Neurodegenerative Diseases (DZNE), Magdeburg, Germany.
89. Faculty of Medicine, University of Lisbon, Portugal
90. Department of Psychiatry, Social Medicine Center East- Donauspital, Vienna, Austria
91. Institute of Clinical Medicine, University of Oslo, Oslo, Norway.
92. Dementia Research Centre, UCL Queen Square Institute of Neurology, London, United Kingdom
93. Unidad de Demencias, Servicio de Neurología y Neurofisiología. Instituto de Biomedicina de Sevilla (IBiS), Hospital Universitario Virgen del Rocío/CSIC/Universidad de Sevilla, Seville, Spain
94. Instituto de Investigacion Sanitaria ‘Hospital la Paz’ (IdIPaz), Madrid, Spain
95. Centro de Biología Molecular Severo Ochoa (UAM-CSIC), Madrid, Spain
96. Hospital Universitario la Paz, Madrid, Spain
97. Department of geriatric Psychiatry, Central Institute for Mental Health, Mannheim, University of Heidelberg, Germany
98. Alzheimer Research Center & Memory Clinic, Andalusian Institute for Neuroscience, Málaga, Spain.

99. Hospital Universitario Ramon y Cajal, IRYCIS, Madrid
100. Department of Psychiatry and Psychotherapy, Medical University of Vienna, Vienna, Austria
101. CAEBI, Centro Andaluz de Estudios Bioinformáticos, Sevilla, Spain.
102. Karolinska Institutet, Center for Alzheimer Research, Department NVS, Division of Neurogeriatrics, Stockholm, Sweden
103. Aging Research Center, Department of Neurobiology, Care Sciences and Society, Karolinska Institutet and Stockholm University, Stockholm, Sweden
104. Institute of Public Health, University of Cambridge, UK
105. Department of Child and Adolescent Psychiatry and Psychotherapy, University Hospital of Psychiatry Zurich, University of Zurich, Zurich, Switzerland
106. Neuroscience Center Zurich, University of Zurich and ETH Zurich, Switzerland
107. Zurich Center for Integrative Human Physiology, University of Zurich, Switzerland
108. Human Genetics, School of Life Sciences, Life Sciences Building, University Park, University of Nottingham, Nottingham, UK
109. A.I Virtanen Institute for Molecular Sciences, University of Eastern Finland, Kuopio, Finland
110. Department of Neurology, Medical School, University of Cyprus, Cyprus
111. Sorbonne University, GRC n° 21, Alzheimer Precision Medicine Initiative (APMI), AP-HP, Pitié-Salpêtrière Hospital, Boulevard de l'hôpital, Paris, France
112. Eisai Inc., Neurology Business Group, 100 Tice Blvd, Woodcliff Lake, NJ 07677, USA
113. Reta Lila Weston Research Laboratories, Department of Molecular Neuroscience, UCL Institute of Neurology, London, UK.
114. Institute of Human Genetics, University of Bonn, School of Medicine & University Hospital Bonn, Bonn, Germany
115. Insitute of Clinical Medicine - Neurology, University of Eastern, Kuopio, Finland
116. Institute of Clinical Medicine – Internal Medicine, University of Eastern Finland, Kuopio, Finland
117. Clinical and Experimental Science, Faculty of Medicine, University of Southampton, Southampton, UK.
118. Unit for Hereditary dementias, Karolinska University Hospital-Solna, Stockholm, Sweden
119. Division of Clinical Geriatrics, Center for Alzheimer Research, Care Sciences and Society (NVS), Karolinska Institutet, Stockholm, Sweden
120. Institute of Public Health and Clinical Nutrition, University of Eastern Finland, Kuopio, Finland
121. Neuroepidemiology and Ageing Research Unit, School of Public Health, Imperial College London, London, United Kingdom
122. Stockholms Sjukhem, Research & Development Unit, Stockholm, Sweden
123. Department of Psychiatry and Psychotherapy, Universitätsklinikum Erlangen, and Friedrich-Alexander Universität Erlangen-Nürnberg, Erlangen, Germany.
124. Laboratory of Cognitive Neuroscience, School of Psychology, Aristotle University of Thessaloniki, Thessaloniki, Greece
125. Neurology Service, Marqués de Valdecilla University Hospital (University of Cantabria and IDIVAL), Santander, Spain.
126. Stockholm Gerontology Research Center, Stockholm, Sweden
127. Public Health Promotion Unit, Finnish Institute for Health and Welfare, Helsinki, Finland
128. Memory Clinic, Department of Neurology, Charles University, 2nd Faculty of Medicine and Motol University Hospital, Czech Republic
129. International Clinical Research Center, St. Anne's University Hospital Brno, Brno, Czech Republic
130. Department of Neurology. Hospital Universitario Donostia. OSAKIDETZA-Servicio Vasco de Salud, San Sebastian, Spain
131. School of Health Sciences, Bangor University, UK
132. Unit of Neurology, University of Parma and AOU, Parma, Italy
133. Institute of Neurology, Catholic University of the Sacred Heart , Rome, Itlay
134. MRC Prion Unit at UCL, UCL Institute of Prion Diseases, London, UK
135. Clinic of Neurology, UH "Alexandrovska", Medical University - Sofia, Sofia, Bulgaria
136. Institute for Urban Public Health, University Hospital of University Duisburg-Essen, Essen,

Germany
137. Department of Neurology, ErasmusMC, Rotterdam, The Netherlands
138. Neurological Tissue Bank of the Biobanc-Hospital Clinic-IDIBAPS, Institut d'Investigacions Biomèdiques August Pi i Sunyer, Barcelona, Spain.
139. Alzheimer's disease and other cognitive disorders Unit. Neurology Department, Hospital Clinic , Barcelona, Spain
140. Laboratory of Brain Aging and Neurodegeneration- FIL-CONICET, Buenos Aires, Argentina
141. Human Genetics, School of Life Sciences, University of Nottingham, UK
142. IRCCS Fondazione Don Carlo Gnocchi, Florence, Italy
143. Department of Clinical Biochemistry, Herlev and Gentofte Hospital, Herlev, Denmark
144. Department of Clinical Medicine, University of Copenhagen, Copenhagen, Denmark
145. DIMEC, University of Parma, Parma, Italy
146. Institut de Biomedicina de València-CSIC (valència, Spain) CIBERNED.
147. Unitat Mixta de de Neurología y Genética, Institut d'Investigació Sanitària La Fe (València, Spain)
148. Univ. Lille, CNRS, Inserm, CHU Lille, Institut Pasteur de Lille, US 41-UMS 2014-PLBS, bilille, Lille, France.
149. Institute of Psychiatry and Psychotherapy, Charité-Universitätsmedizin Berlin, Corporate Member of Freie Universität Berlin, Humboldt-Universität Zu Berlin, and Berlin Institute of Health, Berlin, Germany.
150. German Center for Neurodegenerative Diseases (DZNE), Berlin, Germany.
151. Department of Biomedical Sciences, University of Cagliari, Italy
152. CHUV, Old Age Psychiatry, Department of Psychiatry, Lausanne, Switzerland
153. Old Age Psychiatry, Department of Psychiatry, Lausanne University Hospital, Lausanne, Switzerland
154. Department of Geriatric Psychiatry, University Hospital of Psychiatry Zürich, Zürich, Switzerland
155. Department of Neuropsychiatry and Laboratory of Molecular Psychiatry, Charité, Charitéplatz 1, 10117 Berlin, Germany
156. Department of Clinical Biochemistry, Rigshospitalet, Copenhagen, Denmark
157. Maastricht University, Department of Psychiatry & Neuropsychologie, Alzheimer Center Limburg, Maastricht, the Netherlands
158. Depatamento de Especialidades Quirúrgicas, Bioquímica e Inmunología. Facultad de Medicina. Universidad de Málaga. Málaga, Spain
159. Delft Bioinformatics Lab, Delft University of Technology, Delft, The Netherlands
160. Institute of Social Medicine, Occupational Health and Public Health, University of Leipzig, 04103 Leipzig, Germany.
161. Center of Mental Health, Clinic and Policlinic of Psychiatry, Psychosomatics and Psychotherapy, University Hospital of Würzburg, Wuerzburg, Germany
162. Department of Research and Innovation, Helse Fonna, Haugesund Hospital, Haugesund, Norway.
163. The University of Bergen, Institute of Clinical Medicine (K1), Bergen Norway
164. Departamento de Especialidades Quirúrgicas, Bioquímicas e Inmunología, School of Medicine, University of Málaga, Málaga, Spain.
165. Department of Neuroscience and Mental Health, AOU Città della Salute e della Scienza di Torino, Torino, Italy
166. Athens Association of Alzheimer's disease and Related Disorders, Athens, Greece
167. Department of Geriatrics, St. Olav's Hospital, Trondheim University Hospital, Norway
168. Department of Immunology, Hospital Universitario Doctor Negrín, Las Palmas de Gran Canaria, Spain.
169. Neurology department-Hospital Clínic, IDIBAPS, Universitat de Barcelona, Barcelona, Spain.
170. Taub Institute for Research in Alzheimer's Disease and the Aging Brain, The Gertrude H. Sergievsky Center, Depatment of Neurology, Columbia University, New York, NY
171. 1st Department of Neurology, Aiginition Hospital, National and Kapodistrian University of Athens, Medical School, Greece

172. LVR-Hospital Essen, Department of Psychiatry and Psychotherapy, Medical Faculty, University of Duisburg-Essen, Virchowstr. 174, 45147 Essen, Germany
173. Department of Primary Medical Care, University Medical Centre Hamburg-Eppendorf, 20246 Hamburg, Germany.
174. Institute of Medical Biometry, Informatics and Epidemiology, University Hospital of Bonn, Bonn, Germany.
175. Department of Geriatric Medicine, Oslo University Hospital, Oslo, Norway
176. Laboratory for Advanced Hematological Diagnostics, Department of Hematology and Stem Cell Transplant, Lecce, Italy
177. Department of Biomedical Sciences, Section of Neuroscience and Clinical Pharmacology, University of Cagliari, Italy
178. Department of Psychiatry, Namsos Hospital, Namsos, Norway
179. Department of Internal medicine and Biostatistics, ErasmusMC, Rooterdam, The Netherlands
180. Department of Radiology&Nuclear medicine, ErasmusMC, Totterdam, The Netherlands
181. Laboratory for Cognitive Neurology, Department of Neurosciences, University of Leuven, Belgium
182. Neurology Department, University Hospitals Leuven, Leuven, Belgium
183. Department of Psychiatry and Psychotherapy, University Medical Center Goettingen, Goettingen, Germany
184. Department of Psychiatry, Harvard Medical School, McLean Hospital, Belmont, MA, USA
185. Normandie Univ, UNIROUEN, Inserm U1245, CHU Rouen, Department of Neurology and CNR-MAJ, F 76000, Normandy Center for Genomic and Personalized Medicine, Rouen, France
186. German Center for Neurodegenerative Diseases (DZNE), Goettingen, Germany
187. Medical Science Department, iBiMED, Aveiro, Portugal
188. Department of Nutrition and Diatetics, Harokopio University, Athens, Greece
189. Neurosciences Area. Instituto Biodonostia. San Sebastian, Spain
190. Department of Clinical Biochemistry, Hematology and Immunology, Na Homolce Hospital, Prague, Czech republic
191. Institute of Gerontology and Geriatrics, Department of Medicine, University of Perugia Perugia (Italy)
192. Insitute of Biomedicine, University of Eastern Finland, Finland
193. Center for Life Course Health Research, University of Oulu, Oulu, Finland
194. Medical Research Center Oulu, Oulu University Hospital, Oulu, Finland
195. University of Helsinki and Helsinki University Hospital, Helsinki, Finland
196. Division of Psychological Medicine and Clinial Neurosciences, MRC Centre for Neuropsychiatric Genetics and Genomics, Cardiff University, UK
197. Kings College London, Institute of Psychiatry, Psychology and Neuroscience, UK
198. Division of Psychiatry, University College London, UK
199. Institute of Public Health, University of Cambridge, Cambridge, UK
200. Institute of Genetics, Queens Medical Centre, University of Nottingham, Nottingham, UK
201. XXX
202. Ageing Group, Centre for Public Health, School of Medicine, Dentistry and Biomedical Sciences, Queen's University Belfast, UK
203. The Wellcome Trust Sanger Institute, Wellcome Trust Genome Campus, Hinxton, Cambridge, UK.
204. Dementia Research Centre, Department of Neurodegenerative Disease, UCL Institute of Neurology, London, UK
205. Mercer's Institute for Research on Ageing, St James' Hospital, Dublin, Ireland
206. Department of Molecular Neuroscience, UCL, Institute of Neurology, London, UK
207. School of Biotechnology, Dublin City University, Dublin, Ireland
208. Centre for Public Health, School of Medicine, Dentistry and Biomedical Sciences, Queens University, Belfast, UK
209. Department of Psychiatry, University of Oxford, Oxford, UK
210. Department of Basic and Clinical Neuroscience, Institute of Psychiatry, Psychology and

Neuroscience, Kings College London, London UK
211. Genetic Epidemiology, QIMR Berghofer Medical Research Institute, Herston, Queensland, Australia
212. Division of Neuroscience and Experimental Psychology, School of Biological Sciences, Faculty of Biology, Medicine and Health, University of Manchester, Manchester Academic Health Science Centre, Manchester M13 9PT, UK
213. Oxford Project to Investigate Memory and Ageing (OPTIMA), University of Oxford, Level 4, John Radcliffe Hospital, Oxford, UK
214. Department of Mental Health Sciences, University College London, London, UK
215. Ageing Group, Centre for Public Health, School of Medicine, Dentistry and Biomedical Sciences, Queen's University Belfast, Belfast, UK.
216. Dementia Research Centre, UCL, London, UK
217. Servei de Neurologia. Hospital Clínic Universitari de València, Spain
218. Department of Radiology, University Hospital Bonn, Bonn, Germany
219. German Center for Neurodegenerative Diseases (DZNE), Tübingen, Germany
220. Section for Dementia Research, Hertie Institute for Clinical Brain Research and Department of Psychiatry, Tübingen, Germany
221. Department of Psychiatry and Psychotherapy, University Hospital, LMU Munich, Munich, Germany
222. Service of Neurology, University Hospital Marqués de Valdecilla, IDIVAL, University of Cantabria, Santander, Spain
223. Molecular Medicine Center, Department of Medical chemistry and biochemistry, Medical University of Sofia, Bulgaria
224. Department of Psychiatry, Psychosomatics and Psychotherapy, Center of Mental Health, University Hospital of Würzburg, Germany
225. ENYS (Estudio en Neurociencias y Sistemas Complejos) CONICET- Hospital El Cruce "Nestor Kirchner"- UNAJ, Argentina
226. HIGA Eva Perón, Buenes Aires, Agentina
227. Neurología Clinica, Buenes Aires, Agentina
228. Dirección de Atención de Adultos Mayores del Min. Salud Desarrollo Social y Deportes de la Pcia. de Mendoza, Argentina
229. Laboratorio de Genética Forense del Ministerio Público de la Pcia. de La Pampa, Argentina
230. Fundacion Sinapsis, Santa Rosa, Argentina
231. Hospital Dr. Lucio Molas, Santa Rosa; Fundacion Ayuda Enfermo Renal y Alta Complejidad (FERNAC), Santa Rosa, Argentina
232. Laboratory of Brain Aging and Neurodegeneration- FIL, Buneos Aires, Argentina
233. Centro de Neuropsiquiatría y Neurología de la Conducta (CENECON), Facultad de Medicina, Universidad de Buenos Aires (UBA), C.A.B.A, Buenos Aires, Argentina
234. Center for Neurosciences, Vrije Universiteit Brussel (VUB), Brussels, Belgium
235. Reference Center for Biological Markers of Dementia (BIODEM), Institute Born-Bunge, University of Antwerp, Antwerp, Belgium
236. Institute Born-Bunge, University of Antwerp, Antwerp, Belgium
237. Department of Neurology, UZ Brussel, Brussels, Belgium
238. Fondazione IRCCS, Istituto Neurologico Carlo Besta, Milan Italy
239. Department of Clinical Biochemistry, Herlev and Gentofte Hospital, Herlev Denmark
240. Centre for Memory Disturbances, Lab of Clinical Neurochemistry, Section of Neurology, University of Perugia, Italy
241. University of Milan, Milan, Italy
242. Department of Neurology, UMC Utrecht Brain Center, Utrecht, the Netherlands
243. Translational Health Sciences, Bristol Medical School, University of Bristol, Bristol, BS16 1LE, UK
244. Univ Lille Inserm 1172, CHU Clinical and Research Memory Research Centre (CMRR) of Distalz, Licend, Lille France
245. Neurodegenerative Brain Diseases Group, VIB Center for Molecular Neurology, VIB, Antwerp,

Belgium
246. Cambridge Institute for Medical Research and UK Dementia Research Institute, University of Cambridge, Cambridge, UK
247. German Center for Neurodegenerative Diseases (DZNE), Rostock, Germany
248. Centro de Investigación Biomédica en Red de Diabetes y Enfermedades Metabólicas Asociadas, CIBERDEM, Spain, Hospital Clínico San Carlos, Madrid, Spain
249. Glenn Biggs Institute for Alzheimer's and Neurodegenerative Diseases, San Antonio, TX, USA

##### The GR@ACE study group

Aguilera N[1], Alarcon E[1], Alegret M[1,2], Boada M[1,2], Buendia M[1], Cano A[1], Cañabate P[1,2], Carracedo A[4,5], Corbat´on-Anchuelo A[6], de Rojas I[1], Diego S[1], Espinosa A[1,2], Gailhajenet A[1], García-González P[1,2], Guitart M[1], Gonz´alez-P´erez A[7], Ibarria M[1], Lafuente A[1], Macias J[8], Maro~nas O[4], Mart´ın E[1], Mart´ınez MT[6], Marqui´e M[1,2], Montrreal L[1], Moreno- Grau S[1,2], Moreno M[1], R. Nuñez-Llaves R[1], Olivé C[1] , Orellana A[1], Ortega G[1,2], Pancho A[1], Pelej`a E[1], P´erez-Cordon A[1], Pineda JA[8], Puerta R[1], Preckler S[1], Quintela I[3], Real LM[3,8], Rosende- Roca M[1], Ruiz A[1,2], S´aez ME[7], Sanabria A[1,2], Serrano-Rios M[6], Sotolongo-Grau O[1], T´arraga L[1,2], Valero S[1,2], Vargas L[1]

1 Research Center and Memory clinic. ACE Alzheimer Center Barcelona, Universitat Internacional de Catalunya, Spain.
2. CIBERNED, Center for Networked Biomedical Research on Neurodegenerative Diseases, National Institute of Health Carlos III, Ministry of Economy and Competitiveness, Spain,
3. Dep. of Surgery, Biochemistry and Molecular Biology, School of Medicine. University of M´alaga. M´alaga, Spain,
4. Grupo de Medicina Xen´omica, Centro Nacional de Genotipado (CEGEN-PRB3-ISCIII). Universidad de Santiago de Compostela, Santiago de Compostela, Spain.
5. Fundaci´on P´ublica Galega de Medicina Xen´omica- CIBERER-IDIS, Santiago de Compostela, Spain.
6. Centro de Investigaci´on Biom´edica en Red de Diabetes y Enfermedades Metab´olicas Asociadas, CIBERDEM, Spain, Hospital Clínico San Carlos, Madrid, Spain,
7. CAEBI. Centro Andaluz de Estudios Bioinform´aticos, Sevilla, Spain
8. Unidad Clínica de Enfermedades Infecciosas y Microbiología. Hospital Universitario de Valme, Sevilla, Spain.

#### DEGESCO consortium

Adarmes-Gómez AD[1,2], Alarcón-Martín E[3], Alonso MD[4], Álvarez I[5], Álvarez V[6,7], Amer-Ferrer G[8], Antequera M[9], Antúnez C[9], Baquero M[10], Bernal M[11], Blesa R[2,12], Boada M[2,3], Buiza-Rueda D[1,2], Bullido MJ[2,14,15], Burguera JA[10], Calero M[2,16,17], Carrillo F[1,2], Carrión-Claro M[1,2], Casajeros MJ[18], Clarimón J[2,12], Cruz-Gamero JM[13], de Pancorbo MM[19], de Rojas I[2,3], del Ser T[15], Diez-Fairen M[5], Escuela R[1,2], Garrote-Espina L[1,2], Fortea J[2,12], Franco E[11], Frank-García A[2,15,20], García-Alberca JM[21], Garcia Madrona S[17], Garcia-Ribas G[17], Gómez-Garre P[1,2], Hevilla S[21], Jesús S[1,2], Labrador Espinosa MA[1,2], Lage C[2,22], Legaz A[9], Lleó A[2,12], López de Munáin A[23], López-García S[2,22], Macias-García D[1,2], Manzanares S[8,24], Marín M[11], Marín-Muñoz J[9], Marín T[21], Marquié M[2,3], Martín Montes A[2,14,20], Martínez B[9], Martínez C[7,25], Martínez V[9], Martínez-Lage Álvarez P[26], Medina M[2,15], Mendioroz Iriarte M[27], Menéndez- González M[7,28], Mir P[1,2], Montrreal L[3], Orellana A[3], Pastor P[5], Pérez Tur J[2,29,30], Periñán-Tocino T[1,2], Pineda-Sánchez R[1,2], Piñol Ripoll G[2,31], Rábano A[2,16,32], Real de Asúa D[33], Rodrigo S[11], Rodríguez-Rodríguez E[2,22], Royo JL[13], Ruiz A[2,3], Sanchez del Valle Díaz R[34], Sánchez-Juan P[16], Sastre I[2,14], Sotolongo-Grau O[3], Valero S[2,3], Vicente MP[9], Vigo-Ortega R[1,2], Vivancos L[9]

1. Unidad de Trastornos del Movimiento, Servicio de Neurología y Neurofisiología. Instituto de Biomedicina de Sevilla (IBiS), Hospital Universitario Virgen del Rocío/CSIC/Universidad de Sevilla, Seville, Spain,
2. CIBERNED, Network Center for Biomedical Research in Neurodegenerative Diseases, National Institute of Health Carlos III, Spain,
3. Research Center and Memory clinic. ACE Alzheimer Center Barcelona, Universitat Internacional de Catalunya, Spain,
4. Servei de Neurologia. Hospital Clínic Universitari de València, Spain.
5. Fundació per la Recerca Biomèdica i Social Mútua Terrassa, and Memory Disorders Unit, Department of Neurology, Hospital Universitari Mutua de Terrassa, University of Barcelona School of Medicine, Terrassa, Barcelona, Spain,
6. Laboratorio de Genética Hospital Universitario Central de Asturias, Oviedo, Spain
7. Instituto de Investigación Biosanitaria del Principado de Asturias (ISPA), Oviedo, Spain
8. Department of Neurology, Hospital Universitario Son Espases, Palma, Spain,
9. Unidad de Demencias. Hospital Clínico Universitario Virgen de la Arrixaca, Palma, Spain,
10. Servei de Neurologia, Hospital Universitari i Politècnic La Fe, Velencia, Spain
11. Unidad de Demencias, Servicio de Neurología y Neurofisiología. Instituto de Biomedicina de Sevilla (IBiS), Hospital Universitario Virgen del Rocío/CSIC/Universidad de Sevilla, Seville, Spain
12. Memory Unit, Neurology Department and Sant Pau Biomedical Research Institute, Hospital de la Santa Creu i Sant Pau, Universitat Autònoma de Barce- lona, Barcelona, Spain,
13. Dep. of Surgery, Biochemistry and Molecular Biology, School of Medicine. University of Málaga. Málaga, Spain
14. Centro de Biologia Molecular Severo Ochoa (C.S.I.C.-U.A.M.), Universidad Autonoma de Madrid, Madrid, Spain
15. Instituto de Investigacion Sanitaria ‘Hospital la Paz’ (IdIPaz), Madrid, Spain,
16. CIEN Foundation, Queen Sofia Foundation Alzheimer Center, Madrid, Spain
17. Instituto de Salud Carlos III (IS- CIII), Madrid, Spain;
18. [18]Hospital Universitario Ramón y Cajal; Madrid, Spain,
19. BIOMICs, País Vasco; Centro de Investigación Lascaray. Universidad del País Vasco UPV/EHU, Vitoria-Gasteiz, Spain
20. Neurology Service, Hospital Universitario La Paz (UAM), Madrid, Spain,
21. Alzheimer Research Center & Memory Clinic. Andalusian Institute for Neuroscience. Málaga, Spain,
22. Neurology Service, Marqués de Valdecilla University Hospital (University of Cantabria and IDIVAL), Santander, Spain,
23. Hospital Donostia de San Sebastían, San Sebastían, Spain
24. Fundación para la Formación e Investigación Sanitarias de la Región de Murcia, Palma Spain
25. Servicio de Neurología -Hospital de Cabueñes-Gijón, Gijón, Spain
26. Centro de Investigación y Terapias Avanzadas. Fundación CITA-alzheimer, San Sebastian, Spain
27. Navarrabiomed, Pamplona, Spain
28. Servicio de Neurología Hospital Universitario Central de Asturias, Oviedo, Spain
29. Unitat de Genètica Molecular. Institut de Biomedicina de València-CSIC, Vencia, Spain
30. Unidad Mixta de Neurologia Genètica. Instituto de Investigación Sanitaria La Fe, Valencia, Spain
31. Unitat Trastorns Cognitius, Hospital Universitari Santa Maria de Lleida, Institut de Recerca Biomédica de Lleida (IRBLLeida), Lleida, Spain
32. BT-CIEN,
33. Hospital Universitario La Princesa, Madrid, Spain,
34. Hospital Clínic Barcelona, Spain

#### Demgene

Alexey A Shadrin[1,2], Shahram Bahrami[1,2], Arvid Rongve[3,4], Geir Bråthen[5,6], Ingunn Bosnes[7,8], Eystein Stordal[7,8], Lavinia Athanasiu[1,2], Per Selnes[9], Ingvild Saltvedt[5,10], Sigrid B. Sando[5,6], Sverre Bergh[11], Ingun Ulstein[12], Srdjan Djurovic[13,14], Tormod Fladby[9,15], Dag Aarsland[16,17], Geir Selbæk[12,15,18], Ole A. Andreassen[1,2]

#### EADI

Céline Bellenguez[1], Benjamin Grenier-Boley[1], Jacques Epelbaum[2], David Wallon[3], Didier Hannequin[3], Florence Pasquier[4], Claudine Berr[5], Jean-Francois Dartigues[6], Dominique campion[7], Christophe Tzourio[8], Vincent Dermecourt[4], Nathalie Fievet[1], Olivier Hanon[9], Carole Dufouil[8], Alexis Brice[10], Bruno Dubois[11], Karen Ritchie[5], Phillippe Amouyel[1], Jean-Charles Lambert[1]

1. Univ. Lille, Inserm, CHU Lille, Institut Pasteur Lille, U1167-RID-AGE - Facteurs de risque et déterminants moléculaires des maladies liées au vieillissement, F-59000 Lille, France
2. UMR 894, Center for Psychiatry and Neuroscience, INSERM, Université Paris Descartes, F-75000 Paris , France
3. Normandie Univ, UNIROUEN, Inserm U1245, CHU Rouen, Department of Neurology and CNR-MAJ, F 76000, Normandy Center for Genomic and Personalized Medicine, Rouen, France
4. Univ. Lille, Inserm, CHU Lille, UMR1172, Resources and Research Memory Center (MRRC) of Distalz, Licend, Lille France
5. Univ. Montpellier, Inserm U1061, Neuropsychiatry: epidemiological and clinical research, PSNREC, Montpellier, France
6. University Bordeaux, Inserm, Bordeaux Population Health Research Center, France
7. Normandie Univ, UNIROUEN, Inserm U1245 and CHU Rouen, Department of Genetics and CNR-MAJ, Rouen, France
8. University Bordeaux, Inserm, Bordeaux Population Health Research Center, France
9. Université de Paris, EA 4468, APHP, Hôpital Broca, Paris, France
10. Inserm U1127, CNRS UMR7225, Sorbonne Universités, UPMC Univ Paris 06, UMR_S1127, Institut du Cerveau et de la Moelle épinière, F-75013, Paris, France; 22. APHP, Department of genetics, Pitié-Salpêtrière Hospital, 75013, Paris, France
11. Institut de la Mémoire et de la Maladie d'Alzheimer (IM2A), Département de Neurologie, Hôpital de la Pitié-Salpêtrière, AP-HP, Paris, France; Institut des Neurosciences Translationnelles de Paris (IHU-A-ICM), Institut du Cerveau et de la Moelle Epinière (ICM), Paris, France; 26. INSERM, CNRS, UMR-S975, Institut du Cerveau et de la Moelle Epinière (ICM), Paris, France; Sorbonne Universités, Université Pierre et Marie Curie, Hôpital de la Pitié-Salpêtrière, AP-HP, Paris, France

#### GERAD

Denise Harold[1], Paul Hollingworth[2], Rebecca Sims[2], Amy Gerrish[2], Nicola Denning[2], Amy Williams[2], Charlene Thomas[2], Alun Meggy[2,3], Rachel Marshall[2], Chloe Davies[2], Lauren Luckcuck[2,3], William Nash[2], Kimberley Dowzell[2], Atahualpa Castillo Morales[2,3], Mateus Bernardo-Harrington[2,3], Patrick Kehoe[4], Per Hoffmann[4], Seth Love[4], James Turton[5], Jenny Lord[5], Kristelle Brown[5], Kevin Morgan[5], Emma Vardy[6], Elizabeth Fisher[7], Jason D. Warren[7], Jonathan M. Schott[7], Martin Rossor[7], Natalie S. Ryan[7], Nick C. Fox[7], Rita Guerreiro[7], Simon Mead[7], James Uphill[8], John Collinge[8], Michelle Lupton[8], Ammar Al-Chalabi[9], Christopher E. Shaw[9], Nick Bass[10], Richard Abraham[11], Reinhard Heun[11], Heike Kölsch[11], Britta Schürmann[11], Frank Jessen[11,17], Wolfgang Maier[11,17],André Lacour[12], Christine Herold[12], Simon Lovestone[13], Bernadette McGuinness[14],

David Craig[14], Janet A. Johnston[14], Michael Gill[14], Peter Passmore[14], Stephen Todd[14], John Powell[15], Petra Proitsi[15], Yogen Patel[15], Angela Hodges[16], Tim Becker[17,19], A. David Smith[20], Donald Warden[20], Gordon Wilcock[20], Robert Clarke[21], Aoibhinn Lynch[22], Brian Lawlor[22], Michael Gill[22, 23], Andrew McQuillin[24], Gill Livingston[24], John Hardy[25], David C. Rubinsztein[26], Carol Brayne[27], Rhian Gwilliam[28], Panagiotis Deloukas[28], Yoav Ben-Shlomo[29], David Mann[30], Nigel M. Hooper[31], Stuart Pickering-Brown[31], Clive Holmes[32], Rebecca Sussams[32], Nick Warner[33], Anthony Bayer[34], Andrew B. Singleton[35], Annette M Hartmann[36], Dan Rujescu[36], Ina Giegling[36], Harald Hampel[37, 38], Martin Dichgans[39], Isabella Heuser[40], Dmitriy Drichel[41], Norman Klopp[42], Markus M. Nöthen[43, 44], Manuel Mayhaus[45], Matthias Riemenschneider[45], Sabrina Pinchler[45], Thomas Feulner[45], Wei Gu[45], Hendrik van den Bussche[46], Martin Scherer[46], Jens Wiltfang[47], Johannes Kornhuber[48], Michael Hüll[49], Lutz Frölich[50], H-Erich Wichmann[51], Karl-Heinz Jöckel[52], Susanne Moebus[52], Steffi Riedel-Heller[53], John Kauwe[54], John Morris[55,58], Kevin Mayo[55,56,57], Magda Tsolaki[59], Michael O'Donovan[2], Lesley Jones[2], Michael Owen[2], Valentina Escott-Price[2], Alfredo Ramirez[18, 19], Peter Holmans[2], Julie Williams[2,3]

1. School of Biotechnology, Dublin City University, Dublin, Ireland.
2. Division of Psychological Medicine and Clinical Neurosciences, Medical Research Council (MRC) Centre for Neuropsychiatric Genetics & Genomics, Cardiff University, Cardiff, UK.
3. UK Dementia Research Institute at Cardiff, Cardiff University, Cardiff, UK.
4. University of Bristol Medical School, Learning & Research level 2, Southmead Hospital, Bristol, UK.
5. Institute of Genetics, Queen's Medical Centre, University of Nottingham, UK
6. Institute for Ageing and Health, Newcastle University, Biomedical Research Building, Campus for Ageing and Vitality, Newcastle upon Tyne, UK
7. Department of Neurodegenerative Disease, UCL Institute of Neurology, London, UK.
8. Department of Neurodegenerative Disease, MRC Prion Unit at UCL, Institute of Prion Diseases, London, UK
9. MRC Centre for Neurodegeneration Research, Department of Clinical Neuroscience, King's College London, Institute of Psychiatry, London, UK.
10. Division of Psychiatry, University College London, London, UK.
11. Department of Psychiatry and Psychotherapy, University of Bonn, Bonn, Germany
12. Deutsches Zentrum für Neurodegenerative Erkrankungen (DZNE, Bonn), Bonn, Germany
13. Department of Psychiatry, University of Oxford, Oxford, UK.
14. Ageing Group, Centre for Public Health, School of Medicine, Dentistry and Biomedical Sciences, Queen's University, Belfast, UK.
15. Department of Basic and Clinical Neuroscience, Institute of Psychiatry, Psychology and Neuroscience, King's College London, London, UK.
16. Department of Old Age Psychiatry, Institute of Psychiatry, Psychology and Neuroscience, King's College London, London, UK.
17. German Centre for Neurodegenerative Diseases, Bonn, Germany.
18. Department for Neurodegenerative Diseases and Geriatric Psychiatry, University Hospital Bonn, Bonn, Germany.
19. Institute for Medical Biometry, Informatics and Epidemiology, University of Bonn, Bonn, Germany
20. Oxford Project to Investigate Memory and Ageing (OPTIMA), University of Oxford, Nuffield Department of Clinical Neurosciences, John Radcliffe Hospital, Oxford, UK
21. Oxford Healthy Aging Project, Clinical Trial Service Unit, University of Oxford, Oxford, UK.
22. Mercer's Institute for Research on Aging, St. James's Hospital and Trinity College, Dublin, Ireland.
23. St. James's Hospital and Trinity College, Dublin, Ireland.
24. Department of Mental Health Sciences, University College London, UK.
25. Department of Molecular Neuroscience, UCL, Institute of Neurology, London, UK.
26. Cambridge Institute for Medical Research, University of Cambridge, Cambridge, UK
27. Institute of Public Health, University of Cambridge, Cambridge, UK.
28. The Wellcome Trust Sanger Institute, Hinxton, Cambridge, UK.

29. Population Health Sciences, Bristol Medical School, University of Bristol, Bristol, UK.
30. Clinical Neuroscience Research Group, Greater Manchester Neurosciences Centre, University of Manchester, Salford, UK
31. Division of Neuroscience and Experimental Psychology, School of Biological Sciences, Faculty of Biology, Medicine and Health, University of Manchester, Manchester Academic Health Science Centre, Manchester, UK.
32. Division of Clinical Neurosciences, School of Medicine, University of Southampton, Southampton, UK.
33. Somerset Partnership NHS Trust, Somerset, UK.
34. Institute of Primary Care and Public Health, Cardiff University, University Hospital of Wales, Cardiff, UK.
35. Laboratory of Neurogenetics, National Institute on Aging, National Institutes of Health, Bethesda, MD, 20892, USA.
36. Department of Psychiatry, Martin Luther University Halle-Wittenberg, Halle, Germany.
37. Department of Psychiatry, University of Frankfurt, Frankfurt am Main, Germany.
38. Department of Psychiatry, Ludwig Maximilians University, Munich, Germany.
39. Institute for Stroke and Dementia Research, Klinikum der Universität München, Munich, Germany.
40. Department of Psychiatry and Psychotherapy, Charité University Medicine, Berlin, Germany.
41. Cologne Center for Genomics, University of Cologne, Cologne, Germany.
42. Institute of Epidemiology, Helmholtz Zentrum München, German Research Center for Environmental Health, Neuherberg, Munich, Germany.
43. Institute of Human Genetics, University of Bonn, Bonn, Germany.
44. Department of Genomics, Life & Brain Center, University of Bonn, Bonn, Germany.
45. Department of Psychiatry and Psychotherapy, University Hospital, Saarland, Germany.
46. Institute of Primary Medical Care, University Medical Center Hamburg-Eppendorf, Germany
47. Department of Psychiatry and Psychotherapy, University Medical Center Goettingen, Goettingen, Germany
48. Department of Psychiatry and Psychotherapy, University of Erlangen-Nuremberg, Erlangen, Germany
49. Department of Psychiatry, University of Freiburg, Freiburg, Germany.
50. Central Institute of Mental Health, Medical Faculty Mannheim, University of Heidelberg, Heidelberg, Germany.
51. Institute of Epidemiology, Helmholtz Zentrum München, German Research Center for Environmental Health, Neuherberg, Germany
52. Institute for Medical Informatics, Biometry and Epidemiology, University Hospital of Essen, University Duisburg-Essen, Essen, Germany.
53. Institute of Social Medicine, Occupational Health and Public Health, University of Leipzig, Leipzig, Germany.
54. Departments of Biology, Brigham Young University, Provo, UT, USA.
55. Department of Psychiatry, Washington University School of Medicine, St. Louis, MO, USA.
56. Department of Neurology, Washington University, St. Louis, MO, USA.
57. Department of Genetics, Washington University, St. Louis, MO, USA
58. Hope Center Program on Protein Aggregation and Neurodegeneration, Washington University School of Medicine, St. Louis, MO, USA.
59. Department of Neurology, Medical School, Aristotle University of Thessaloniki, Thessaloniki, Greece.

#### Asian Parkinson's Disease Genetics Consortium (APDGC)

Jia Nee Foo[1,2], Elaine Guo Yan Chew[1,2], Sun Ju Chung[3], Rong Peng[4], Yinxia Chao[5,6], Louis CS Tan[6,7], Moses Tandiono[1,2], Michelle M Lian[1,2], Ebonne Y Ng[5], Kumar-M. Prakash[5], Wing-Lok Au[7], Wee-Yang Meah[2], Shi Qi Mok[2], Azlina Ahmad Annuar[8], Anne YY Chan[9], Ling Chen[10], Yongping

Chen[4], Beom S Jeon[11], Lulu Jiang[10], Jia Lun Lim[8,12], Juei-Jueng Lin[13], Chunfeng Liu[14], Chengjie Mao[14], Vincent Mok[9], Zhong Pei[10], Hui-Fang Shang[4], Chang-He Shi[15], Kyuyoung Song[16], Ai Huey Tan[12], Yih-Ru Wu[17], Yu-ming Xu[15], Renshi Xu[18], Yaping Yan[19], Jing Yang[15], Bao Rong Zhang[19], Woon-Puay Koh[10], Shen-Yang Lim[12], Chiea Chuen Khor[2,20], Jianjun Liu[2], Eng-King Tan[5,6]

1. Lee Kong Chian School of Medicine, Nanyang Technological University Singapore, 11 Mandalay Road, Singapore 308232, Singapore
2. Human Genetics, Genome Institute of Singapore, A*STAR, 60 Biopolis Street, Singapore 138672, Singapore
3. Department of Neurology, Asan Medical Center, University of Ulsan College of Medicine, Seoul, South Korea
4. Department of Neurology, West China Hospital, Sichuan University, Chengdu, Sichuan Province 610041, Sichuan, People's Republic of China
5. Department of Neurology, National Neuroscience Institute, Singapore General Hospital, 20 College Road, Singapore 169856, Singapore
6. Duke-National University of Singapore Medical School, 8 College Road, Singapore 169857, Singapore
7. Department of Neurology, National Neuroscience Institute, 11 Jalan Tan Tock Seng, Singapore 308433, Singapore
8. Department of Biomedical Science, Faculty of Medicine, University of Malaya, Kuala Lumpur, Malaysia
9. Margaret K.L. Cheung Research Centre for Management of Parkinsonism, Gerald Choa Neuroscience Centre, Lui Che Woo Institute of Innovative Medicine, Division of Neurology, Department of Medicine and Therapeutics, Prince of Wales Hospital, The Chinese University of Hong Kong, Hong Kong, Hong Kong SAR, People's Republic of China
10. Department of Neurology, The First Affiliated Hospital,Sun Yat-Sen University, 58 Zhongshan Road II, Guangzhou 510080, People's Republic of China
11. Department of Neurology, Seoul National University Hospital, 101 Daehak-ro, Jongno-gu, Seoul 110-744, South Korea
12. Department of Medicine and the Mah Pooi Soo and Tan Chin Nam Centre for Parkinson's and Related Disorders, Faculty of Medicine, University of Malaya, Kuala Lumpur, Malaysia
13. Department of Neurology, Chushang Show-Chwan Hospital, No.75 Jishan Road Section 2, Zhushan District, Nantou, Taiwan
14. Department of Neurology, Second Affiliated Hospital of Soochow University, 1055 Sanxiang Road, Suzhou 215004, People's Republic of China
15. Department of Neurology, The First Affiliated Hospital of Zhengzhou University, Zhengzhou 450000, Henan, People's Republic of China
16. Department of Biochemistry and Molecular Biology, University of Ulsan College of Medicine, Seoul, South Korea
17. Department of Neurology, Chang Gung Memorial Hospital, Chang Gung University, Taipei 10507, Taiwan
18. Department of Neurology, Jiangxi Provincial People's Hospital, Nanchang 330006, Jiangxi, People's Republic of China
19. Department of Neurology, Second Affiliated Hospital, College of Medicine, Zhejiang University, Hangzhou 310009, Zhejiang Province, People's Republic of China
20. Singapore Eye Research Institute, 20 College Road Discovery Tower, Level 6 The Academia, Singapore 169856, Singapore

### Supplementary Methods

#### QUALITY CONTROL AND ANALYSIS PER DATASET

##### ***Alzheimer’s Disease – ADSP & ADGC datasets***

###### 1) Participants and sources of data

Phenotypic information and genotypes were obtained from publicly released genome-wide association study datasets assembled by the Alzheimer's Disease Genetics Consortium (ADGC) and derived from whole-genome sequencing (WGS) data generated by the Alzheimer Disease Sequencing Project (ADSP), with phenotype and genotype ascertainment described elsewhere. The cohorts' queried accession numbers, as well as the sequencing technology or single nucleotide polymorphism (SNP) genotyping platforms are described in **Supplementary Tables 8 and 9**. The microarray datasets are largely part of the ADGC and as such they will be referred thereafter as the ADGC.

###### 2) Quality control procedures

Prior to ancestry, principal components and relatedness determination, and HLA-alleles imputation, in each cohort-platform, variants were excluded based on genotyping rate (< 95%), MAF < 1%, and Hardy-Weinberg equilibrium in controls ($p < 10^{-6}$) using PLINK v1.9[1]. gnomAD[2] database-derived information was used to filter out SNPs that met one of the following exclusion criteria[3,4]: (i) located in a low complexity region, (ii) located within common structural variants (MAF > 1%), (iii) multiallelic SNPs with MAF > 1% for at least two alternate alleles, (iv) located within a common insertion/deletion, (v) having any flag different than PASS in gnomADv.3, (vi) having potential probe polymorphisms. The latter are defined as SNPs for which the probe may have variable affinity due to the presence of other SNP(s) within 20 bp and with MAF > 1%. Individuals with more than 5% genotype missingness were excluded. Duplicate individuals were identified with KING[5] and their clinical, diagnostic and pathological data (including age-at-onset of cognitive symptoms, age-at-examination for clinical diagnosis, age-at-last exam, age-at-death), as well as sex, race, and *APOE* genotype were cross-referenced across cohorts. Duplicate entries with irreconcilable phenotype or discordant sex were flagged for exclusion.

###### 3) Ancestry determination

For each cohort, we first determined the ancestry of each individual with SNPWeights v2[6] using reference populations from the 1000 Genomes Consortium[7]. By applying an ancestry percentage cut-off > 75%, the samples were stratified into five super populations: South-Asians, East-Asians, Amerindians, Africans, and Europeans, and an Admixed group composed of individuals not passing the 75% cut-off in any single ancestry (**Supplementary Table 9**)[3]. The analyses were split into three ancestry groups: Europeans, Africans, and Amerindians-Latinos. The first two groups are composed of individuals passing the 75% threshold in their respective ancestry. The Amerindian-Latinos includes individuals in the Amerindians ancestry group (75% cut-off), and individuals in the Admixed group with at least 15% Amerindians and who identified as Hispanic/Latinos ethnicity. The rationale to include these additional individuals is to compensate the paucity of the Amerindians only group and to have a similar ancestry composition as in the Latin American Research Consortium on the Genetics of Parkinson's Disease (LARGE-PD). Last, enriching for Amerindians ancestry enables us to assess the effect of *HLA-DRB1**04:07 since the HLA haplotype DQA1*03:01~ DQB1*03:02~ DRB1*04:07 is a common haplotype in this ancestry group.

4) **Imputation**

Each cohort-genotyping platform was imputed on the TOPMed imputation server per ancestry group to obtain an imputation quality ($R^2$) per ancestry group. For the local-GWAS at the HLA locus we retained variants with $R^2 > 0.30$, MAF > 1%, and present in 50% of the imputed cohorts.
The HLA -alleles and -amino-acids were imputed on platform and ancestry specific reference panels available through HIBAG[8] or trained in-house as previously described[9]. In the allele-level analyses, alleles with an imputation posterior probability lower than 0.5 were considered as undetermined as recommended by HIBAG developers, and only allele with carrier frequency above1% were retained for analysis. In the haplotype-level analyses, only individuals with non-missing allele genotypes were included in the haplotype level analysis. Three-locus HLA class I or class II haplotypes were determined using the haplo.em function from the R haplo.stats package. Only haplotypes with posterior probability >0.5 and a carrier frequency of >1% were included in the analysis. In the amino-acid-level analyses, HIBAG[8] was used to convert P-coded alleles to amino acid sequences for exon 2, 3 of HLA class I genes, and exon 2 of class II genes.

5) **Samples retained for analysis**

**Supplementary Table 10** describes the demographics of individuals retained for analysis. Analyses were implemented into 6 different groups separating WGS data and TOPMed imputed and by ancestry group: ADSP-European, ADSP-African, ADSP-Amerindian-Latino, ADGC-European, ADGC-African, ADGC-Amerindian-Latino.

##### 6) Statistical analyses

In the following paragraph a variable refers indifferently to a variant in the local-GWAS at HLA locus, a HLA-allele, a HLA-haplotype, or a HLA-amino-acids. The AD risk associated with each variable was estimated using a linear mixed model regression on case-control diagnosis. The HLA -allele, -haplotype, and -amino-acids level analyses were run as dominant model (collapsing homozygotes for the minor frequency variable with heterozygotes). All statistical analyses were performed in R (v4.0.2) and adjusted for sex, six genetic principal components estimated with the *PC-Air* method[10] implemented in *GENESIS*[11], and covaried by a sparse genetic relationship matrix estimated with the *PC-Relate* method[12] implemented in *GENESIS*. Case-control analyses were not adjusted for age given that controls were older than cases in some subgroups. Correcting for age when cases are younger than controls leads to the model incorrectly inferring the age effect on AD risk, resulting in statistical power loss[3].

#### ***<u>Alzheimer's Disease – UK Biobank dataset</u>***

##### 1) Participants, quality control and variant imputation

The UK Biobank data includes 488,377 participants which were genotyped on single nucleotide polymorphism (SNP) microarrays and imputed at high resolution using two reference panels: (i) the Haplotype Reference Consortium (HRC) for most variants with minor allele frequency > 0.001 and (ii) the UK10K+1000Genomes for variants not in the HRC panel[13]. The quality control prior to imputation has been extensively described in Bycroft et al.[13]. The proxy-AD phenotype defined in Bellenguez et al.[14] (i.e., cases are individuals who have an ICD10 code linked to AD in their medical record[15] or reported a first degree with Alzheimer's disease, March, 2021 release). We restricted our analysis to 388,051 unrelated individuals after pruning for 3$^{rd}$ degree relatedness using the following criteria to rank order individuals for removal: (i) highest number of relatives, (ii) not a proxy-AD case (iii) and youngest individual.

##### 2) Ancestry determination

We split the UK Biobank unrelated individuals into two groups: British ancestry and non-British/other ancestries. The British group correspond to identified who self-identified as white British and who clustered on together in the principal ancestry component analysis performed in Bycroft et al. (field ID: 22006). The British ancestry group was composed of 52,426 proxy-AD cases, and 272,624 controls. The non-British/other ancestries group was composed of 7,840 proxy-AD cases and 55,161 controls. This last group was heterogeneous in term of ancestral origin, but most individuals identified as non-British European.

##### 3) HLA Imputation

The HLA -alleles and -amino-acids were imputed on platform and ancestry specific reference panels available through HIBAG[8] or trained in-house as previously described[9]. In the allele-level analyses, alleles with an imputation posterior probability lower than 0.5 were considered as undetermined as recommended by HIBAG developers, and only allele with carrier frequency above1% were retained for analysis. In the haplotype-level analyses, only individuals with non-missing allele genotypes were included in the haplotype level analysis. Three-locus HLA class I or class II haplotypes were determined using the haplo.em function from the R haplo.stats package. Only haplotypes with posterior probability >0.5 and a carrier frequency of >1% were included in the analysis. In the amino-acid-level analyses, HIBAG[8] was used to convert P-coded alleles to amino acid sequences for exon 2, 3 of HLA class I genes, and exon 2 of class II genes.

##### 4) Statistical analyses

In the following paragraph a variable refers indifferently to a variant in the local-GWAS at HLA locus, a HLA-allele, a HLA-haplotype, or a HLA-amino-acids. The HLA -allele, -haplotype, and -amino-acids level analyses were run as dominant model (collapsing homozygotes for the minor frequency variable with heterozygotes). Proxy-AD association were tested with plink2 (v2.00a2LM) using the –glm flag covarying for age at last visit, sex, genotyping array, assessment center and the first 20 PCs provided by the UK Biobank.

***Alzheimer's Disease – EADB, GR@ACE, GERAD, EADI, DemGene, Bonn, CCHS datasets***

Demographics, quality control and GWAS analysis are fully described in [14] and demographics are also shown in **Supplementary Table 11**. The HLA analyses was conducted plink2 (v2.00a2LM) using the –glm flag covariates per cohort were described in [14].

***Alzheimer's Disease – NCGG dataset***

Demographics, quality control and GWAS analysis are fully described in [16]. The HLA analyses was conducted plink2 (v2.00a2LM) using the –glm flag covariates per cohort were described in [16].

***Alzheimer's Disease – GARD dataset***

Demographics, quality control and GWAS analysis are fully described in [17]. The HLA analyses was conducted plink2 (v2.00a2LM) using the –glm flag covariates per cohort were described in [17].

**Alzheimer's Disease –JGSCAD dataset**

Demographics, quality control and GWAS analysis are fully described in [18]. The HLA analyses was conducted plink2 (v2.00a2LM) using the –glm flag covariates per cohort were described in [18].

***Alzheimer's Disease Neuropathology – NACC and RUSH datasets***

1) **Participants and sources of data**

Participants were enrolled and followed up at one of Alzheimer's Disease Center (ADC) across the US. Genetic data were obtained from the Rush Religious Orders Study and Memory and Aging Project (ROSMAP) [19] and from the Alzheimer's Disease Center (ADC) cohorts 1 to 7 parts of the ADGC[20] (see **Supplementary Table 3** for data accession number). ROSMAP samples were assessed by the Rush ADC and their neuropathological assessment followed procedures described respectively in [21]. Neuropathological assessment for samples with genotyping from ADGC was obtained from National Alzheimer's Coordinating Center (NACC) and followed postmortem evaluation protocol [22].

#### 2) Quality control procedures, ancestry determination, and imputation

The content of this section is identical to the corresponding sections in “Alzheimer’s Disease – ADSP & ADGC datasets” given that these samples were included in the association with AD status.

#### 3) Samples retained for analysis

**Supplementary Table 12** describes the demographics of individuals retained for neuropathology analyses: Tau Braak staging, neuritic plaques density. We also defined three categories: AD pathology only, Lewy body (LB) pathology only, and dual pathology (AD and LB) and compared these against controls without AD and LB pathologies. The schematic below describes these categories and follows the classification defined in[23].

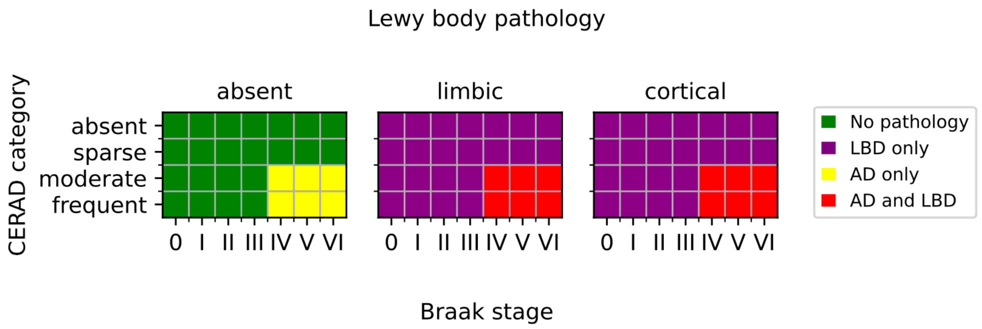

**Supplementary Table 13** provides the demographics and number of individuals per category.

#### 4) Statistical analyses

The statistical analyses follow the method described in the “Alzheimer’s Disease – ADSP & ADGC datasets” corresponding section.

***<u>Alzheimer’s Disease Cerebrospinal Fluid – EADB and Swedish datasets</u>***

#### 1) Participants and sources of data.

The EADB participants (as described above) for which cerebrospinal fluid (CSF) amyloid beta and/or (phosphorylated) Tau measurements were available were included. The Swedish cohorts originate from Gothenburg H70 Birth cohort studies and are clinical AD samples from Sweden all gathered and analyzed in Gothenburg. The genetic data for the

EADB cohorts has been processed in a consistent approach[14], in which the Illumina Infinium Global Screening Array (GSA, GSAsharedCUSTOM_24+v1.0) was predominantly used besides the Axiom 815K Spanish biobank array (Thermo Fisher). The genetic data for the Swedish cohorts were generated with the Illumina Neurochip array.

##### 2) Quality control procedures and imputation

The quality control procedures of the EADB datasets are described here in above[14]. For the Swedish datasets, QC and imputation procedures were previously described elsewhere[24]. In short, low-quality variants were excluded based on call rate, minor alle frequency (MAF< 0.01) and Hardy-Weinberg disequilibrium ($P < 1 \times 10^{-6}$). Individuals were removed based on per-sample call rate, sex mismatch, excessive heterozygosity or non-European ancestry. The Sanger imputation service was used to impute post-QC, using the reference panel of Haplotype Reference Consortium data (HRC1.1). The UCSC LiftOver program (https://genome-store.ucsc.edu/) and Plink v2.0 (www.cog-genomics.org/plink/2.0/) were used to lift the GRCh37 genomic positions to GRCh38, the genomic build for all other datasets.

##### 3) Samples retained for analysis

**Supplementary Table 14** describes the demographics of individuals retained for analysis. The association analyses with HLA haplotypes, alleles and amino acids were only performed for those individuals for which genotype-level data was available (rather than GWAS summary statistics). For the rs601945 association analyses all cohorts were considered.

##### 4) Statistical analyses

For the HLA-locus, -allele, haplotype, and amino acid association analyses, similar association analysis procedures were performed. Within each cohort, linear regression was performed, using PLINK v2.0, for the continuous phenotypes Aβ42, tau and pTau. Association tests were adjusted for gender, age, assay type (if applicable), and ten ancestry principal components. METAL was used for meta-analysis of the per cohort association results, applying the default approach that utilizes p-value and direction of effect, while weighted according to sample size.

Association analyses were repeated for subgroups, stratified according to diagnosis status, resulting in a group only including AD subjects, and one including individuals with no or mild cognitive impairment. Covariates were those described for the main analyses above.

***Parkinson's Disease – IPDGC, McGill, NINDS, NGRC, Oslo, PPMI, APDGC, UK Biobank datasets***

Demographics, phenotyping, quality control, imputation and analysis of the European ancestry cohorts part of local-GWAS at HLA summary statistics have been extensively described in [25]. Similarly, the phenotyping, quality control and HLA imputation PD cohorts used in the HLA -alleles, -haplotypes, and -amino-acids level analysis were previously described in [9], demographics are presented in **Supplementary Table 15**.

***Parkinson's Disease – EastAsians-PD and 23andMe datasets***

For the EastAsians-PD and 23andMe cohorts, the HLA alleles, haplotypes, amino acids statistics were derived from GWAS summary statistics data using the DISH software[26] as described in [27]. Demographics, quality control and GWAS analysis were previously described[27–29] and available demographics are reported in **Supplementary Table 16**.

***Parkinson's Disease – LARGE-PD dataset***

Demographics, quality control and GWAS analysis are fully described in [30] and demographics are also shown in **Supplementary Table 16**.

#### References

1. Chang, C. C. *et al.* Second-generation PLINK: rising to the challenge of larger and richer datasets. *GigaScience* **4**, 7 (2015).
2. Karczewski, K. J. *et al.* The mutational constraint spectrum quantified from variation in 141,456 humans. *Nature* **581**, 434–443 (2020).
3. Le Guen, Y. *et al.* A novel age-informed approach for genetic association analysis in Alzheimer's disease. *Alzheimer's Research & Therapy* **13**, 72 (2021).
4. Le Guen, Y. *et al.* Common X-Chromosome Variants Are Associated with Parkinson Disease Risk. *Annals of Neurology* **90**, 22–34 (2021).
5. Manichaikul, A. *et al.* Robust relationship inference in genome-wide association studies. *Bioinformatics* **26**, 2867–2873 (2010).
6. Chen, C. Y. *et al.* Improved ancestry inference using weights from external reference panels. *Bioinformatics* **29**, 1399–1406 (2013).
7. Auton, A. *et al.* A global reference for human genetic variation. *Nature* **526**, 68–74 (2015).
8. Zheng, X. *et al.* HIBAG—HLA genotype imputation with attribute bagging. *Pharmacogenomics J* **14**, 192–200 (2014).
9. Yu, E. *et al.* Fine mapping of the HLA locus in Parkinson's disease in Europeans. *NPJ Parkinsons Dis* **7**, 84 (2021).
10. Conomos, M. P., Miller, M. B. & Thornton, T. A. Robust Inference of Population Structure for Ancestry Prediction and Correction of Stratification in the Presence of Relatedness. *Genetic Epidemiology* **39**, 276–293 (2015).

11. Gogarten, S. M. *et al.* Genetic association testing using the GENESIS R/Bioconductor package. *Bioinformatics* **35**, 5346–5348 (2019).
12. Conomos, M. P. *et al.* Genetic Diversity and Association Studies in US Hispanic/Latino Populations: Applications in the Hispanic Community Health Study/Study of Latinos. *The American Journal of Human Genetics* **98**, 165–184 (2016).
13. Bycroft, C. *et al.* The UK Biobank resource with deep phenotyping and genomic data. *Nature* **562**, 203–209 (2018).
14. Bellenguez, C. *et al.* New insights on the genetic etiology of Alzheimer's and related dementia. *medRxiv* 2020.10.01.20200659 (2020) doi:10.1101/2020.10.01.20200659.
15. Jansen, I. E. *et al.* Genome-wide meta-analysis identifies new loci and functional pathways influencing Alzheimer's disease risk. *Nature Genetics* **51**, 404–413 (2019).
16. Shigemizu, D. *et al.* Ethnic and trans-ethnic genome-wide association studies identify new loci influencing Japanese Alzheimer's disease risk. *Transl Psychiatry* **11**, 151 (2021).
17. Kang, S. *et al.* Potential Novel Genes for Late-Onset Alzheimer's Disease in East-Asian Descent Identified by APOE-Stratified Genome-Wide Association Study. *J Alzheimers Dis* **82**, 1451–1460 (2021).
18. Miyashita, A. *et al.* SORL1 Is Genetically Associated with Late-Onset Alzheimer's Disease in Japanese, Koreans and Caucasians. *PLOS ONE* **8**, e58618 (2013).
19. Bennett, D. A. *et al.* Overview and findings from the rush Memory and Aging Project. *Current Alzheimer research* **9**, 646–63 (2012).

20. Kunkle, B. W. *et al.* Genetic meta-analysis of diagnosed Alzheimer's disease identifies new risk loci and implicates Aβ, tau, immunity and lipid processing. *Nature Genetics* **51**, 414–430 (2019).

21. Schneider, J. A. *et al.* Cognitive impairment, decline and fluctuations in older community-dwelling subjects with Lewy bodies. *Brain* **135**, 3005–3014 (2012).

22. Besser, L. M. *et al.* The revised national Alzheimer's coordinating center's neuropathology form-available data and new analyses. *Journal of Neuropathology and Experimental Neurology* **77**, 717–726 (2018).

23. Tsuang, D. *et al.* APOE ϵ4 increases risk for dementia in pure synucleinopathies. *JAMA Neurology* **70**, 223–228 (2013).

24. Najar, J. *et al.* Polygenic risk scores for Alzheimer's disease are related to dementia risk in APOE ε4 negatives. *Alzheimer's & Dementia: Diagnosis, Assessment & Disease Monitoring* **13**, e12142 (2021).

25. Nalls, M. A. *et al.* Identification of novel risk loci, causal insights, and heritable risk for Parkinson's disease: a meta-analysis of genome-wide association studies. *The Lancet Neurology* **18**, 1091–1102 (2019).

26. Lim, J., Bae, S.-C. & Kim, K. Understanding HLA associations from SNP summary association statistics. *Sci Rep* **9**, 1337 (2019).

27. Naito, T. *et al.* Trans-Ethnic Fine-Mapping of the Major Histocompatibility Complex Region Linked to Parkinson's Disease. *Mov Disord* **36**, 1805–1814 (2021).

28. Foo, J. N. *et al.* Identification of Risk Loci for Parkinson Disease in Asians and Comparison of Risk Between Asians and Europeans: A Genome-Wide Association Study. *JAMA Neurol* **77**, 746–754 (2020).

29. Satake, W. *et al.* Genome-wide association study identifies common variants at four loci as genetic risk factors for Parkinson's disease. *Nat Genet* **41**, 1303–1307 (2009).

30. Loesch, D. P. *et al.* Characterizing the Genetic Architecture of Parkinson's Disease in Latinos. *Ann Neurol* **90**, 353–365 (2021).

31. Reynisson, B. *et al.* Improved Prediction of MHC II Antigen Presentation through Integration and Motif Deconvolution of Mass Spectrometry MHC Eluted Ligand Data. *J. Proteome Res.* **19**, 2304–2315 (2020).

32. Racle, J. *et al.* Robust prediction of HLA class II epitopes by deep motif deconvolution of immunopeptidomes. *Nat Biotechnol* **37**, 1283–1286 (2019).

**Supplementary Table 1. Number of individuals per disease and ancestry group included in the meta-analyses.**

| | Ancestry | N cases/ proxy-cases | N controls | Reference doi |
|---|---|---|---|---|
| **Parkinson's disease** | | | | |
| European datasets in Yu et al. 2021 | European | 33984 | 490861 | doi:10.1038/s41531-021-00231-5 |
| 23andMe individuals (i) in Nalls et al. 2014 | European | 3261 | 29499 | doi:10.1038/ng.3043 |
| 23andMe individuals (ii) in Nalls et al. 2014 | European | 866 | 32538 | doi:10.1038/ng.3043 |
| 23andMe individuals in Chang et al. 2017 | European | 6476 | 302042 | doi:10.1038/ng.3955 |
| 23andMe individuals in Nalls et al. 2019 | European | 2448 | 571441 | doi:10.1016/s1474-4422(19)30320-5 |
| Japaneses in Naito et al. 2021 | East Asian | 988 | 2521 | doi:10.1002/mds.28583 |
| Taiwaneses in Foo et al. 2020 | East Asian | 216 | 225 | doi:10.1001/jamaneurol.2020.0428 |
| Singaporeans/Malays in Foo et al. 2020 | East Asian | 2536 | 21840 | doi:10.1001/jamaneurol.2020.0428 |
| South Koreans in Foo et al. 2020 | East Asian | 1494 | 599 | doi:10.1001/jamaneurol.2020.0428 |
| Hong-Kongers in Foo et al. 2020 | East Asian | 199 | 166 | doi:10.1001/jamaneurol.2020.0428 |
| Chineses in Foo et al. 2020 | East Asian | 2279 | 2021 | doi:10.1001/jamaneurol.2020.0428 |
| Amerindian/European-Latinos in Loesch et al. 2021 | Amerindian-European | 807 | 690 | doi:10.1002/ana.26153 |
| | **Total** | **55554** | **1454443** | |
| **Alzheimer's disease** | | | | |
| ADGC-European TOPMed imputed | European | 13027 | 12748 | doi:10.1038/s41588-019-0358-2 |
| ADSP-European WGS | European | 4127 | 3020 | doi:10.1038/s41588-019-0358-2 |
| UK Biobank – British ancestry | European | 52426 | 272624 | doi:10.1038/s41586-018-0579-z |
| UK Biobank – non-British/Other ancestries | European | 7840 | 55161 | doi:10.1038/s41586-018-0579-z |
| EADB, GR@ACE, GERAD, EADI, DemGene, Bonn, CCHS | European | 35084 | 55762 | doi:10.1101/2020.10.01.20200659 |
| ADGC-African TOPMed imputed | African | 253 | 1837 | doi:10.1001/jamaneurol.2020.3536 |
| ADSP-African WGS | African | 849 | 1240 | doi:10.1001/jamaneurol.2020.3536 |
| South Koreans – GARD | East Asian | 872 | 895 | doi:10.1101/2020.07.02.20145557 |
| Japanese – JGSCAD | East Asian | 374 | 376 | doi:10.1371/journal.pone.0058618 |
| Japanese – NCGG | East Asian | 3744 | 3093 | doi:10.1038/s41398-021-01272-3 |
| ADGC-Amerindian-Latino TOPMed imputed | Amerindian-European | 1542 | 1906 | unpublished |
| ADSP-Amerindian-Latino WGS | Amerindian-European | 1233 | 2327 | unpublished |
| | **Total** | **121371** | **410989** | |

**Supplementary Table 2. HLA -alleles, -haplotypes, -amino-acids levels association across tested variables in Alzheimer's Disease.**

External spreadsheet.

**Supplementary Table 3. HLA -alleles, -haplotypes, -amino-acids levels association across tested variables in Parkinson's Disease.**

External spreadsheet.

**Supplementary Table 4.** **(on the next page)**

**Supplementary Table 5. List of Tau peptides that were tested for binding with *HLA-DRB1**04:01, *HLA-DRB1**04:04, and *HLA-DRB1**04:05.**

External spreadsheet.

**Supplementary Table 6. List of α-synuclein peptides that were tested for binding with *HLA-DRB1**04:01, *HLA-DRB1**04:04, and *HLA-DRB1**04:05.**

External spreadsheet.

**Supplementary Table 4. *HLA-DRB1**04 alleles are associated with reduced Tau and neurofibrillary tangles but not with Amyloid-β or neuritic plaques, when testing their association with Alzheimer's disease neuropathology and cerebrospinal fluid biomarkers.** p-Tau: phosphorylated Tau, t-Tau: total Tau, N: number of individuals, MAF: minor allele frequency, OR: odds ratio, β: parameter estimate, CI: confidence interval. Braak: Tau Braak staging, Neur: Neuritic plaques density.

| | | DRB1*04:01 | | | | DRB1*04:04 | | | | DRB1 H13 | | | |
|---|---|---|---|---|---|---|---|---|---|---|---|---|---|
| | **Phenotype** | **N** | **Freq** | **β [95% CI]** | **pval** | **N** | **Freq** | **β [95% CI]** | **pval** | **N** | **Freq** | **β [95% CI]** | **pval** |
| All individuals | **Tau Braak staging** | 6804 | 0.164 | -0.17[-0.28; -0.07] | 1.1E-03 | 6804 | 0.058 | 0.06[-0.11; 0.22] | 0.5 | 7456 | 0.293 | -0.13[-0.21; -0.05] | 1.4E-03 |
| | **Neuritic plaques density** | 5385 | 0.165 | -0.06[-0.14; 0.01] | 0.11 | 5385 | 0.057 | -0.02[-0.14; 0.1] | 0.72 | 5876 | 0.292 | -0.04[-0.1; 0.02] | 0.19 |
| | **total-Tau in CSF** | 5392 | 0.16 | -0.05[-0.12; 0.02] | 0.17 | 5392 | 0.052 | -0.24[-0.36; -0.12] | 1.1E-04 | 5289 | 0.232 | -0.11[-0.17; -0.05] | 5.5E-04 |
| | **p-Tau in CSF** | 5371 | 0.16 | -0.02[-0.09; 0.06] | 0.66 | 5371 | 0.052 | -0.29[-0.41; -0.17] | 2.1E-06 | 5269 | 0.234 | -0.08[-0.14; -0.02] | 1.0E-02 |
| | **Aβ42 in CSF** | 5471 | 0.16 | 0.07[-0.01; 0.14] | 0.07 | 5471 | 0.051 | 0.09[-0.03; 0.21] | 0.16 | 5368 | 0.232 | 0.08[0.01; 0.14] | 0.02 |
| Dx adjusted | **Tau Braak staging** | 5826 | 0.16 | -0.08[-0.16; -0.01] | 0.02 | 5826 | 0.057 | 0.05[-0.06; 0.16] | 0.38 | 6388 | 0.287 | -0.05[-0.1; 0.01] | 0.08 |
| | **Neuritic plaques density** | 3796 | 0.163 | -0.02[-0.07; 0.03] | 0.42 | 4602 | 0.057 | -0.03[-0.1; 0.04] | 0.43 | 5020 | 0.289 | -0.01[-0.06; 0.03] | 0.63 |
| | **total-Tau in CSF** | 5364 | 0.16 | -0.04[-0.11; 0.02] | 0.2 | 5264 | 0.052 | -0.22[-0.32; -0.11] | 5.0E-05 | 5263 | 0.233 | -0.09[-0.14; -0.03] | 1.6E-03 |
| | **p-Tau in CSF** | 5343 | 0.161 | -0.02[-0.08; 0.05] | 0.61 | 5242 | 0.052 | -0.27[-0.37; -0.16] | 1.7E-06 | 5243 | 0.234 | -0.06[-0.12; -0.01] | 0.02 |
| | **Aβ42 in CSF** | 5443 | 0.16 | 0.07[0.01; 0.13] | 0.03 | 5342 | 0.051 | 0.08[-0.03; 0.18] | 0.15 | 5342 | 0.232 | 0.07[0.01; 0.12] | 1.0E-02 |
| Cases | **Tau Braak staging** | 4689 | 0.157 | -0.1[-0.17; -0.03] | 5.7E-03 | 4689 | 0.057 | 0.07[-0.04; 0.18] | 0.21 | 5126 | 0.283 | -0.07[-0.12; -0.01] | 0.02 |
| | **Neuritic plaques density** | 3796 | 0.162 | -0.02[-0.05; 0.02] | 0.38 | 3796 | 0.057 | -0.01[-0.06; 0.05] | 0.85 | 4124 | 0.289 | -0.01[-0.04; 0.02] | 0.39 |
| | **total-Tau in CSF** | - | 0.157 | -0.06[-0.15; 0.04] | 0.25 | - | 0.055 | -0.32[-0.48; -0.16] | 9.6E-05 | - | 0.228 | -0.14[-0.23; -0.06] | 8.2E-04 |
| | **p-Tau in CSF** | - | 0.159 | -0.02[-0.12; 0.08] | 0.67 | - | 0.055 | -0.33[-0.49; -0.17] | 8.2E-05 | - | 0.228 | -0.1[-0.19; -0.01] | 0.02 |
| | **Aβ42 in CSF** | - | 0.159 | 0.10[0.02; 0.18] | 0.02 | - | 0.055 | -0.01[-0.14; 0.12] | 0.85 | - | 0.227 | 0.09[0.02; 0.16] | 1.0E-02 |
| | **Age-at-AD-onset** | 11315 | 0.152 | -0.07[-0.54; 0.41] | 0.78 | 11315 | 0.054 | 0.89[0.15; 1.63] | 0.02 | 11900 | 0.278 | 0.39[0.03; 0.76] | 0.03 |
| Controls | **Tau Braak staging** | 1137 | 0.173 | 0.08[-0.12; 0.28] | 0.46 | 1137 | 0.058 | -0.14[-0.46; 0.18] | 0.39 | 1262 | 0.303 | 0.06[-0.09; 0.2] | 0.45 |
| | **total-Tau in CSF** | - | 0.171 | 0.05[-0.05; 0.15] | 0.31 | - | 0.047 | -0.24[-0.42; -0.07] | 6.6E-03 | - | 0.244 | 0.01[-0.08; 0.09] | 0.85 |
| | **p-Tau in CSF** | - | 0.171 | 0.06[-0.04; 0.17] | 0.25 | - | 0.047 | -0.26[-0.44; -0.08] | 5.5E-03 | - | 0.246 | 0.03[-0.06; 0.12] | 0.56 |
| | **Aβ42 in CSF** | - | 0.17 | 0.05[-0.07; 0.16] | 0.42 | - | 0.046 | 0.16[-0.04; 0.35] | 0.11 | - | 0.243 | 0.06[-0.04; 0.15] | 0.25 |

**Supplementary Table 7. Association of HLA haplotypes in linkage with *HLA-DRB4**01:03.** Lack of association of DRB1*07:01~DQA1*02:01~DQB1*02:02 and DRB1*09:01~DQA1*03:02~DQB1*03:03 advocates against HLA-DRB4*01:03 involvement in the protective effect observed in AD and PD. Effect sizes are reported as odds ratio (OR), with 95% confidence interval [CI], and significance (p-value). FreqC: frequency of carriers, N: number of individuals

| | | Parkinson's Disease | | | | Alzheimer's Disease | | | | AD + PD | | |
|---|---|---|---|---|---|---|---|---|---|---|---|---|
| HLA-DRB3/4/5 | HLA haplotype | FreqC | N | OR | pval | FreqC | N | OR | pval | OR | pval | p_het |
| DRB4*01:03 | DRB1_04_01.DQA1_03_01.DQB1_03_02 | 0.088 | 1473386 | 0.95[0.92; 0.98] | 3.6E-03 | 0.097 | 521560 | 0.91[0.88; 0.94] | 1.1E-06 | 0.93[0.91; 0.96] | 5.2E-08 | 0.11 |
| DRB4*01:03 | DRB1_04_01.DQA1_03_03.DQB1_03_01 | 0.12 | 1481518 | 0.98[0.97; 0.99] | 5.2E-04 | 0.146 | 532448 | 0.97[0.94; 1.0] | 0.08 | 0.98[0.97; 0.99] | 1.1E-04 | 0.66 |
| DRB4*01:03 | DRB1_04_02.DQA1_03_01.DQB1_03_02 | 0.019 | 1478393 | 0.92[0.85; 0.99] | 0.04 | 0.019 | 85940 | 0.96[0.86; 1.08] | 0.53 | 0.93[0.88; 1.0] | 0.04 | 0.50 |
| DRB4*01:03 | DRB1_04_03.DQA1_03_01.DQB1_03_02 | 0.077 | 35369 | 0.82[0.73; 0.92] | 9.6E-04 | 0.077 | 10995 | 1.06[0.92; 1.22] | 0.42 | 0.91[0.83; 1.0] | 0.04 | 0.01 |
| DRB4*01:03 | DRB1_04_04.DQA1_03_01.DQB1_03_02 | 0.077 | 1475734 | 0.84[0.8; 0.89] | 1.7E-10 | 0.092 | 524205 | 0.85[0.82; 0.89] | 3.1E-14 | 0.85[0.82; 0.88] | 3.5E-23 | 0.80 |
| DRB4*01:03 | DRB1_04_05.DQA1_03_03.DQB1_04_01 | 0.11 | 35369 | 0.99[0.92; 1.06] | 0.76 | 0.227 | 10995 | 1.08[0.98; 1.18] | 0.11 | 1.02[0.97; 1.08] | 0.46 | 0.15 |
| DRB4*01:03 | DRB1_04_06.DQA1_03_01.DQB1_03_02 | 0.047 | 35369 | 0.95[0.83; 1.09] | 0.45 | 0.058 | 10995 | 0.9[0.77; 1.06] | 0.21 | 0.93[0.84; 1.03] | 0.16 | 0.63 |
| DRB4*01:03 | DRB1_04_07.DQA1_03_01.DQB1_03_02 | 0.198 | 1498 | 0.58[0.44; 0.76] | 6.1E-05 | 0.063 | 1865 | 0.75[0.47; 1.21] | 0.23 | 0.62[0.49; 0.78] | 4.6E-05 | 0.36 |
| DRB4*01:03 | DRB1_04_07.DQA1_03_03.DQB1_03_01 | 0.021 | 524845 | 0.87[0.73; 1.04] | 0.12 | 0.019 | 519510 | 0.88[0.81; 0.96] | 4.0E-03 | 0.88[0.81; 0.95] | 1.1E-03 | 0.93 |
| DRB4*01:03 | DRB1_04_10.DQA1_03_03.DQB1_04_02 | 0.039 | 5007 | 0.91[0.67; 1.24] | 0.55 | 0.037 | 6846 | 1.24[0.96; 1.6] | 0.1 | 1.09[0.9; 1.33] | 0.37 | 0.13 |
| DRB4*01:03 (2/3) /DRB4*01:01 (1/3) | DRB1_07_01.DQA1_02_01.DQB1_02_02 | 0.201 | 1506744 | 1.01[0.97; 1.05] | 0.65 | 0.186 | 92453 | 1.01[0.97; 1.04] | 0.75 | 1.01[0.98; 1.03] | 0.58 | 0.92 |
| DRB4*01:03N | DRB1_07_01.DQA1_02_01.DQB1_03_03 | 0.08 | 1485027 | 1.2[1.12; 1.28] | 2.1E-07 | 0.065 | 95130 | 1.08[1.02; 1.14] | 8.1E-03 | 1.12[1.08; 1.17] | 9.4E-08 | 0.02 |
| DRB4*01:03 | DRB1_09_01.DQA1_03_02.DQB1_03_03 | 0.03 | 1510253 | 1.02[0.97; 1.07] | 0.43 | 0.048 | 96309 | 1.04[0.97; 1.11] | 0.29 | 1.03[0.99; 1.07] | 0.2 | 0.70 |

**Supplementary Table 8.** Queried US based cohorts' part of the Alzheimer's disease ADSP and ADGC analyses.

| Cohort/Project | Genotyping Platform | Cohort-Platform ID | Sample (N) | Data Repository and Access ID |
|---|---|---|---|---|
| ADSP WGS | Whole Genome Sequencing | ADSP_WGS | 16906 | NIAGADS DSS (NG00067.v5) / NACC |
| ACT | Illumina Human 660W-Quad | ACT | 2790 | NIAGADS (NG00034) / dbGaP (phs000234) |
| ADC1 | Illumina Human 660W-Quad | ADC1 | 2731 | NIAGADS (NG00022) / NACC |
| ADC2 | Illumina Human 660W-Quad | ADC2 | 928 | NIAGADS (NG00023) / NACC |
| ADC3 | Illumina Human OmniExpress | ADC3 | 1526 | NIAGADS (NG00024) / NACC |
| ADC4 | Illumina Human OmniExpress | ADC4 | 1054 | NIAGADS (NG00068) / NACC |
| ADC5 | Illumina Human OmniExpress | ADC5 | 1224 | NIAGADS (NG00069) / NACC |
| ADC6 | Illumina Human OmniExpress | ADC6 | 1333 | NIAGADS (NG00070) / NACC |
| ADC7 | Illumina Infinium Human OmniExpressExome | ADC7 | 1462 | NIAGADS (NG00071) / NACC |
| ADDNEUROMED | Illumina Human 610-Quad | ADM_Q | 315 | Synapse AddNeuroMed (syn4907804) |
| | Illumina Human OmniExpress | ADM_O | 329 | Synapse AddNeuroMed (syn4907804) |
| ADNI | Illumina Human 610-Quad | ADNI_Q | 757 | LONI ADNI |
| | Illumina Human OmniExpress | ADNI_OE | 361 | LONI ADNI |
| | Illumina Omni 2.5 | ADNI_O25 | 812 | LONI ADNI |
| | Illumina Human OmniExpress | ADNI_DOD | 204 | LONI ADNIDOD |
| ADNI3 | Illumina Global Screening Array (GSA) | ADNI3 | 327 | LONI ADNI |
| IIDP African Americans | Illumina Human 1M-Duo | IIDP_AA | 1175 | NIAGADS (NG00047) |
| IIDP Yorubans | Illumina Human 1M-Duo | IIDP_YOR | 1264 | NIAGADS (NG00047) / cf. gaaindata.org/partner/IIDP |
| CIDR | Illumina Human Omni1-Quad | CIDR | 3101 | NIAGADS (NG00015) / dbGAP (phs000160) |
| GenADA | Affymetrix 500K | GSK | 1571 | dbGaP (phs000219) |
| LATC | Illumina Multi-Ethnic – BU | LATC | 63 | RADC Rush / Latino CORE Study |
| NIA-LOAD | Illumina Human 610-Quad | LOAD | 5220 | NIAGADS (NG00020) |
| MARS | Illumina Multi-Ethnic – BU | MARS | 708 | RADC Rush / Minority Aging Research Study |
| MAYO | Illumina Human Hap300 | MAYO_1 | 2099 | Synapse AMP-AD (syn5591675) |

| | | | | |
|---|---|---|---|---|
| MAYO2 | Illumina Omni 2.5 | MAYO_2 | 314 | Synapse AMP-AD (syn5550404) |
| MIRAGE | Illumina Human CNV370-Duo | MIRAGE_370 | 397 | NIAGADS (NG00031) |
| | Illumina Human 610-Quad | MIRAGE_610 | 1105 | NIAGADS (NG00031) |
| MTC | Illumina Human OmniExpress | MTC | 542 | NIAGADS (NG00096) |
| OHSU | Illumina Human CNV370-Duo | OHSU | 647 | NIAGADS (NG00017) |
| ROSMAP | Affymetrix GeneChip 6.0 - Broad Institute | ROSMAP_1B | 1126 | RADC Rush / Synapse AMP-AD (syn3219045) |
| | Affymetrix GeneChip 6.0 - TGen | ROSMAP_1T | 582 | RADC Rush / Synapse AMP-AD (syn3219045) |
| | Illumina Human OmniExpress 12 - Chop | ROSMAP_2C | 382 | RADC Rush / Synapse AMP-AD (syn7824841) |
| | Illumina Multi-Ethnic - BU | ROSMAP_3BU | 494 | RADC Rush |
| TARCC | Affymetrix 6.0 | TARCC | 2718 | NIAGADS (NG00097) / TARCC study |
| TGEN2 | Affymetrix 6.0 | TGEN | 1599 | NIAGADS (NG00028) |
| UPITT | Illumina Human Omni1-Quad | UPITT | 2440 | NIAGADS (NG00026) |
| UM-VU-MSSM | Illumina Human 1M-Duo, Illumina 1M | UVM_A | 1153 | NIAGADS (NG00042) |
| | Affymetrix 6.0 | UVM_B | 864 | NIAGADS (NG00042) |
| | Illumina Human 550K. Illumina Human 610-Quad | UVM_C | 445 | NIAGADS (NG00042) |
| WASHU | Illumina Human 610-Quad | WASHU_1 | 670 | NIAGADS (NG00030) |
| WASHU2 | Illumina Human OmniExpress | WASHU_2 | 235 | NIAGADS (NG00087) |
| WHICAP | Illumina Human OmniExpress | WHICAP | 647 | NIAGADS (NG00093) |

**Supplementary Table 9.** Demographics of the cohorts queried among the ADSP and ADGC in-house analyses in Alzheimer's disease.
AFR: African, AMR: American (central and south; admixed), EAS: East Asian, SAS South Asian, EUR: European, otherwise ADMIX: admixed of these super ancestry categories.

| | | Ancestry | | | | | | Diagnosis | | Sex - Females | | Age | |
|---|---|---|---|---|---|---|---|---|---|---|---|---|---|
| Cohort | N total | AFR N | ADMIX N | AMR N | EAS N | SAS N | EUR N | CN N | AD N | CN N(%) | AD N(%) | CN μ(σ) | AD μ(σ) |
| ADSP WGS | 16906 | 2240 | 4012 | 58 | 68 | 19 | 10509 | 6717 | 6434 | 4510(67.1) | 3896(60.6) | 78.2(8.5) | 74.1(10.5) |
| ACT | 2790 | 70 | 64 | 7 | 73 | 0 | 2576 | 1833 | 713 | 1000(54.6) | 462(64.8) | 82.9(6.5) | 82.1(6.6) |
| ADC1 | 2731 | 92 | 58 | 47 | 20 | 0 | 2514 | 603 | 1946 | 354(58.7) | 1039(53.4) | 79.8(10.8) | 70.7(9.5) |
| ADC2 | 928 | 0 | 2 | 0 | 0 | 0 | 926 | 124 | 707 | 87(70.2) | 366(51.8) | 80.1(9.2) | 72.9(7.1) |
| ADC3 | 1526 | 0 | 5 | 0 | 0 | 0 | 1521 | 482 | 858 | 305(63.3) | 468(54.5) | 79.6(9.6) | 72.5(10.3) |
| ADC4 | 1054 | 6 | 10 | 1 | 0 | 0 | 1037 | 420 | 452 | 257(61.2) | 237(52.4) | 79.2(8.7) | 72.6(9.0) |
| ADC5 | 1224 | 0 | 1 | 0 | 0 | 0 | 1223 | 579 | 415 | 376(64.9) | 226(54.5) | 82.0(8.9) | 74.1(8.7) |
| ADC6 | 1333 | 0 | 2 | 0 | 0 | 0 | 1331 | 352 | 567 | 238(67.6) | 304(53.6) | 80.1(8.9) | 66.9(12.0) |
| ADC7 | 1462 | 0 | 4 | 0 | 0 | 0 | 1458 | 763 | 536 | 493(64.6) | 281(52.4) | 78.0(7.9) | 72.8(7.7) |
| ADDNEURO | 644 | 0 | 2 | 0 | 0 | 0 | 642 | 186 | 256 | 105(56.5) | 164(64.1) | 76.4(6.6) | 73.0(6.7) |
| ADNI | 2134 | 63 | 69 | 21 | 30 | 5 | 1945 | 606 | 761 | 260(42.9) | 330(43.4) | 78.5(7.8) | 74.1(7.4) |
| ADNI3 | 327 | 4 | 12 | 1 | 4 | 0 | 306 | 228 | 24 | 142(62.3) | 10(41.7) | 72.5(6.1) | 72.7(9.5) |
| CIDR | 3101 | 93 | 2780 | 70 | 0 | 0 | 158 | 1505 | 1530 | 1033(68.6) | 986(64.4) | 74.5(9.4) | 75.5(9.6) |
| GSK | 1571 | 0 | 1 | 1 | 0 | 0 | 1569 | 773 | 798 | 497(64.3) | 459(57.5) | 73.4(7.9) | 72.5(8.6) |
| IIDP AA | 1175 | 815 | 359 | 0 | 0 | 0 | 1 | 1001 | 172 | 663(66.2) | 107(62.2) | 83.3(5.3) | 83.6(6.7) |
| IIDP YOR | 1264 | 1253 | 10 | 0 | 0 | 0 | 1 | 1145 | 104 | 732(63.9) | 79(76.0) | 82.6(5.9) | 77.9(7.2) |
| LATC | 63 | 13 | 23 | 24 | 0 | 0 | 0 | 15 | 2 | 15(100.0) | 2(100.0) | 77.4(5.4) | 78.0(0.0) |
| MARS | 708 | 423 | 275 | 1 | 0 | 0 | 1 | 463 | 79 | 392(84.7) | 54(68.4) | 79.6(6.1) | 77.3(7.1) |
| MAYO | 2413 | 7 | 24 | 2 | 4 | 0 | 2335 | 1225 | 948 | 642(52.4) | 546(57.6) | 75.5(6.5) | 74.0(6.0) |
| MIRAGE | 1502 | 1 | 28 | 2 | 0 | 0 | 1471 | 738 | 601 | 436(59.1) | 366(60.9) | 72.1(7.3) | 68.8(8.6) |
| MTC | 542 | 5 | 29 | 12 | 0 | 0 | 496 | 202 | 272 | 130(64.4) | 157(57.7) | 71.7(8.9) | 72.6(9.3) |
| NIA-LOAD | 5220 | 112 | 642 | 13 | 8 | 0 | 4445 | 2091 | 2351 | 1278(61.1) | 1546(65.8) | 70.6(12.6) | 73.6(7.8) |
| OHSU | 647 | 3 | 2 | 0 | 1 | 0 | 635 | 379 | 201 | 205(54.1) | 127(63.2) | 85.7(7.5) | 85.0(6.9) |
| ROSMAP | 2584 | 13 | 50 | 28 | 9 | 0 | 2451 | 1102 | 951 | 795(72.1) | 690(72.6) | 85.4(7.4) | 84.1(6.5) |
| TARCC | 2718 | 75 | 218 | 821 | 7 | 2 | 1557 | 1124 | 908 | 788(70.1) | 502(55.3) | 70.1(9.8) | 70.1(8.9) |
| TGEN2 | 1599 | 0 | 9 | 1 | 0 | 1 | 1512 | 573 | 1005 | 255(44.5) | 640(63.7) | 80.8(8.7) | 72.8(8.0) |
| UM-VU-MSSM | 2462 | 5 | 16 | 0 | 0 | 0 | 2441 | 1195 | 1206 | 724(60.6) | 778(64.5) | 74.1(8.2) | 74.2(7.9) |
| UPITT | 2440 | 7 | 8 | 1 | 0 | 0 | 2355 | 896 | 1406 | 563(62.8) | 908(64.6) | 75.6(6.2) | 73.2(6.6) |
| WASHU | 670 | 0 | 0 | 0 | 0 | 0 | 670 | 202 | 429 | 125(61.9) | 239(55.7) | 77.9(8.7) | 74.0(9.6) |
| WASHU2 | 235 | 10 | 1 | 0 | 0 | 0 | 224 | 116 | 68 | 65(56.0) | 38(55.9) | 73.7(8.6) | 74.0(8.1) |
| WHICAP | 647 | 0 | 7 | 0 | 0 | 0 | 640 | 554 | 85 | 335(60.5) | 60(70.6) | 82.7(6.7) | 84.1(7.5) |

**Supplementary Table 10. Demographics by ancestry of ADSP and ADGC individuals included in the analyses.** AAD: age-at death, AAL: age-at-last-exam, AAE: age-at-exam, AAO: age-at-onset.

| | | | Sex | Age | Age Type | | | |
|---|---|---|---|---|---|---|---|---|
| Cohort | Diagnosis | N | Female (%) | Age μ(σ) | AAD | AAL | AAE | AAO |
| African ancestry ADSP WGS | AD | 849 | 70.0% | 75.0(8.8) | - | - | 77.0(7.7)[1.1%] | 74.9(8.8)[98.8%] |
| | CN | 1240 | 75.1% | 75.7(9.0) | 82.5(8.9)[5.6%] | 75.3(8.9)[94.3%] | - | - |
| European ancestry ADSP WGS | AD | 4127 | 56.6% | 73.8(11.0) | 69.0(-)[0.0%] | 86.2(4.1)[0.1%] | 78.8(8.5)[20.0%] | 72.5(11.2)[79.2%] |
| | CN | 3020 | 60.8% | 81.2(7.2) | 85.2(7.2)[23.4%] | 80.0(6.8)[76.2%] | - | - |
| Amerindian-Latino ancestry ADSP WGS | AD | 355 | 69.9% | 76.4(10.0) | - | - | 73.4(9.1)[2.8%] | 76.5(10.0)[92.4%] |
| | CN | 1366 | 69.2% | 74.6(7.3) | 88.0(7.0)[0.3%] | 74.5(7.3)[82.1%] | - | - |
| African ancestry ADGC imputed | AD | 253 | 74.3% | 76.8(9.3) | 74.2(3.3)[2.0%] | - | 79.2(8.4)[64.8%] | 72.3(9.6)[32.0%] |
| | CN | 1837 | 65.8% | 81.9(6.6) | 80.3(8.0)[2.6%] | 81.9(6.5)[97.4%] | - | - |
| European ancestry ADGC imputed | AD | 7151 | 58.3% | 74.2(9.9) | 71.5(8.1)[13.0%] | 85.9(6.4)[0.7%] | 83.9(6.9)[11.4%] | 73.0(9.7)[74.9%] |
| | CN | 3277 | 54.0% | 83.5(9.1) | 83.5(9.1)[100.0%] | - | - | - |
| Amerindian-Latino ancestry ADGC imputed | AD | 1051 | 64.6% | 74.3(10.2) | 78.0(2.8)[0.2%] | 87.2(8.3)[0.6%] | 76.4(9.7)[0.5%] | 74.2(10.2)[98.8%] |
| | CN | 1354 | 70.4% | 69.9(10.1) | 81.4(7.5)[1.1%] | 69.8(10.1)[98.9%] | - | - |

**Supplementary Table 11. Demographic descriptions of the different meta-analyzed GWAS from Bellenguez et al. and subset included in the HLA -level analysis (EADB, GR@ACE, GERAD, EADI, DemGene, Bonn, CCHS).**

| | AD or proxy-ADD cases | | | | | Controls | | | |
|---|---|---|---|---|---|---|---|---|---|
| | N | % females | Age | Age at onset | APOE e4 allele frequency | N | % females | Age | APOE e4 allele frequency |
| **EADB-TOPMed** | 20,301 | 61.7 | 72.0±10.4 | *71.1±10.5* | 32.6 | 21,839 | 57.3 | 67.0±14.3 | 13.2 |
| *Belgium* | *1,230* | *64.6* | *78.7±5.9* | *78.3±5.9* | *31.6* | *1,474* | *61.8* | *70.1±8.4* | *13.6* |
| *Bulgaria* | *164* | *54.9* | *65.0±8.6* | *65.1±8.6* | *22.9* | - | - | - | - |
| *Switzerland* | *182* | *64.3* | *76.0±6.6* | *76.9±6.0* | *19.2* | *388* | *55.9* | *74.8±4.0* | *10.1* |
| *Czech Republic* | *183* | *60.7* | *75.8±7.8* | - | *31.7* | *61* | *65.6* | *66.9±7.2* | *10.7* |
| *Denmark* | *403* | *57.1* | *79.6±7.8* | *79.6±7.8* | *33.7* | *654* | *54.4* | *73.1±8.5* | *15.4* |
| *Spain* | *3,273* | *67.0* | *75.3±9.0* | *75.2±9.0* | *27.2* | *1,685* | *63.3* | *69.3±12.0* | *10.0* |
| *Finland* | *1,151* | *64.0* | *70.9±8.8* | *69.8±8.5* | *42.0* | *1,806* | *51.4* | *71.8±7.1* | *15.9* |
| *France* | *1,664* | *60.2* | *67.4±11.9* | *63.2±10.8* | *33.3* | *3,106* | *63.8* | *44.9±15.4* | *11.5* |
| *Germany* | *1,628* | *60.3* | *74.8±9.4* | *74.6±9.8* | *33.1* | *2,050* | *56.1* | *74.2±8.0* | *12.3* |
| *Greece* | *614* | *63.0* | *73.1±8.0* | *72.9±8.0* | *23.8* | *1,246* | *57.3* | *73.1±5.6* | *9.2* |
| *Italy* | *3,271* | *68.1* | *73.7±8.9* | *72.2±8.7* | *25.0* | *1,317* | *56.8* | *72.2±10.5* | *8.6* |
| *The Netherlands* | *2,438* | *55.8* | *66.2±10.7* | *65.6±10.5* | *41.9* | *2,389* | *47.5* | *60.1±12.0* | *17.9* |
| *Portugal* | *80* | *75.0* | *69.9±9.2* | *69.2±8.9* | *30.0* | *74* | *75.7* | *67.2±6.8* | *17.6* |
| *Sweden* | *1,533* | *63.0* | *72.8±11.2* | *72.8±11.2* | *40.7* | *3,089* | *61.8* | *70.6±9.8* | *15.6* |
| *United Kingdom* | *2,487* | *51.1* | *68.1±10.7* | *66.4±10.1* | *34.4* | *2,500* | *51.8* | *74.4±7.2* | *12.8* |
| **EADB-HRC** | 163 | 54.0 | 71.5±7.9 | 71.5±7.9 | 31.8 | 405 | 48.2 | 77.2±2.1 | 14.1 |
| **EADI** | 2,400 | 65.6 | 74.3±10.1 | 73.9±10.1 | 29.4 | 6,338 | 60.3 | 80.0±7.6 | 10.5 |
| **GERAD** | 3,030 | 63.2 | 78.1±9.3 | 77.8±9.3 | 35.1 | 7,153 | 51.2 | 50.7±11.7 | 15.4 |
| **Bonn** | 635 | 65.5 | 77.8±9.8 | 77.8±9.3 | 30.1 | 1,210 | 54.8 | 69.9±9.3 | 12.6 |
| **RS1** | 1,165 | 72.9 | 83.7±0.2 | 83.7±0.2 | 33.4 | 4,739 | 56.7 | 82.8±0.1 | 12.9 |
| **RS2** | 141 | 59.6 | 82.8±0.6 | 82.8±0.6 | 27.1 | 1,961 | 54.1 | 73.3±0.2 | 14.1 |
| **GR@ACE/DEGESCO** | 6,497 | 64.1 | 81.8±8.8 | 81.8±8.8 | 23.0 | 6,785 | 49.1 | 55.9±15.8 | 11.0 |
| **DemGene** | 1,693 | 65.5 | 72.2±8.8 | 71.6±8.8 | 39.5 | 5,926 | 47.7 | 68.5±11.1 | 18.2 |
| **CCHS** | 365 | 68.5 | 82.7±6.9 | 82.7±6.9 | 31.3 | 6,106 | 54.3 | 58.5±13.7 | 15.8 |
| **NxC** | 269 | 72.4 | 78.7±6.9 | 78.7±6.9 | 26.0 | 675 | 44.4 | 51.9±8.9 | 10.0 |

**Supplementary Table 12. Demographics by ancestry of ROSMAP and ADGC individuals included in the neuropathology analyses.** AAD: age-at death, AAL: age-at-last-exam, AAE: age-at-exam, AAO: age-at-onset.

| Phenotype | Diagnosis | N | Phenotype μ(σ) | Sex Female (%) | Age Age μ(σ) | Age Type AAD | AAL | AAE | AAO |
|---|---|---|---|---|---|---|---|---|---|
| Tau Braak staging | AD | 5148 | 5.14(0.91) | 2926(56.8%) | 73.5(10.2) | 71.0(8.1)[12.5%] | 87.0(5.0)[0.4%] | 83.8(7.0)[14.2%] | 71.8(9.8)[72.9%] |
| | CN | 1262 | 2.44(1.32) | 733(58.1%) | 86.3(7.7) | 86.3(7.7)[100.0%] | - | - | - |
| | MCI/Others | 1072 | 2.98(1.62) | 553(51.6%) | 79.8(10.1) | 76.9(8.3)[10.3%] | 84.5(7.2)[4.3%] | 83.8(6.8)[22.7%] | 78.2(11.3)[49.8%] |
| Neuritic plaques density | AD | 4146 | 2.77(0.42) | 2279(55.0%) | 73.6(10.5) | 69.7(3.1)[0.1%] | 86.8(5.1)[0.4%] | 84.1(6.8)[15.8%] | 71.5(9.9)[83.7%] |
| | CN | 896 | 1.04(1.10) | 543(60.6%) | 87.7(7.4) | 87.7(7.4)[100.0%] | - | - | - |
| | MCI/Others | 860 | 0.86(0.92) | 447(52.0%) | 80.5(10.4) | 85.2(7.4)[3.3%] | 84.5(7.2)[5.3%] | 83.8(6.8)[28.3%] | 78.2(11.5)[57.4%] |

**Supplementary Table 13. Demographics by ancestry of ROSMAP and ADGC individuals included in the dual-pathology analyses.** AAD: age-at death. AD-LB-: without AD and LB pathology, AD+LB-: only AD pathology without LB pathology, AD-LB+: only LB pathology without AD pathology, AD+LB+: dual pathology (AD and LB).

| | | Sex | AAD |
|---|---|---|---|
| **Phenotype** | **N** | **Female (%)** | **Age μ(σ)** |
| AD-LB- | 1290 | 722(56.0%) | 87.5(8.2) |
| AD+LB- | 2641 | 1478(56.0%) | 83.0(9.4) |
| AD-LB+ | 298 | 146(49.0%) | 87.0(8.2) |
| AD+LB+ | 889 | 453(51.0%) | 82.4(9.3) |

**Supplementary Table 14. Demographic information on cohorts of Cerebrospinal Fluid analyses.**

| | | | | | Diagnoses | | | | |
|---|---|---|---|---|---|---|---|---|---|
| **Country** | **Cohort** | **n** | **age (sd)** | **male** | **AD** | **MCI** | **control** | **other dem** | **data type** |
| Belgium | DEM | 587 | 78.3 (7.3) | 40% | 72% | 27% | 1% | 0% | Genotype level |
| Finland | ADGEN | 226 | 70.2 (8.0) | 34% | 89% | 1% | 10% | 0% | Genotype level |
| France1 | BALTAZAR | 420 | 77.0 (6.7) | 45% | 43% | 57% | 0% | 0% | Genotype level |
| France2 | MEMENTO | 389 | 69.2 (8.9) | 47% | 0% | 100% | 0% | 0% | Summary statistics |
| France3 | CNRMAJ-Rouen | 127 | 66.0 (8.7) | 47% | 100% | 0% | 0% | 0% | Genotype level |
| Germany1 | Delcode | 465 | 71.7 (5.9) | 52% | 13% | 23% | 64% | 0% | Summary statistics |
| Germany2 | KND | 309 | 67.3 (8.7) | 57% | 18% | 82% | 0% | 0% | Genotype level |
| Germany3 | TUM | 151 | 70.2 (9.2) | 48% | 98% | 1% | 0% | 1% | Genotype level |
| Germany4 | PAGES | 136 | 73.4 (7.7) | 40% | 70% | 30% | 0% | 0% | Genotype level |
| Germany5 | UHB | 111 | 70.3 (7.2) | 42% | 69% | 30% | 1% | 0% | Genotype level |
| Netherlands | ADC & Pearl ND | 2936 | 64.1 (8.9) | 59% | 42% | 10% | 24% | 23% | Genotype level |
| Spain1 | ACE | 609 | 72.7 (8.2) | 43% | 27% | 59% | 8% | 6% | Summary statistics |
| Spain2 | SIGNAL & SPIN | 394 | 70.6 (8.0) | 43% | 34% | 45% | 19% | 2% | Genotype level |
| Spain3 | Valdecilla | 98 | 67.0(9.0) | 39% | 10% | 37% | 45% | 8% | Summary statistics |
| Sweden1 | Birth cohort & Clin. AD | 856 | 75.0 (9.4) | 45% | 51% | 0% | 49% | 0% | Genotype level |
| Sweden2 | Uppsala university | 260 | 71.0 (6.3) | 46% | 58% | 37% | 0% | 6% | Genotype level |

DEM=Antwerp prospective dementia cohort. All, except the Swedish Birth cohort & clinical AD samples, are part of EADB.

**Supplementary Table 15. Demographics for the IPDGC, McGill, NINDS, NGRC, Oslo, PPMI, APDGC, UK Biobank datasets included in Yu et al. (2021) analysis[9].**

| Cohort | Diagnosis | N | Female (%) | Age at onset | Age at recruitment or last visit |
|---|---|---|---|---|---|
| APDGC | PD | 603 | 27.7 | 77.4 (8.4) | |
| | CN | 295 | 50.3 | | 81.9 (12.7) |
| IPDGC | PD | 5163 | 35.6 | 61.2 (12.6) | |
| | CN | 5389 | 44.2 | | 64.3 (14.8) |
| McGill | PD | 1240 | 36.8 | 58.5 (10.6) | |
| | CN | 995 | 49.0 | | 44.15 (14.7) |
| NINDS | PD | 847 | 40.4 | 66.1 (11.1) | |
| | CN | 773 | 58.1 | | 58.7 (16.4) |
| NGRC | PD | 1922 | 32.6 | 58.3 (12.0) | |
| | CN | 1938 | 61.2 | | 70.3 (14.1) |
| Oslo | PD | 474 | 35.9 | 55.7 (11.3) | |
| | CN | 459 | 42.5 | | 61.8 (11.0) |
| PPMI | PD | 398 | 33.3 | 59.8 (9.9) | |
| | CN | 159 | 33.5 | | 61.1 (10.7) |
| UKB Proxy | Proxy-PD | 14422 | 56.8 | | 58.5 (7.1) |
| | CN | 308694 | 53.6 | | 56.8 (8.0) |
| UKB cases | PD | 1490 | 37.1 | | 62.8 (5.4) |
| | CN | 33251 | 53.7 | | 56.8 (8.0) |

**Supplementary Table 16. Demographics for the 23andMe, East-Asians-PD, and LARGE-PD datasets.** Sex and Age were partly not available for the 23andMe individuals.

| | | | Sex | Age |
|---|---|---|---|---|
| **Cohort** | **Diagnosis** | **N** | **Female (%)** | **Age μ(σ)** |
| European ancestry | PD | 3261 | - | - |
| 23andMe | CN | 29499 | - | - |
| European ancestry | PD | 866 | - | - |
| 23andMe | CN | 32538 | - | - |
| European ancestry | PD | 6476 | 38.9% | - |
| 23andMe | CN | 302042 | 48.2% | - |
| European ancestry | PD | 2448 | 39.1% | - |
| 23andMe | CN | 571411 | 54.9% | - |
| East Asian ancestry | PD | 988 | 55.0% | 58.8(10.1) |
| Japaneses (Naito et al. 2021) | CN | 2521 | 45.2% | 49.9(14.2) |
| East Asian ancestry | PD | 2279 | 42.9% | 59.7(10.7) |
| Chineses (Foo et al., 2020) | CN | 2021 | 45.0% | 52.6(13.5) |
| East Asian ancestry | PD | 2536 | 43.0% | 66.6(9.7) |
| Singaporeans/Malays (Foo et al., 2020) | CN | 21840 | 55.2% | 60.8(7.6) |
| East Asian ancestry | PD | 1494 | 54.2% | 66.1(9.4) |
| South Koreans (Foo et al., 2020) | CN | 599 | 56.4% | 71.0(9.3) |
| East Asian ancestry | PD | 199 | 35.2% | 63.8(10.0) |
| Hong-Kongers (Foo et al., 2020) | CN | 166 | 32.5% | 61.9(7.9) |
| East Asian ancestry | PD | 216 | 51.9% | 72.4(9.5) |
| Taiwaneses (Foo et al., 2020) | CN | 225 | 55.6% | 72.4(7.7) |
| Amerindian/European-Latinos | PD | 807 | 47.0% | 61.7(12.8) |
| South Americans (Loesch et al., 2021) | CN | 690 | 67.0% | 56.5(14.6) |

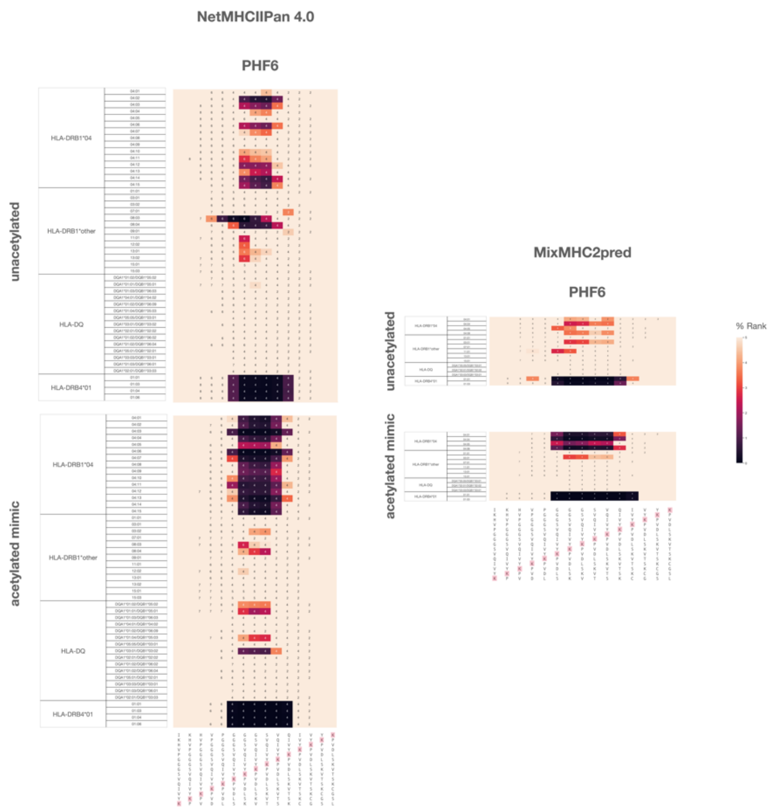

**Supplementary Figure 1. HLA binding predictions for PHF6*.** Predictions were made using NetMHCIIPan 4.0[31] (left) using Mixed MHC pred 2 Server[32] (right). Cmap indicates a percentile rank generated by comparing the peptide's score against the scores of five million random 15 mers selected from SWISSPROT database (best score = 0%; worst score = 100%). 15mer sequences incorporating 15mer sequences incorporating PHF6* in its unacetylated (bottom) or K2380 mimic acetylated (top) form . Annotations within heat map show the position of the lysines of interest (K280 for PHF6*, see highlighted sequences) within the 9mer core that is predicted to bind to the HLA molecule. 9mer cores excluding the lysine of interest were not predicted for binding and left blank.

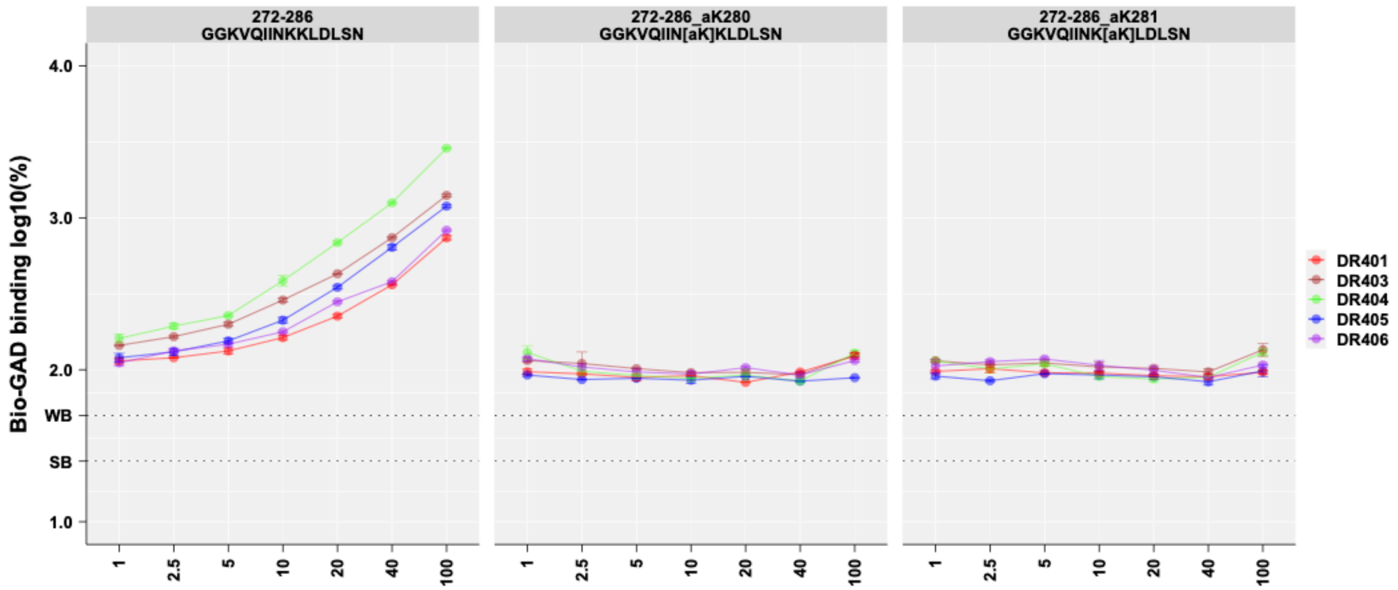

**Supplementary Figure 2. Dose response of PHF6* peptides**. Each Tau peptide at different concentrations competed with biotinylated peptide binding to *HLA-DRB1**04:01, *HLA-DRB1**04:03, *HLA-DRB1**04:04, *HLA-DRB1**04:05, and *HLA-DRB1**04:06. Tau peptide with fluorescence that was lower than 25% and 25-50% of biotinylated peptide was considered strong binder (SB) and weak binder (WB), respectively.

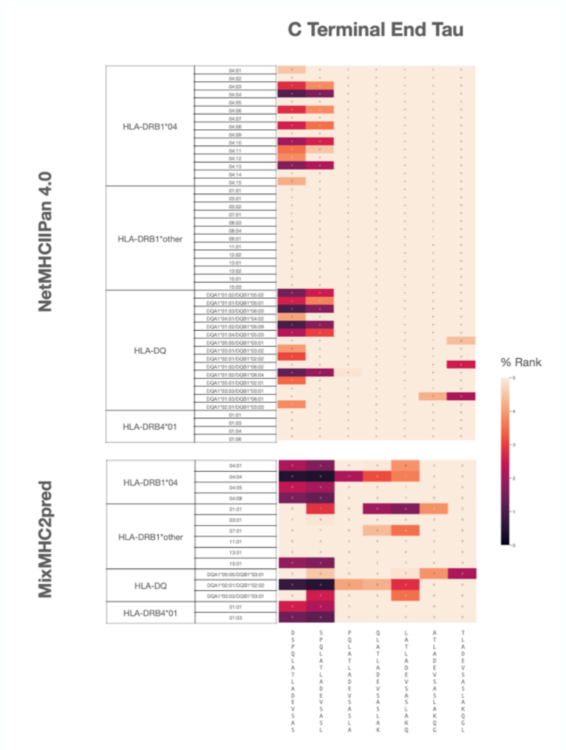

**Supplementary Figure 3. HLA binding predictions for Tau C terminal end.** The predicted binding of 15mer sequences of the C terminal end of tau for various HLA molecules is shown using (a) NetMHCIIPan 4.0 [31]. Cmap indicates a percentile rank generated by comparing the peptide's score against the scores of five million random 15 mers selected from SWISSPROT database (best score = 0%; worst score = 100%). (b) Mixed MHC pred 2 Server [32]. Cmap indicates the percent of random peptides that would have a score higher than the peptide provided among peptides of sizes 12-25 amino acids (best score = 0%; worst score = 100%).
